# Development and validation of an ECG-based 10-year risk prediction model for Major Adverse Cardiac and Cerebrovascular Events in UK Biobank

**DOI:** 10.64898/2026.03.12.26347852

**Authors:** Adam Sturge, Stefan van Duijvenboden, Barbara Casadei, Aiden Doherty

## Abstract

**Background:** Electrocardiograms (ECG) are commonly used to diagnose heart conditions, but whether they add to traditional risk factors in predicting cardiovascular disease (CVD) is currently unclear. We investigated whether ECG measurements, taken at rest, exercise, and recovery, improve the prediction of major cardiovascular and cerebrovascular events (MACCE) both independently and when added to a clinical risk model derived from QRISK3 risk factors currently used in UK primary care.

**Methods:** We obtained ECG recordings from 41,076 UK Biobank participants without a history of MACCE who underwent a submaximal cycle ergometry test. We developed two ECG risk scores: one derived from conventional ECG parameters, measured at rest, peak exercise and recovery (C-ECG), and a neural-network risk model based on raw ECG recordings (ECG_AI_). We estimated the association between ECG scores and incident MACCE, using Cox proportional hazards models adjusted for traditional risk factors. Incremental predictive value was assessed relative to a newly derived Cox clinical risk score, constructed by refitting Cox models using the QRISK3 risk factors. All models were internally validated using five-fold cross-validation and 1,000 bootstrap iterations. Predictive performance was evaluated using Harrell’s C-index and net benefit.

**Findings:** Incident MACCE was reported in 4,082 (9.9%) individuals, 3,463 (9.7%) of whom had valid ECG parameters and a median follow-up of 12.5 years. C-ECG and ECG_AI_ scores were independently associated with MACCE, with hazard ratios of 1.76 (95% CI: 1.63-1.91) and 1.18 (95% CI: 1.15-1.21) per SD increase, respectively. When added to the Cox clinical risk score, both C-ECG and ECG_AI_ scores modestly improved model discrimination, ΔC-index 0.03 (95% CI: 0.02–0.04) and ΔC-index 0.03 (95% CI: 0.02–0.03), respectively. ECG_AI_ risk scores were observed to significantly improve the categorical NRI among women (NRI = 0.09, 95% CI: 0.06–0.11) at a risk threshold of 10%, suggesting enhanced risk stratification in this subgroup.

**Interpretation:** In individuals without a history of prior MACCE, ECG-derived risk scores independently predict the 10-year risk of MACCE. However, when combined with QRISK3 risk factors, ECG risk scores only marginally improve risk prediction.

**Funding:** UK Engineering and Physical Sciences Research Council for the University of Oxford Health Data Science Centre for Doctoral Training [EP/S02428X/1] and Wellcome Trust.

## Introduction

The early identification of individuals at high risk for cardiovascular disease (CVD) is the cornerstone of effective primary prevention. Currently, cardiovascular risk prediction centres on population-tailored algorithms, such as QRISK3^1,2^, which consider traditional risk factors and are heavily influenced by non-modifiable risk factors such as age and sex. Whilst well validated for clinical use on large populations, traditional risk scores suffer from limited accuracy and considerable heterogeneity across individuals^3,4^. This is further compounded by a reliance on prior diagnosis of key risk factors, such as atrial fibrillation, which are often underdiagnosed^5^ and recorded with varying degrees of accuracy^6^. By capturing real-time, patient-specific information on myocardial structure, perfusion and electrical activity, the electrocardiogram (ECG) can provide useful insights into the impact of clinical and subclinical risk factors on the heart. While resting ECG parameters have been strongly associated with incident CVD, independent of established risk factors^7–10^, relatively few studies have evaluated whether exercise ECG adds predictive value beyond traditional risk scores for long-term CVD risk prediction. Exercise ECG testing poses a dynamic physiological stress on the heart that may unmask underlying cardiac anomalies not evident at rest^10–13^, potentially enabling improved risk stratification for primary prevention.

However, previous investigations into the role of resting and exercise ECGs for risk prediction are often limited by (i) small sample sizes, (ii) non-population-based study designs, (iii) short follow-up duration, and (iv) lack of integration with currently used clinical risk models^10^. Moreover, until recently, most investigations have relied on traditional ECG measures, which often differ between studies, lack sensitivity to subtle changes, and only explore a narrow subset of available information. By contrast, artificial intelligence (AI) techniques are able to detect subtle patterns in raw ECG data that are often not captured by traditional approaches^14–16^, regularly outperform conventional ECG markers in predicting arrhythmias, conduction abnormalities, and repolarisation markers that have been linked to increased cardiovascular risk^15,17–19^. However, little evidence exists regarding the added value of AI to the long-term prediction of incident major cardiovascular and cerebrovascular events (MACCE).

Here, we trained a neural network using raw ECG signals recorded during a submaximal exercise test, to predict the 10-year risk of MACCE in 41,076 UK Biobank participants with no prior history of MACCE using deep-survival models. We subsequently assessed the association of conventional and deep learning-derived ECG risk factors with incident MACCE and their added value to risk factors from the currently adopted UK risk assessment tool within primary care, QRISK3^1,2^. We also investigated the individual predictive performance of ECG measurements recorded during rest, exercise, and recovery, as well as their added value to QRISK3 risk factors.

## Methods

### Study design and population UK Biobank

The UK Biobank is a large, prospective cohort study of 502,636 participants (aged 40-69, 46% male) recruited across 22 assessment centres in England, Wales, and Scotland between 2006 and 2010^20^ . At recruitment, all participants provided written informed consent for passive follow-up through linkage to their health care records for multiple health-related outcomes, a complete touch screen questionnaire, verbal interview, physical measurements, and biological sampling^21,22^. The UK Biobank obtained ethical approval from the Northwest Multi-Centre Research Ethics Committee [21/NW/0157], the National Information Governance Board for Health and Social Care in England and Wales, and the Community Health Index Advisory Group in Scotland.

### Exercise ECG testing

Of the broader UK Biobank cohort, 96,524 participants consented to take part in the Cardio Assessment sub-study conducted between 2009 and 2013. ECG recordings were obtained using a single-lead electrocardiograph device (CAMUSB 6.5, Cardiosoft v6; 500 Hz sampling rate) at rest (15 seconds), during a graded 6-minute submaximal cycle ergometry test (eBike, Firmware), and post-exercise recovery (1-minute).

Exercise workload was assigned based on one’s age, sex, weight, height, and resting heart rate^23^. According to protocol, participants were instructed to perform the test with a predefined target power at 35% or 50% of the maximum predicted workload following a baseline health assessment^23^. Individuals not selected for the exercise protocol or with incomplete, corrupted, or missing ECG data were excluded from the study.

### ECG Analysis

We analysed ECG recordings according to the framework established by Sörnmo et al. in 2005^24^. ECGs were first processed to identify and exclude baseline wander and high-frequency noise, following the pipeline described by Zheng et al^25^. Detailed implementation, including signal pre-processing and quality control procedures, are provided in Supplementary Note 1.

### Conventional ECG parameters

We assessed 12 ECG features encompassing morphology and rhythm, measured during 15 seconds of rest, the final 15 seconds of exercise, and the final 15 seconds of recovery. Parameters were selected based on prior associations with CVD incidence^7,8^, mortality, and risk factors such as atrial fibrillation^26,27^. Morphological features—including PR, QRS, and QTc intervals—were extracted from a signal-averaged beat within each 15-second window. QTc was corrected using Bazett’s formula^28^ at rest and Fridericia’s^29^ during exercise and recovery. Further details are provided in the Supplementary Methods (pp. 1–2).

Rhythm features were selected based on previously validated methods by Lake and Moorman et al. 2011^26^, and Sarkar et al. 2008^27^ and included eight established markers associated with incident CVD. These comprised the number of premature ventricular contractions (PVCs), coefficient sample entropy (COSEn), and six features derived from the Poincaré plot of RR intervals over time, defined in Supplementary Figure 1. These include: (i) irregularity evidence, which quantifies the number of beats that deviate from normal sinus rhythm; (ii) regularity evidence, which measures the variation in RR intervals between windows of 3 consecutive beats and the median RR interval; (iii) density evidence, which captures how closely clustered changes in RR interval are over time; (iv) anisotropy evidence, which assesses the directional consistency of RR interval changes over time; (v) AF evidence, a composite measure that estimates the presence and severity of atrial fibrillation by balancing irregularity against the number of normal and premature beats and (vi) Evidence of Atrial Tachycardia, a composite measure that estimates the presence and severity of Atrial Tachycardia based on the regularity and distribution of heartbeats on a Poincaré distribution^27^. A full description of the candidate features and implementation details is provided in Supplementary Note 1.1.2.

### Generation of AI ECG risk score (ECG_AI_)

We developed a deep learning ECG risk model using complete 7-minute 15-second ECG recordings, comprising measurements taken during rest (15 s), exercise (6 min), and recovery (1 min), segmented into 15-second windows to match the duration of the initial resting stage of the submaximal exercise test. We used an adapted ResNet-V2 with 50 layers^30^ and 1D convolutions, combined with a long short-term memory (LSTM) network to generate a single continuous ECG risk score (ECG_AI_) for each participant. To model time-to-event outcomes, we employed a deep survival framework. In contrast to traditional deep learning approaches, deep survival models account for both time to outcome and censoring by explicitly weighting censored cases in the loss function^31^. Further implementation details are provided in Supplementary Note 1.1.3. 10-year risk estimates were derived by integrating ECG_AI_ scores with the baseline survival function evaluated up to 12.5 years using Breslow’s method^32^. A schematic overview of the ECG risk score development process is shown in Figure 1.

**Figure 1:**
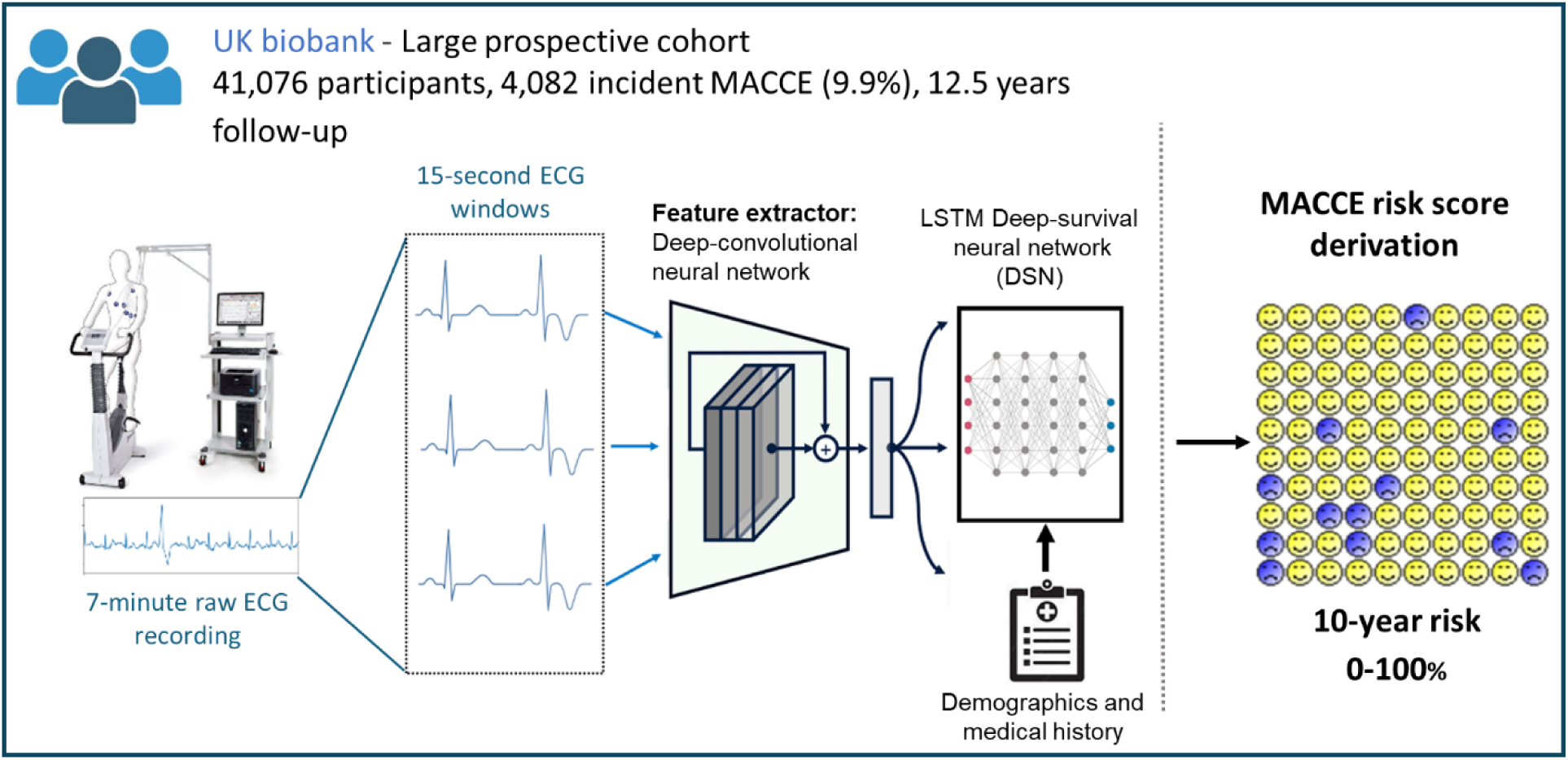
Overview of the deep survival ECG risk score derived from 7-minute 15-second submaximal exercise ECG recordings, collected from 41,076 participants (4,082 MACCE events; 9.9%). Abbreviations: MACCE = Major Adverse Cardiovascular & Cerebrovascular events, DSN = Deep survival network.

### Ascertainment of Major Adverse Cardiovascular & Cerebrovascular Events

The primary outcome of interest is the 10-year risk of incident Major Adverse Cardio/Cerebrovascular Events (MACCE), defined as the earliest recorded episode of hospitalisation or death due to ischaemic heart disease, coronary heart disease, heart failure or cerebrovascular disease including ischaemic stroke and transient ischaemic attack. The follow-up period was determined by the first appearance of International Classification of Diseases codes, Ninth or Tenth revisions (ICD9/10) codes in either HES or death registration data. Participants who did not experience an event were censored at their time of death or the end of the follow-up period. A full breakdown of the definition of MACCE is presented in Supplementary Tables 1-2.

### Model development and evaluation

We selected clinical risk factors from the QRISK3 risk calculator^1,33^ (Supplementary Table 3) as the baseline risk score for model comparison given its robust validation in the UK population and recommended use in clinical guidelines^2^. We mapped covariates to available data from the UKB assessment centre and hospital episode statistics, reported by the NHS information centre and central register Scotland, for participants from England, Wales, and Scotland, respectively. Risk factor definitions, along with relevant UKB field IDs and ICD 9/10 codes, are reported in Supplementary Table 1-3.

To account for documented miscalibration of QRISK3 in the UK Biobank^33^, the published QRISK3 algorithm was not applied directly. Instead, all model coefficients and the baseline survival function were re-estimated using Cox proportional hazards models^34^. This resulted in a newly derived clinical risk score based on the QRISK3 covariate set, tailored to the underlying risk structure of the study population. For clarity, this model is referred to as Cox clinical throughout the rest of the manuscript. The baseline survival function was calculated using Breslow’s method^32^ with mean-centred continuous covariates, and binary risk factors set to 0. Due to insufficient data on family history of CVD, including the age of diagnosis, this variable was excluded. Atrial fibrillation, renal failure, lupus, and antipsychotic medication were also removed due to limited power and high collinearity. Final model coefficients and baseline survival estimates are reported in Supplementary Section C (Tables 1C–4C, Appendix p.56).

We evaluated the added value of ECG recordings to three baseline risk models: age + sex, Cox clinical, and a deep survival network derived from QRISK3 risk factors^31^. This approach was informed by prior works, such as Attia et al (2021)^15^ and Steinfeldt et al. (2022)^35^, which demonstrated the effectiveness of deep learning in ECG analysis and risk prediction.

We performed internal validation using stratified nested five-fold cross-validation, withholding one-fold for testing and 10% of the training data used for validation. Following Steinfeldt et al. (2022), we evaluated model performance on an aggregated test set, using 1000 bootstrap iterations to obtain 95% confidence intervals^35^. Further details on model development and validation are provided in Supplementary Note 1.

### Statistical analysis

We calculated the follow-up time for each participant as the number of years from the initial ECG scan until the earliest date of either (i) first recorded MACCE, (ii) death not caused by CVD or (iii) the most recent publication of NHS hospital episode statistics for England: 31st October 2022; Scotland: 31^st^ August 2022; and Wales 5^th^ May 2022^36^.

We excluded all participants with known history of MACCE, in addition to those receiving calcium channel blockers or antiarrhythmic medications (Supplementary Table 4), prior to the submaximal exercise ECG test (Figure 2) as these drugs can alter heart rate response, ECG morphology, and autonomic function, potentially confounding the association between exercise ECG features and future cardiovascular risk.

**Figure 2:**
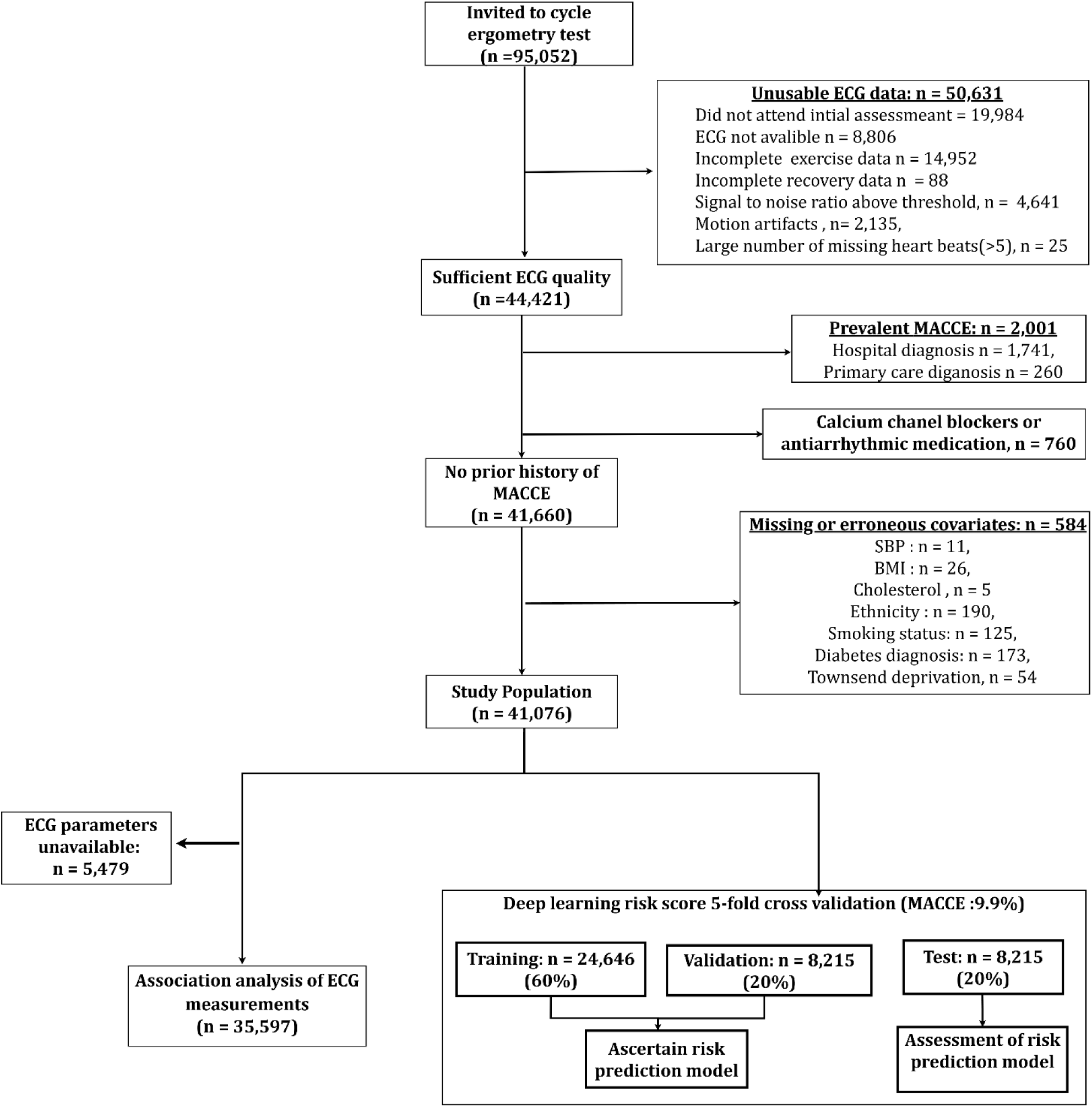
Characteristics of study population: Flowchart illustrating the number of participants included in the study, along with any exclusion criteria and derivation of development/validation sets for MACCE. Abbreviations: n = Number of participants, MACCE = Major Adverse Cardiovascular & Cerebrovascular events.

Continuous variables were standardised using z-score normalisation. Multiclass categorical variables were one-hot encoded, creating binary variables for each sub-category. We dichotomised ethnicity into white/non-white due to the heavy class imbalance (Table 1), while smoking was factorised into three categories (never, previous, current).

**Table 1:**
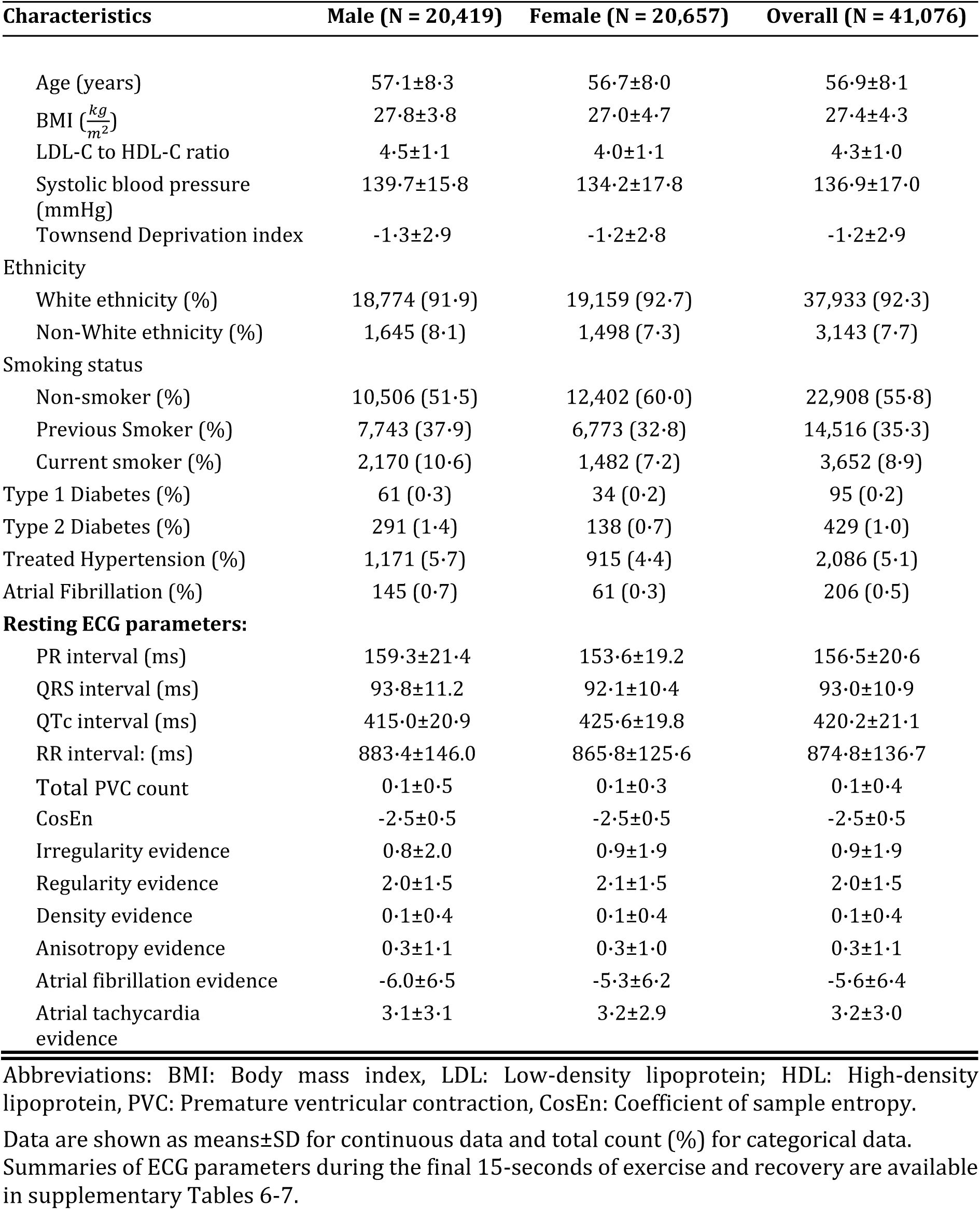
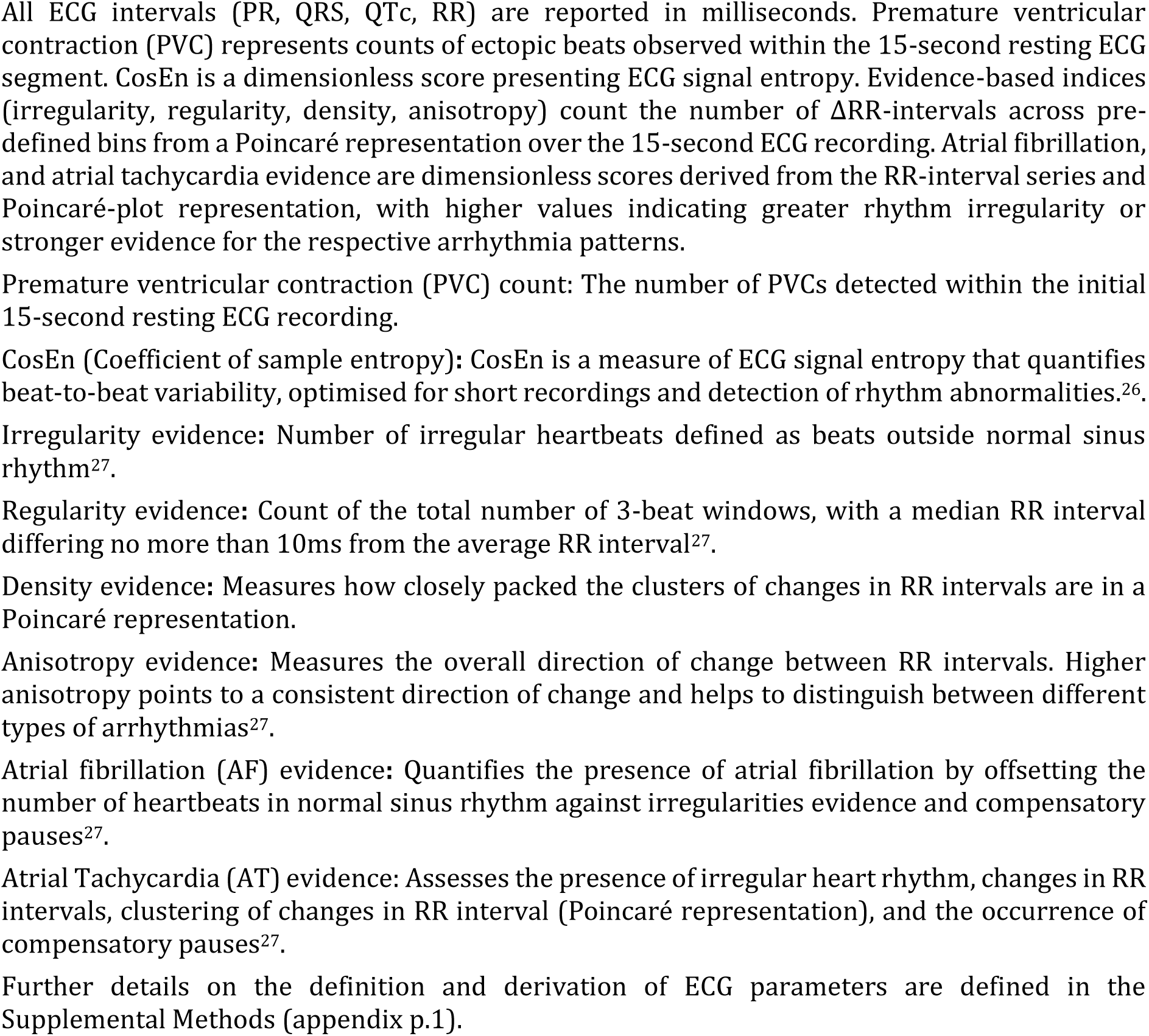
Baseline characteristics and ECG parameters of 41,076 UK Biobank participants without a history of MACCE who underwent a submaximal cycle ergometry test.

For risk factors missing in <1% of participants we applied a complete case analysis. For variables missing >1%, we performed single imputation using Bayesian Ridge regression^37^, fitted on training folds and applied to each test set independently.

Minimum sample size calculations conducted using the framework proposed by Riley et al. (2018)^38^, confirmed sufficient events to develop models with up to 71 covariates. These included QRISK3 risk factors, ECG parameters derived from rest, exercise, and recovery phases, and interactions with age modelled using polynomial terms (Supplementary Table 5).

### Independent association of ECG measurements with incident MACCE

We investigated the strength of the associations between ECG parameters and risk of MACCE using multivariable-adjusted Cox proportional hazard analysis, minimally adjusted for age, sex, smoking status, ethnicity, and deprivation, measured by Townsend’s deprivation index^39^. We modelled the continuous association between ECG_AI_ scores and incident MACCE using penalised splines with 3 degrees of freedom. To aid interpretability, ECG_AI_ was min–max scaled (0–1), and hazard ratios were expressed relative to the 20th percentile (HR = 1). Statistical significance was determined using a chi-squared test with a threshold of 0.05, adjusted for multiple testing using the Benjamini-Hochberg procedure^40^. We conducted a complete case analysis, removing any individual with missing PR, RR, QRS or QTc interval.

### Added value of ECG measurements for risk prediction

Model performance was assessed using Harrell’s C-index^41^ and the integrated calibration index (ICI)^42^. We assessed the reclassification of participants for each 10-year risk score using standardised net benefit (NB)^43^ and net reclassification index (NRI)^44^ at the currently recommended clinical threshold of 10%. P-values and z-statistics were derived via bootstrapping for 1000 iterations, with a significance threshold of 0.05^45–47^. This study complies with TRIPOD guidance on the reporting of risk prediction studies (checklist provided in Supplemental Section D)^48^.

### Sensitivity analysis

To investigate possible reverse causation bias, we repeated the Cox regression association analyses after removing MACCE that occurred within the first 2 and 5 years of follow-up.

We conducted a sensitivity analysis, individually assessing the contribution of each stage during the submaximal exercise test to MACCE risk prediction. We developed independent risk scores for the 15-second resting period, the final 15 seconds of exercise, and the final 15 seconds of recovery, respectively.

We subsequently evaluated the incremental value of conventional ECG parameters identified as significantly associated with MACCE in prior association analyses. These parameters were incorporated into each baseline risk score in the subset of participants with available conventional ECG data.

## Results

Following exclusion for reasons highlighted in Figure 2, 41,076 individuals without prior history of MACCE and with complete ECG recordings were included in the study. The study population consisted of a balanced set of middle-aged men and women (50.3% female, mean age 56.9 ± 8.1 years, Table 1). After a median follow-up period of 12.5 years (IQR: 0.26), 4,082 (9.9%) individuals were diagnosed with MACCE (3,392 non-fatal and 690 fatal). Conventional ECG parameters were available in a subset of 35,597 individuals and are summarised in Table 1 and Supplementary Tables 6-7. In this subgroup, there were 1,798 (9.9%) and 793 (4.6%) MACCE events, for men and women, respectively. We found no statistically significant difference in MACCE incidence between this subgroup and the full cohort (Chi-squared statistic = 0.92; p-value = 0.34); incident MACCE = 3,463 (9.7%); median follow-up = 12.5 (IQR 0.27) years.

### Association of ECG risk scores with MACCE Outcomes

We first evaluated the independent association of our deep learning-derived ECG risk score (ECG_AI_), with incident MACCE over 12.5 years of follow-up, alongside conventional ECG parameters measured during rest, exercise, and recovery in participants with available data. We observed that ECG_AI_ risk scores derived from complete 7-minute 15-second submaximal exercise ECG recordings, had a linear association with incident MACCE (adjusted hazard ratio [HR]: 1.18 per SD increase, 95% CI:1.15-1.21, p < 0.001, Figure 3). We observed higher risk of MACCE for people in the two highest ECG_AI_ quintiles relative to those in the bottom ECG_AI_ quintile (HR: 1.20, 95% CI: 1.07-1.34; and HR: 1.31, 95% CI: 1.18-1.47; respectively). We found that ECG_AI_ scores and conventional ECG parameters derived independently from rest, exercise, and recovery were also positively associated with incident MACCE (Figure 4).

**Figure 3:**
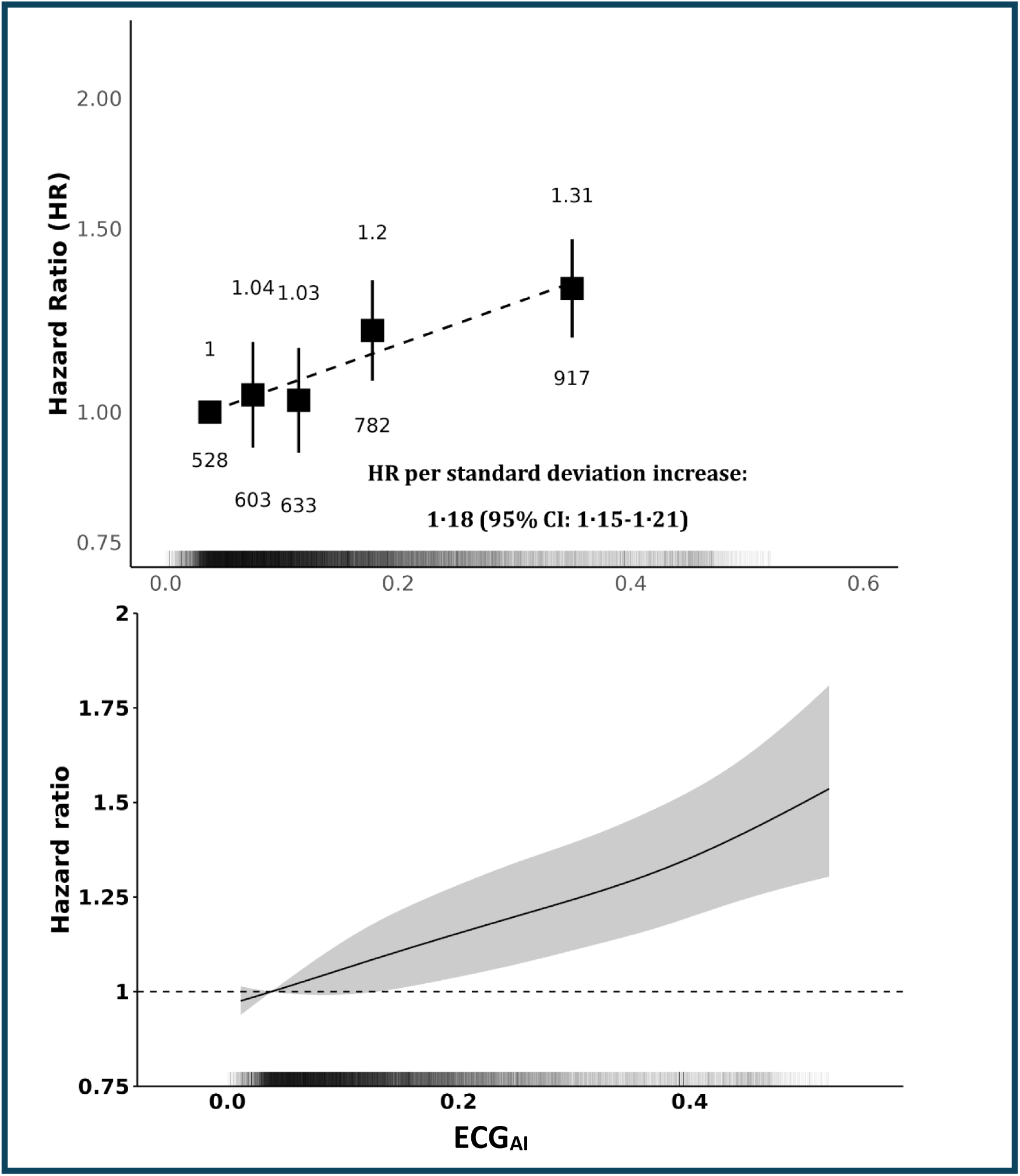
Association between the output of a deep survival network trained on 7-minute 15-second ECG recordings (ECG_AI_), during a submaximal cycle ergometry exercise test, and the risk of incident Major Adverse Cardiovascular and Cerebrovascular Events (MACCE) over 12.5 years of follow-up. Associations were estimated using Cox proportional hazards models with penalised splines (3 degrees of freedom), with adjustment for age, sex, smoking status, ethnicity, and Townsend deprivation index. (Top) Forest plots showing the associations between MACCE and quintiles of the deep learning–derived ECG_AI_ risk score; (Bottom) the continuous ECG_AI_ score with hazard ratios expressed relative to the 20th percentile (HR = 1), among 35,597 UK Biobank participants with valid ECG parameters, and 3,463 (9.7%) reported MACCE. In the top figure, the hazard ratio and number of events are plotted above and below each data point, respectively. The bottom figure shows a spline plot of the hazard ratio and 95% confidence interval for the association between ECG_AI_ risk scores and MACCE. The grayscale bar at the bottom represents the distribution of ECG_AI_ values across the study population. Darker regions indicate a higher density of participants with corresponding ECG_AI_ values, while lighter regions indicate lower density.

**Figure 4:**
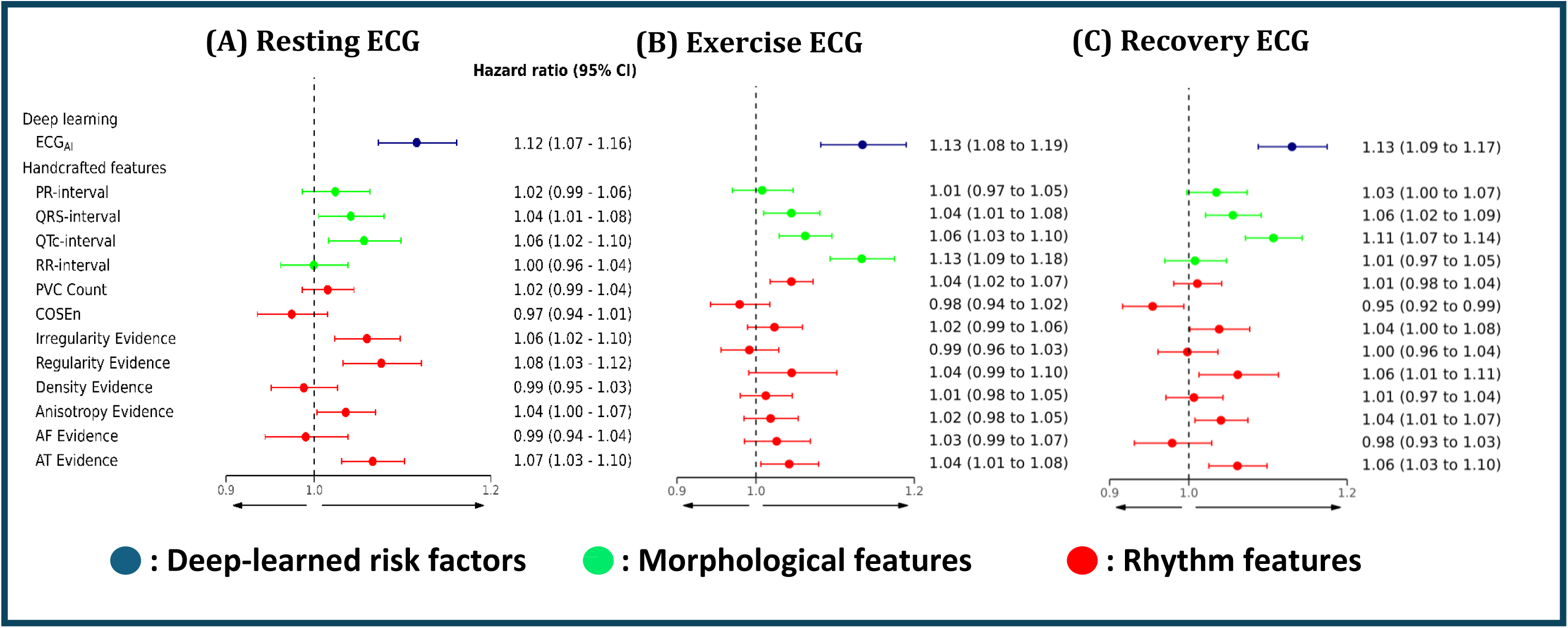
Association of deep-learned ECG risk scores (ECG_AI_) and conventional ECG parameters obtained during (A) 15 seconds of rest, (B) the final 15 seconds of a 6-min period of submaximal exercise, (C) the final 15 seconds of a 1-min recovery period post exercise with the 10-year risk of MACCE. Each ECG parameter was analysed in a separate Cox model, adjusted for age, smoking status, Townsend deprivation index^39^, and ethnicity. Irregularity, Regularity, Anisotropy, and AF evidence were adjusted for the number of beats in the signal to account for higher absolute values in participants with faster heart rates. Hazard ratios are reported per standard deviation increase. Cox proportional hazard models were developed on the aggregated test sets across five cross-validation folds, consisting of 35,597 participants and 3,463 (9.7%) MACCE. The ECG parameters were selected based on previously reported associations with CVD mortality^7,8^, incidence, common risk factors, and other conditions, including chronic kidney disease^9^ and atrial fibrillation^26,27^. We also considered rhythm features that have previously demonstrated strong predictive performance for CVD ^20,21^.

### Predictive performance of submaximal exercise ECG recordings

Next, we assessed the predictive performance of univariate ECG measurements for the 10-year risk of incident MACCE, among participants with valid conventional ECG parameters. After 10 years of follow-up, we report 2,591 (7.3%) MACCE.

In models excluding clinical risk factors, ECG_AI_ demonstrated positive predictive performance for incident MACCE (Table 2), with exercise ECG measurements yielding the strongest predictive performance. ECG_AI_ derived from exercise ECG measurements achieved a C-index of 0.591 (95% CI 0.583 - 0.599) and a net benefit of 0.044 (0.029 - 0.058), at a risk threshold of 10%. Across all phases of the submaximal exercise test, ECG_AI_ outperformed conventional ECG parameters and individual QRISK3 risk factors other than age and systolic blood pressure (Supplementary Table 8). Continuous calibration curves of univariate ECG_AI_ risk scores demonstrated systematic underprediction in individuals at lower observed risk of MACCE (<5%); however, they were generally well-calibrated for individuals with observed risks > 5% (Supplementary Figure 2).

**Table 2:** Comparative performance of clinical and univariate ECG_AI_ risks scores for the prediction of Major Adverse Cardiovascular & Cerebrovascular risk.

| Model |  | Hazard Ratio<br>(95% CI) | C-index<br>(95% CI) | Net benefit<br>(95% CI) |
| --- | --- | --- | --- | --- |
| Age |  | 1.12 (1.11 - 1.13) | 0.66(0.66 - 0.67) | 0.18(0.16 - 0.20) |
| Cox clinical |  | 1.06 (1.06 - 1.08) | 0.67 (0.67 - 0.68) | 0.22 (0.20 - 0.24) |
| Clinical DSN |  | 1.09 (1.07 - 1.12) | 0.70 (0.70 - 0.71) | 0.21 (0.18 - 0.24) |
| ECG<sub>AI</sub> | Rest | 1.08 (1.07 - 1.12) | 0.57 (0.56 - 0.58) | 0.03 (0.02 - 0.04) |
|  | Exercise | 1.06 (1.05 - 1.11) | 0.59 (0.58 - 0.60) | 0.04 (0.03 - 0.06) |
|  | Recovery | 1.07 (1.06 - 1.13) | 0.57 (0.56 - 0.58) | 0.05 (0.03 - 0.06) |
|  | Complete ECG | 1.11 (1.09 - 1.12) | 0.57 (0.56 - 0.58) | 0.03 (0.02 - 0.04) |

Abbreviations: CI = Confidence interval, DSN = Deep survival network.
Data are reported on 35,597 participants with valid ECG parameters, and 2,591 (7.27%) Major Adverse Cardiovascular & Cerebrovascular events (MACCE) were reported within 10 years. 95% confidence intervals were obtained via 1000 bootstrap iterations.
Each row corresponds to a separate Cox model in which the model was fitted independently with the highlighted risk factor.
ECG<sub>AI</sub> scores were derived from a deep survival network using 7-minute 15-second ECG recording during a submaximal exercise cycle ergometry test.
Hazard ratios are derived from separate Cox proportional hazards models, unadjusted for other risk factors.
Unadjusted hazard ratios are reported per unit increase, while net benefit is reported at a 10% risk threshold.
Clinical DSN: Age, BMI, SBP, SBP5, Diabetes type 1, Diabetes type 2, HDL ratio, Townsend deprivation index, Smoker former, Smoker current, Smoker never, Ethnic Background, Corticosteroid, Migraine, Hypertension, Severe mental illness.
Fractional polynomials: Age FP1 (males) = $(age/10)^{-1}$ , Age FP2 (males) = $(age/10)^3$ , BMI FP 1 (males) = $(BMI/10)^{-2}$ , BMI FP 2 (males) = $(BMI/10)^{-2} \times \log(BMI/10)$ , Age FP1 (Females) = $age/10^{-2}$ , Age FP2 (Females) = $age/10$ , BMI FP 1 (Females) = $(BMI/10)^{-2}$ , BMI FP 2 (Females) = $(BMI/10)^{-2} \times \log(BMI/10)$ .

### Added value of Submaximal ECG measurements to clinical risk scores

To evaluate the added value of ECG parameters for 10-year prediction of incident MACCE, we assessed the performance of ECG_AI_ when added to three baseline models: age + sex, Cox clinical, and clinical deep survival networks, across the full study cohort (n = 41,076), among whom 3,053 MACCE occurred after 10 years of follow-up. We subsequently compared each risk score to our clinical deep survival network + ECG model, derived from QRISK3 risk factors and complete 7-minute 15-second ECG recordings.

We found all models were well-calibrated within the study population. Supplementary Figure 3-5 shows the continuous calibration curves for each risk score before and after the addition of ECG measurements. Age + sex models yielded C-indices of 0.664 (95% CI: 0.654–0.673) in men and 0.669 (0.657–0.684) in women. Adding ECG_AI_ modestly improved discrimination (men: ΔC-index 0.008, Net Reclassification Index (NRI) 0.169; women: ΔC-index 0.005, NRI 0.163; all p < 0.001; Figure 5, Table 3).

**Figure 5:**
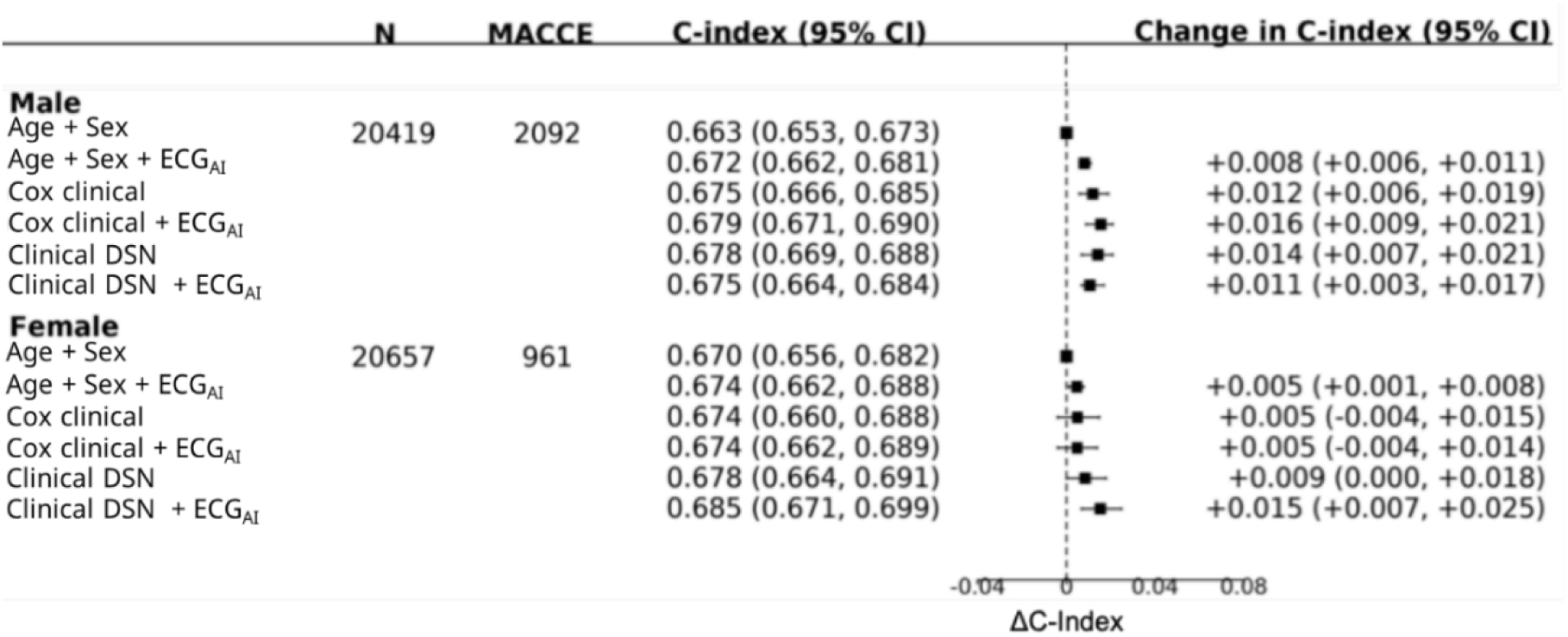
Forest Plot showing Harrell’s C-index for age + sex, Cox clinical, and clinical Deep Survival Network (DSN) risks scores before and after the addition of ECG data from a submaximal exercise cycle ergometry test (7-minute 15-second recordings) in 41,076 UK Biobank participants, stratified by sex. Abbreviations: N = Number of participants, MACCE = Major Adverse Cardiovascular & Cerebrovascular events, DSN = Deep survival network, CI = Confidence interval. Squares represent change in C-index compared to age and horizontal lines are 95% confidence intervals generated using 1,000 bootstrap samples. 95 % CIs were derived via bootstrapping for 1000 iterations: significance threshold = 0.05. Data are reported on 41,076 participants with 3,053 (7.4%) Major Adverse Cardiovascular & Cerebrovascular events (MACCE) observed over 10 years of follow-up. Cox clinical: Age FP1 + Age FP2 + BMI FP1 + BMI FP2 + SBP + SBP5 + Diabetes type 1 + Diabetes type 2 + HDL ratio + Townsend deprivation index + Smoker former + Smoker current + Smoker never + Ethnic Background + Corticosteroid + Migraine + Hypertension + Severe mental illness + Age FP1 × Townsend deprivation index + Age FP2 × Townsend deprivation index + Age FP1 × BMI FP 1 + Age FP1 × BMI FP 2 + Age FP1 × BP + Age FP1 × Diabetes type 1 + Age FP1 × Diabetes type 2 + Age FP1 × hypertension + Age FP2 × BMI FP1 + Age FP2 × BMI FP2 + Age FP2 × SBP + Age FP2 × Diabetes type1 + Age FP2 × Diabetes type2 + Age FP2 × Hypertension. Fractional polynomials: *Age FP1 (males) =* (𝑎𝑔𝑒/10)^−1^*, Age FP2 (males) = (age/10)*^3^*, BMI FP 1 (males) =* (𝐵𝑀𝐼/10)^−2^*, BMI FP 2 (males) =* (𝐵𝑀𝐼/10)^−2^ 𝑋 𝑙𝑜𝑔(𝐵𝑀𝐼/10)*, Age FP1 (Females) =* 𝑎𝑔𝑒/10^−2^*, Age FP2 (Females) = age/10, BMI FP 1 (Females) =* (𝐵𝑀𝐼/10)^−2^*, BMI FP 2 (Females) =* (𝐵𝑀𝐼/10)^−2^ 𝑋 𝑙𝑜𝑔 (𝐵𝑀𝐼/10). Clinical DSN: Age, BMI, SBP, SBP5, Diabetes type 1, Diabetes type 2, HDL ratio, Townsend deprivation index, Smoker former, Smoker current, Smoker never, Ethnic Background, Corticosteroid, Migraine, Hypertension, Severe mental illness. ECG_AI_ scores were derived from a deep survival network using 7-minute 15-second ECG recording during a submaximal exercise cycle ergometry test.

**Table 3:** Added value of ECGAI for the prediction of Major Adverse Cardiovascular & Cerebrovascular risk in 41,076 UK Biobank participants.

| Model | Comparator | Net benefit<br>(95% CI) | Categorical NRI<br>(95% CI) | Continuous NRI<br>(95% CI) |
| --- | --- | --- | --- | --- |
| Age + Sex | Age + Sex + ECG<sub>AI</sub> | **0.01 (0.0 - 0.02) | 0.01 (0.00 - 0.02) | **0.144 (0.11 - 0.18) |
| Cox clinical | Cox clinical + ECG<sub>AI</sub> | 0.00 (-0.01 - 0.01) | 0.00 (-0.007 - 0.007) | **0.097 (0.061 - 0.13) |
| Clinical DSN | Clinical DSN + ECG<sub>AI</sub> | 0.01 (-0.01 - 0.02) | *0.01 (0.00 - 0.03) | **0.27 (0.24 - 0.31) |

Abbreviations: NRI = Net reclassification index, DSN = Deep survival network, CI = Confidence interval.
Data are reported on 41,076 participants with 3,053 (7.4%) Major Adverse Cardiovascular & Cerebrovascular events (MACCE) over 10 years of follow-up.
P-values and 95% confidence intervals were derived via bootstrapping for 1000 iterations, with a significance threshold of 0.05.
\*p value < 0.05
\*\*p value < 0.001
Clinical DSN: Age, BMI, SBP, SBP5, Diabetes type 1, Diabetes type 2, HDL ratio, Townsend deprivation index, Smoker former, Smoker current, Smoker never, Ethnic Background, Corticosteroid, Migraine, Hypertension, Severe mental illness.
ECG<sub>AI</sub> scores were derived from a deep survival network using 7-minute 15-second ECG recording during a submaximal exercise cycle ergometry test.

Cox clinical showed strong discrimination, with a C-index of 0.675 (95% CI: 0.666–0.685) in men and 0.674 (0.660–0.688) in women. In the overall population adding ECG_AI_ to cox clinical risk scores was associated with a modest improvement in discrimination (ΔC-index 0.002, 95% CI 0.000–0.003; p=0.002) and a continuous NRI of 0.097 (95% CI 0.061–0.133; p<0.001). In men, C-index increased by 0.003 (95% CI: 0.002–0.005), with a continuous NRI of 0.108 (95% CI: 0.063–0.152). For women, we found similar gains in continuous NRI (0.093; 95% CI: 0.029–0.157), but no significant change in C-index. No improvements were observed in categorical NRI or net benefit at the 10% risk threshold (Table 3, Supplementary Table 9).

### Added value of ECG to clinical deep neural networks

To determine if neural networks improve risk prediction over conventional models, we compared a deep-survival neural network, trained using QRISK3 risk factors (clinical deep survival network) against Cox clinical. We observed a significant improvement in model discrimination over Cox clinical (ΔC-index = 0.032, 95% CI: 0.026, 0.038, p < 0.001) (Figure 5). However, no significant changes in reclassification at the recommended intervention threshold of 10% were reported (Supplementary Table 10).

Adding ECG_AI_ to the clinical deep survival network further improved risk stratification over the baseline model, with modest but statistically significant improvements in continuous and categorical NRI at a risk threshold of 10% (Table 3). In contrast to Cox clinical, we observed significant improvements in model discrimination for women (ΔC-index = 0.007; 95% CI: 0.002–0.011; p < 0.001), while no significant change was reported among men.

When comparing the clinical deep survival network + ECG risk scores to Cox clinical, we observed a significant improvement in continuous NRI for men and women alike (Table 4, Supplementary Table 11). At a 10% risk threshold, we report an increase in categorical NRI among women of 0.085 (95% CI: 0.061–0.108; p < 0.001) compared to Cox clinical alone. No improvement was observed in men or in the combined population (Table 4).

**Table 4:** Performance of clinical deep survival networks (DSN), following the addition of raw ECG signals recorded during a 7-minute 15 second submaximal exercise test, to assess the 10-year risk of Major Adverse Cardiovascular & Cerebrovascular Disease

| Model | Comparator | Net benefit<br>(95% CI) | Categorical NRI<br>(95% CI) | Continuous NRI<br>(95% CI) |
| --- | --- | --- | --- | --- |
| Age + Sex | Age + Sex (DSN) + ECG<sub>AI</sub> | 0.01 (-0.01 - 0.02) | *0.013 (0.00 - 0.02) | **0.13 (0.09 - 0.16) |
| Cox clinical | Clinical DSN + ECG<sub>AI</sub> | 0.00 (-0.02 - 0.01) | 0.01 (-0.01 - 0.02) | **0.23 (0.19 - 0.27) |

Abbreviations: CI = Confidence interval; DSN = Deep survival network, NRI = Net reclassification index.
Data are reported on 41,076 participants with 3,053 (7.4%) Major Adverse Cardiovascular & Cerebrovascular events (MACCE) observed over 10 years of follow-up.
p-values were derived via bootstrapping for 1000 iterations, with a significance threshold of 0.05.
\*p value < 0.05
\*\*p value < 0.001
Clinical DSN: Age, BMI, SBP, SBP5, Diabetes type 1, Diabetes type 2, HDL ratio, Townsend deprivation index, Smoker former, Smoker current, Smoker never, Ethnic Background, Corticosteroid, Migraine, Hypertension, Severe mental illness.
Fractional polynomials: Age FP1 (males) = $(age/10)^{-1}$ , Age FP2 (males) = $(age/10)^3$ , BMI FP 1 (males) = $(BMI/10)^{-2}$ , BMI FP 2 (males) = $(BMI/10)^{-2} \times \log(BMI/10)$ , Age FP1 (Females) = $age/10^{-2}$ , Age FP2 (Females) = $age/10$ , BMI FP 1 (Females) = $(BMI/10)^{-2}$ , BMI FP 2 (Females) = $(BMI/10)^{-2} \times \log(BMI/10)$ .
ECG<sub>AI</sub> scores were derived from a deep survival network using 7-minute 15-second ECG recording during a submaximal exercise cycle ergometry test.

### Subgroup and sensitivity analysis

During the study period, 2,722 men and 1,360 women experienced a MACCE. ECG_AI_ risk scores showed a similarly strong association with MACCE across sexes for all stages of the submaximal exercise test. In contrast, conventional ECG parameters demonstrated stronger associations in men, and greater uncertainty among women (Supplementary Figures 6–8).

Excluding cases within the first 2 years (n = 347) and 5 years (n = 1,087) did not materially change the hazard ratios for ECG_AI_ or conventional parameters (Supplementary Tables 12–13).

Stratifying across age and sex (Supplementary Tables 14–17) we found that clinical deep survival network + ECG_AI_ yielded the greatest improvements in reclassification in younger men (<45 years of age) and older women (>55 years of age). Among men aged <45 (n = 2,476), categorical NRI rose by 1.1% (95% CI: 0.5–1.7%) and net benefit by 4.8% (95% CI: 2.3–7.7%). For women aged 55–65 (n = 8,739), categorical NRI increased by 3.6% (95% CI: 0.9–6.4%), and in those >65 years of age(n = 3,135), by 9.5% (95% CI: 4.2–14.8%), with a 6.8% increase in net benefit (95% CI: 1.0–12.5%).

We also assessed the added value of ECG_AI_ scores derived from 15-second recordings during rest, exercise, and recovery to each baseline risk model (Supplementary Tables 18-23; Figures 9–11). When added separately to each baseline model, ECG_AI_ derived from resting and exercise ECG data was associated with improvements in the continuous NRI; however, no change was observed in categorical NRI evaluated at a 10% risk threshold (Supplementary Tables 22–23).

### Conventional ECG parameters

As a secondary analysis, we evaluated the added value of conventional ECG parameters to each baseline risk model in 35,596 individuals with available ECG parameters across rest, exercise, and recovery. We selected ECG parameters that were significantly associated with MACCE during the previous association analysis.

ECG_AI_ did not significantly outperform multivariable models using conventional ECG parameters in either of the Cox or deep survival models (Supplementary Table 24). Although conventional parameters performed better than ECG_AI_ in models excluding clinical risk factors, adding them to Cox clinical or our clinical deep survival network provided no significant improvement in discrimination or reclassification (Supplementary Tables 25–26).

## Discussion

In this study we present ECG_AI_, a novel deep neural network–based risk score for estimating long-term risk of major adverse cardiovascular and cerebrovascular events (MACCE). To our knowledge, this is the largest population-based prospective cohort study investigating the added value of submaximal exercise ECG recordings to existing clinical risk scores in individuals with no history of CVD. The main findings of our investigation are: (i) ECG measurements during rest, exercise, and recovery are independently associated with the 10-year risk of incident MACCE; (ii) ECG measurements recorded from a submaximal exercise test are predictive of MACCE in the absence of clinical risk factors; (iii) ECG measurements from a risk-stratified submaximal exercise test had minimal impact on reclassifying patients under current treatment guidelines.

This study builds on prior research investigating the application of deep learning to ECG data for cardiovascular risk prediction. Hughes et al. (2023) and Sau et al. (2024) independently demonstrated that deep-learning-derived ECG risk scores can improve predictive performance beyond established clinical risk factors^49,50^. Our study differs from these prior investigations in several important ways. First, instead of relying on clinical 12-lead resting ECGs, we analysed single-lead ECG recordings acquired during a submaximal exercise test, independently evaluating the prognostic value of ECG features across rest, exercise, and recovery phases. In doing so, we address a key gap identified by the most recent United States Preventive Services Task Force (USPSTF) review, citing a lack of evidence on the prognostic value of exercise ECG^10^. Additionally, by assessing the incremental value of ECG features when added to the established clinical risk factors used in QRISK3, the risk tool currently adopted in UK primary care, we demonstrate the additive benefit of ECG-derived metrics beyond conventional risk stratification in populations without prior CVD. Finally, in contrast to prior studies we evaluated model calibration and reclassification, providing a more comprehensive evaluation of clinical utility, adhering to TRIPOD and USPSTF recommendations^10,48^.

Through development and internal validation, we demonstrate that deep ECG_AI_ risk scores yield modest predictive performance in identifying individuals at high risk of MACCE without additional clinical information. This aligns with prior observational studies on deep learning–based ECG risk models derived using 12-lead resting ECG measurements, which reported C-indices of 0.55-0.64^49,50^. Adding ECG_AI_ to established clinical risk scores resulted in a modest improvement in discrimination. However, improvements in reclassification were minimal, as assessed by net reclassification improvement and net benefit analysis. These findings are consistent with the conclusion of the USPSTF that while resting and exercise ECGs may improve absolute predictive performance; they provide limited benefit in identifying additional high-risk individuals for primary prevention^10^. Possible explanations for the modest predictive performance of submaximal exercise ECGs may include both methodological and physiological factors. For example, single-lead exercise recordings are subject to lower signal quality due to noise, motion artefacts and reduced spatial resolution compared to standard 12-lead resting ECGs, which may obscure subtle ECG features including signs of ischaemia or repolarisation abnormalities^51^. Furthermore, most participants achieved only 50% of their estimated maximum workload (submaximal exercise), which may not elicit sufficient cardiovascular stress to unmask latent electrophysiological or ischemic abnormalities. Finally, pre-screening for exercise suitability introduced additional selection within the already low-risk UK Biobank population^20^, yielding a healthier subgroup than both the broader cohort and prior outpatient populations. This enhanced selection likely contributed to the lower observed incidence of CVD events and may have attenuated the positive predictive value of the ECG-derived risk scores.

### Strengths & Limitations

Our study has several strengths and limitations. Key strengths include its large sample size of 41,076 individuals without prior cardiovascular disease and novel comparison of deep-learning risk scores derived from ECGs recorded during rest, submaximal exercise, and in post-exercise recovery, an approach not undertaken in prior large-scale studies using deep-learning, which have been limited to standard 12-lead resting ECGs^49,50^. Finally, following USPSTF and NICE recommendations, we quantified the incremental value of ECG derived risk scores to clinical risk factors from established risk models such as QRISK3, by evaluating not only discrimination but also calibration and reclassification, providing a more complete view of predictive performance and clinical utility^2,10^.

Nevertheless, this study is subject to several limitations. First, the UK Biobank is subject to a well-documented healthy volunteer selection bias^20^, further compounded in our study by the strict eligibility criteria for the submaximal exercise test, with participants exhibiting reduced risk of CVD compared to the general population (7.3% versus ∼18% in the general population)^52^. As a result, current clinical risk thresholds may be suboptimal within the study population, potentially affecting reclassification and net benefit analyses. Second, motion artefacts and noise introduced during exercise limited the reliability of ECG morphology measurements across the full cohort, leading to the exclusion of a subset of participants from association analyses and risk score development using conventional ECG parameters. Finally, we could not externally validate these results due to a lack of access to a comparable dataset, limiting the generalisability of our findings beyond the UK Biobank.

## Conclusion

This study found that although submaximal exercise ECG measurements were independently associated with incident MACCE, they provided minimal incremental value to existing clinical risk scores. While deep learning risk scores integrating clinical risk factors and submaximal ECG measurements improved discrimination compared with Cox clinical and enhanced risk reclassification in women, further validation in external and more diverse populations is required. Our findings align with the conclusions of the USPSTF, showing limited evidence that exercise ECGs improve clinical risk prediction in low-risk populations and reinforce the limited utility of routine ECG screening for population-level cardiovascular prevention.

### Research in context Evidence before this study

We conducted a search in Google Scholar and PubMed databases from the beginning of available records up to 16th October 2025, with no language restrictions. Our search terms consisted of “ECG/electrocardiography”, “Exercise ECG”, “ECG AND machine learning”, “ECG AND Deep learning”, “ECG and MACE”, “ECG AND MACCE”, “Survival analysis”. Existing studies have shown modest improvements in cardiovascular risk prediction when resting or exercise ECG is added to clinical risk factors. However, they primarily focused on a narrow set of hand-selected ECG features, which often lack sensitivity to subtle abnormalities and capture only a fraction of the raw signal. In contrast, deep learning of raw ECG signals has demonstrated state-of-the-art performance in ECG analysis, often outperforming conventional markers in predicting cardiovascular outcomes. To date, no study has investigated the integration of submaximal exercise ECG measurements with clinical risk scores using deep neural networks.

### Added value of this study

To our knowledge, this is the largest population-based cohort study to evaluate the added value of exercise ECG beyond clinical risk factors derived from QRISK3, the risk model currently used in UK primary care, in individuals without a history of major adverse cardiovascular and cerebrovascular events (MACCE), and the first to apply deep survival models in this context. We found that ECG signals measured during rest, exercise and post-exercise recovery were associated with incident MACCE, independent of established clinical risk factors. Moreover, deep learning models trained on complete submaximal ECG recordings provided modest improvements in the discrimination of established clinical risk scores. However, ECG measurements did not meaningfully reclassify individuals across treatment thresholds defined by current clinical guidelines.

### Implications of all the available evidence

Current evidence suggests that ECG measurements are strongly associated with the risk of cardiovascular disease and can enhance prediction when combined with established clinical risk factors. However, there is little evidence on whether exercise ECG recordings can improve risk stratification based on current clinically recommended risk thresholds for statin treatments. Our findings support current guidelines advising against the use of exercise ECG for low-risk individuals. Our study motivates further research into the utility of deep learning models in higher-risk populations, where the potential benefits of incorporating exercise ECG data may be more pronounced.

## Data Availability

UK Biobank data are available to bona fide researchers on application at

https://www.ukbiobank.ac.uk/enable-your-research/apply-for-access.

## Authorship Contributions

AD: conceptualisation, data curation, formal analysis, methodology, supervision, writing - review & editing. AS: conceptualisation, data curation, formal analysis, methodology, validation, writing - review & editing. BC: conceptualisation, methodology, supervision, writing - review & editing. SVD: data curation, formal analysis, methodology, writing - review & editing.

## Data sharing

UK Biobank data are available to bona fide researchers on application at https://www.ukbiobank.ac.uk/enable-your-research/apply-for-access.

## Code availability

All risk score derivation and analysis scripts are freely available for academic use on GitHub: https://github.com/OxWearables/.

## Acknowledgements

We would like to thank the UK Biobank team and participants, without whom this research would not be possible.

## Funder Acknowledgements

This research was conducted using data from UK Biobank, a major biomedical database, via application number 59070. This project was supported by the Engineering and Physical Sciences Research Council (EPSRC) Centre for Doctoral Training in Health Data Science (EP/S02428X/1). AD’s research team is supported by a range of grants from the Wellcome Trust [223100/Z/21/Z, 227093/Z/23/Z], Novo Nordisk, Swiss Re, Boehringer Ingelheim, National Institutes of Health’s Oxford Cambridge Scholars Program, British Heart Foundation Centre of Research Excellence (RE/18/3/34214), and funding administered by the Danish National Research Foundation in support of the Pioneer Centre for SMARTbiomed. For the purpose of open access, the author(s) has applied a Creative Commons Attribution (CC BY) license to any Author Accepted Manuscript version arising.

