## Supplementary for "Development and validation of an ECG-based 10-year risk prediction model for Major Adverse Cardiac and Cerebrovascular Events in UK Biobank"

Supplement to: Adam Sturge, Dr Stefan van Duijvenboden, Prof. Barbara Casadei, Prof. Aiden Doherty

#### Supplemental Materials

Supplementary Table 10 ... 38
Supplementary Table 11 ... 39
Supplementary Table 12 ... 40
Supplementary Table 13 ... 41
Supplementary Table 14: ... 42
Supplementary Table 15 ... 43
Supplementary Table 16 ... 44
Supplementary Table 17 ... 45
Supplementary Table 18 ... 46
Supplementary Table 19 ... 47
Supplementary Table 20 ... 48
Supplementary Table 21 ... 49
Supplementary Table 22 ... 51
Supplementary Table 23 ... 52
Supplementary Table 24 ... 53
Supplementary Table 25 ... 54
Supplementary Table 26 ... 55
**Section C: Model coefficients, formulas and usage ... 56**
Table 1C Cox clinical ... 56
Table 2C Cox clinical + $ECG_{AI}$ ... 58
Table 3C Cox clinical in sub cohort of 35,597 with available conventional ECG parameters ... 61
Table 4C Cox clinical + Conventional ECG parameters ... 64
**Section D: TRIPOD guidelines for predictive model development ... 70**

### Section A

### Supplemental Note 1: Model development

#### 1.1 Development of Electrocardiogram risk scores for the 10-year risk prediction of Major Adverse Cardiovascular and Cerebrovascular Events

##### 1.1.1 ECG pre-processing

Recorded ECGs were stored in XML format without prior filtering or post-processing. To mitigate artefacts such as power line interference, electrode contact noise, motion artefacts, muscle contraction, baseline wandering, and high-frequency noise[11], a three-stage automated pre-processing pipeline was applied based on the framework proposed by Zheng[12] :

**Outlier removal**: Each ECG signal was detrended using a cumulative moving average per lead to isolate short-term fluctuations. Outliers were subsequently removed by clipping the ECG at a max voltage before re-inserting the trend data, restoring any lost information.

**Denoising**: A bandpass Butterworth filter (0.5–50 Hz) was applied to remove baseline wandering and high-frequency noise introduced by the 500 Hz sampling frequency, preserving clinically relevant signal components.

**Smoothing**: Finally, a local polynomial regression smoother (LOESS)[2] was applied to remove the effect of baseline wandering and remaining artefacts in the signal. Following this, each ECG was normalised per-participant via Z-score normalisation.

**Quality control checks:** ECG. signals with a signal-to-noise ratio (SNR) below 0 dB, insufficient heartbeats (less than 5 during rest, 300 during exercise, or 20 during recovery), or evidence of significant baseline interference (maximum amplitude >1.5 mV, baseline wander-to-QRS ratio >3:1, or QRS amplitude <250 µV) were excluded from further analysis.

We define SNR as the ratio of the standard deviation of the median heartbeat amplitude (signal component) to the median amplitude across all heartbeats (noise component), measured over windows of 5 beats at rest and 20 beats during exercise. Baseline interference was assessed using a Fast Fourier Transform (FFT) to isolate low-frequency components.

##### 1.1.2 Feature engineering

**QRS detection and wave delineation**

We implemented QRS detection and wave delineation  using a 1-d convolutional neural network (CNN), proposed by Stefan van Duijvenboden et al ,2023.[5] The model had previously been trained on lead 1 of the UK biobank cardio assessment sub-study to detect premature ventricular contractions (PVCs) and returned the probability that a given ECG segment was: (I) normal QRS complex, (ii) PVC or (iii) neither. Once QRS complexes are identified, an extended window is defined to include the P-wave, QRS complex, and T-wave. A second CNN is then used to delineate the onset and offset of each wave, identifying the P, Q, R, S, and T points

**Morphological feature extraction**

Candidate Morphological features consisted of the average PR, RR, QRS, and QTc interval durations. We compute the average PR and RR interval by calculating the elapsed time between the onset set of the preceding P and R wave and the subsequent R wave, respectively, this is then averaged across all beats in the signal. For QRS and QTc extraction, we first compute the median heartbeat via a Hilbert transform. The QRS duration is then measured as the time elapsed between the onset of the Q wave and subsequent S wave whilst the QT interval is the time elapsed between the Q and T wave. The QTc is then calculated using Bazett's[1] and Fridericia's[6] formulas for resting and exercise ECG segments, respectively.

##### Rhythm feature extraction

Using the RR interval time series obtained from wave delineation, we constructed a Poincare representation of first-order derivatives for consecutive RR intervals (Section B figure s2). ECG rhythm can then be characterised by specific patterns present in the Poincare representation, allowing for further feature extraction. Following the example in Skar et al., 2008, the Poincare plot is split into 13 unique segments and divided into bins of 30ms by 30ms. We defined normal sinus rhythm as any beat within the origin segment (segments 0), which had a radius of 80ms. We then count the number of beats in each segment and bin, to capture specific traits of relating to the rhythm of the ECG The definition and formula used to derive each feature is presented in Tables 1A-2A.

#### Table 1A: ECG Morphological features.

| ECG morphological features | Description |
|---|---|
| Mean PR interval | Time interval between the peak of the P wave and the peak of the R wave |
| Mean QRS duration | Total duration from the start of the Q wave to the end of the S wave |
| Mean QTc interval | Time interval between the peak of the Q wave and the peak of the T wave |
| Mean RR interval | Time interval between the peak of the R wave to the peak of the subsequent R wave R wave |

#### Table 2A: ECG Rhythm features

| Feature | Description | Formula |
|---|---|---|
| COSEn entropy | Quantifies the variability in the RR interval, by modelling the probability that the RR of a given set of beats will closely match with another set in the ECG.<br><br>Higher CoseEn indicates a more irregular cardiac cycle. | **SampEn − ln(2$r$) − ln (mean RR interval)**<br><br>Where:<br>SampEn = −ln(p) =−ln(A/B) − ln(B) − ln(A)<br><br>r: Maximum distance of two RR intervals to be considered a match<br>A = Number of RR interval matches within given range<br>B =Number of RR intervals within a given range<br>P = Conditional probability of a match A/B |
| Number of Premature ventricular contractions | Number of ectopic beats observed per 15-secon ECG window. | $Totoal\ PVC = \sum Observed\ PVC$ |
| Irregularity Evidence | Number of irregular heartbeats defined as beats outside of Normal sinus rhythm (NSR). | $\sum_{n=1}^{12} BinCount_n$ |

| | | |
|---|---|---|
| Regularity Evidence | Number of short-term median RR-windows, defined as 3 consecutive beats, which differed no more than 10ms from the median RR of the full 15 second segment. | $\sum_{i=1}^{n=w/m} \begin{cases} \lvert RR_{i\,short} - RR_{long} \rvert \leq 10ms : 1 \\ \lvert RRi_{short} - RR_{long} \rvert > 10ms : 0 \end{cases}$<br>Where:<br>w = The number of RR intervals in the current window<br>m = The number of RR intervals in a single segment<br>$RR_{long}$ : Median $(RR_1, RR_1, \ldots, RR_w,)$<br>$RR_{i\,short}$ : Median $(RR_j \ldots RR_k)$<br>for j = (i-1) *m+1, k = i*m for i = 1,2,…,n |
| Density Evidence | Measures how closely clustered the changes in RR intervals are across the Poincare representation, within segments, by subtracting the number of points in a segment by the number of clusters. Lower density estimates point to a more spares distribution of beats in each cluster. | $\sum_{n=5}^{12}(PointCount_n - BinCount_n$ |
| Anisotropy Evidence | Measured the direction of change of the RR interval across the current time window. Provides a measure for the orientation of the Poincare representation. Higher Anisotropy points to a consistent direction of change and helps to distinguish between different types of arrhythmias. | $\left\lvert \sum_{n=9,11} (PointCount_n) - \sum_{n=10,12} (PointCount_n \right.$<br>$+ \left\lvert \sum_{n=6,7} (PointCount_n) - \sum_{n=5,8} (PointCount_n) \right.$ |
| PAC Evidence | Measures the number of premature atrial contractions. | $\sum_{n=1}^{4}(PointCount_n - BinCount_n)$<br>$+ \sum_{n=5,6,10}(PointCount_n - BinCount_n)$<br>$-\sum_{n=7,8,12}(PointCount_n - BinCount_n)$ |
| Evidence of Atrial fibrillation | A composite measure which measures the presence and severity of Atrial fibrillation by offsetting evidence of irregularity against the number of normal heartbeats and premature atrial contractions | Irregularity evidence –<br>$\sum_{i=1}^{n} \begin{cases} \lvert RR_{i-1} - RR_i \rvert = 0 : 1 \\ \lvert RRi_{i-1} - RR_i \rvert \neq 0 : 0 \end{cases}$<br>- 2 × PAC evidence |
| Evidence of atrial tachycardia | Assesses the presence of irregular heart rhythm, changes in RR intervals, clustering of changes in RR interval (Poincare representation), and the occurrence of compensatory pauses | Irregularity Evidence<br>+ Anisotropy Evidence + Density Evidence<br>+ Regularity Evidence -4 × PAC Evidence |

##### 1.1.3 AI ECG risk score implementation

Our AI ECG risk score comprises three core components: (i) a convolutional neural network (CNN) for extracting key ECG features, (ii) an LSTM model to capture temporal changes in the ECG over time, and (iii) a fully connected deep-survival network to evaluate overall cardiovascular risk. The CNN produces a learned representation of the ECG, highlighting critical features, which the LSTM analyses for time-based patterns. The deep-survival network then uses this learned representation to provide a single risk metric ($ECG_{AI}$), estimating the hazard of a cardiovascular event based on the ECG, analogous to the log partial hazard in traditional Cox regression.

We selected an adapted ResNet-V2 with 50 layers[7] and 1D convolutions, totalling 17M parameters, for ECG feature extraction, chosen for its state-of-the-art performance and robust validation for ECG analysis[10]. ECGs were processed in 15-second windows as 3-dimensional vectors of size (3,7500) (3 leads by 15× 500hz). The learned feature vector was of size 2048. An LSTM was added to model-to-model temporal dependencies between each ECG window. The resulting latent representation of the ECG is passed through a fully connected layer to identify the most relevant features associated with MACCE. Separately, QRISK3 risk factors are processed by a multilayer perceptron to capture non-linear interactions before being combined with the ECG-based score to generate a 10-year MACCE risk prediction.

If either the ECG or QRISK3 risk factors are unavailable, risk scores can instead be generated by taking the output of either network alone. Details of the model architecture for the LSTM and deep-survival networks are provided in Table 3A-4A.

**Model training implementation.**

We trained the model to minimise the negative log partial likelihood over a 12-year follow-up period, as outlined in the original deep-survival framework[8].Optimisation was performed using the Adam optimizer[4]. A learning rate scheduler dynamically adjusted the rate, reducing it after 5 consecutive epochs without improvement to help the optimizer reach a better local minimum. Training was distributed across four Tesla V100-SXM2 GPUs, each with 32GB of memory. The complete training protocol is outlined in Table 4A.

Model development and validation were implemented in Python 3·7 and Pytorch 1·5; training was performed using the Pycox package for deep-survival networks[9], whilst evaluation metrics were generated using both Pycox[9] and lifelines[3] packages.

**Table 3A: LSTM and Deep-survival model architecture**

| Hyper-parameter | Value |
|---|---|
| Network: LSTM network | |
| Hidden layers | 3 |
| Hidden size | 256 |
| Dropout rate | 0·2 |
| Network: ECG hazard network | |
| Hidden layers | 2 |
| Hidden nodes [layer1, layer2] | [256,128] |
| Dropout rate | 0·4 |
| Network: non-linear risk network | |
| Hidden layers | 2 |
| Hidden nodes [layer1, layer2] | [12,12] |
| Dropout rate | 0·1 |

**Table 4A: Model Training protocol for deep-survival network including hyper-parameters and testing procedure.**

| | |
|---|---|
| Batch Size | 1024 |
| Epochs | 200 |
| Loss function | Average negative log partial likelihood |
| Optimiser | Adam |
| Learning rate | $1e^{-5}$ |
| Learning rate decay | 0·05 |
| Learning rate decay patience | 10 |
| **Train-test procedure** | |
| Train split | 60% |
| Validation split | 20% |
| Test split | 20% |
| Evaluation | Boot strapping (1000 iterations) |

##### 1.2 Cox clinical implementation

Sixteen covariates were selected from the original QRISK3 risk score (Supplementary Table 3; Table 1C), Fractional polynomials for age, BMI, and interaction terms for age with BMI, smoking status, type1/2 diabetes, systolic blood pressure, deprivation, and hypertension were selected to match the original QRISK3 algorithm. Model coefficients were re-calculated using Cox Proportional hazard models to re-calibrate the model to the study population. The 10-year baseline survival and coefficients for each model are provided in Supplementary section C.

##### 1.3 Risk score generation

10-year risk scores were subsequently calculated by adjusting the baseline survival probability of no events occurring at the 10-year point by the output of the selected risk model (1). The baseline survival was calculated using Breslow's method implemented in the lifelines[3] and Pycox[9] packages for Cox clinical and deep survival models, respectively. The 10-year baseline survival rates for the respective models are provided below.

$$Risk\ score = 100 * (1 - S_{t(10)}^{e^{F(X)}}) \qquad (1)$$

*Where $S_{t(10)}$ is the baseline survival at 10-years of follow-up, and F(x) is the weighted sum of risk factors from either a Cox Proportional hazard model or deep-survival model.*

### Section B

#### Supplementary Figures

##### Supplementary Figure 1 Poincare representation of ECG rhythm profiles

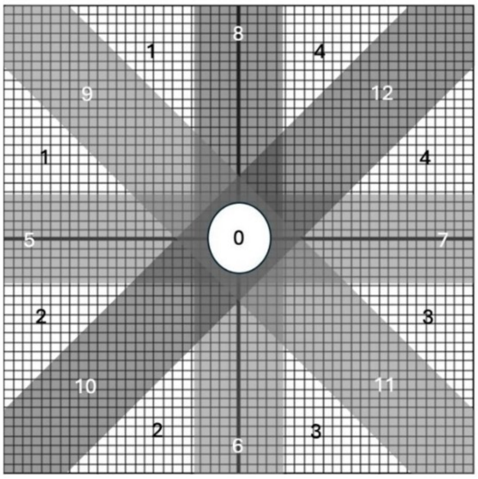

(a)

| Segment | Sequence | ΔRR Values |
|---|---|---|
| 0 | SSS/LLL | \|ΔRR\| < 80ms |
| 1 | S-L-S | \|ΔRR(i-1)\| ≠ \|ΔRR(i)\| |
| 2 | L-M-S | \|ΔRR(i-1)\| ≠ \|ΔRR(i)\| |
| 3 | L-S-L | \|ΔRR(i-1)\| ≠ \|ΔRR(i)\| |
| 4 | S-M-L | \|ΔRR(i-1)\| ≠ \|ΔRR(i)\| |
| 5 | L-L-S | At least one \|ΔRR\| in segment 0 |
| 6 | L-S-S | At least one \|ΔRR\| in segment 0 |
| *7* | S-S-L | At least one \|ΔRR\| in segment 0 |
| *8* | *S-L-L* | At least one \|ΔRR\| in segment 0 |
| *9* | *S-L-S* | ΔRR(i-1) ≅ -ΔRR(i) |
| *10* | *S-L-S* | ΔRR(i-1) ≅ ΔRR(i) |
| *11* | *L-S-L* | ΔRR(i-1) ≅ -ΔRR(i) |
| *12* | *L-M-L* | ΔRR(i-1) ≅ ΔRR(i) |

(b)

*(a) 2-D histogram (Poincare plot) showing the rate of Change in* ΔRR at one time step i on the x axis against the ΔRR at the previous time step i-1 on the y axis. The Poincare plot is divided into 13 segments denoted in (b) and populated by the rate of change in RR interval (ΔRR(i), ΔRR(i-1)). Segment 0 represents heart beats in normal sinus rhythm and has a radius of 80 ms. (b) divides the Poincare plot into segments based on the rate of change in RR interval, small (D) medium (M) or large(L), with a sequence of S-M-L indicating a short RR interval followed by a medium RR interval followed by a large RR interval.

*Abbreviations: S = short RR interval; M = medium RR interval; L = large RR interval.*
*Adapted from: Shantanu Sarkar* et al, A Detector for a Chronic Implantable Atrial Tachyarrhythmia Monitor.*

**Supplementary figure 2: Smoothed calibration curves for 10-year MACCE risk prediction using ECG risk scores derived from deep survival neural networks applied to raw ECG data measured during (A) rest, (B) peak exercise, (C) late-stage recovery, and (D) the complete 7-minute 15-second submaximal exercise ECG recording**

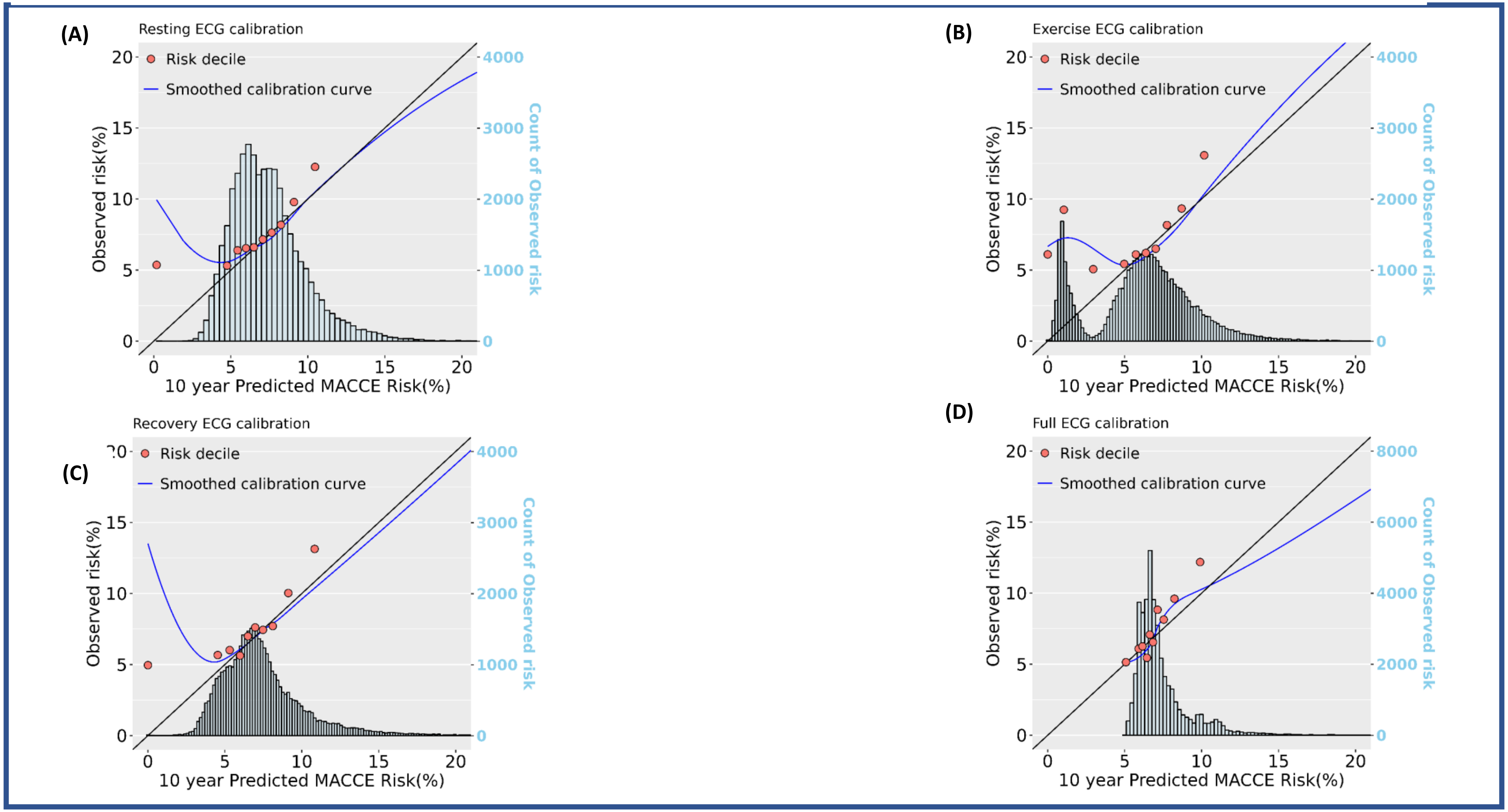

The red dots show the mean predicted risk vs observed risk at each risk tenth. The black line shows perfect calibration, where the predicted risk perfectly matches the observed risk. ECG scores were derived from a deep survival network trained on ECG recordings during 15-seconds of rest, the final 15-seconds of exercise (peak exercise intensity), final 15-seconds of recovery and full 7-minute 15-second submaximal exercise ECG recording.

**Supplementary figure 3 : Smoothed calibration curves for 10-year MACCE risk prediction using Age + sex + ECG risk scores derived from ECG measurements during (A) rest, (B) peak exercise, (C) late-stage recovery, and (D) the complete 7-minute 15-second submaximal exercise ECG recording.**

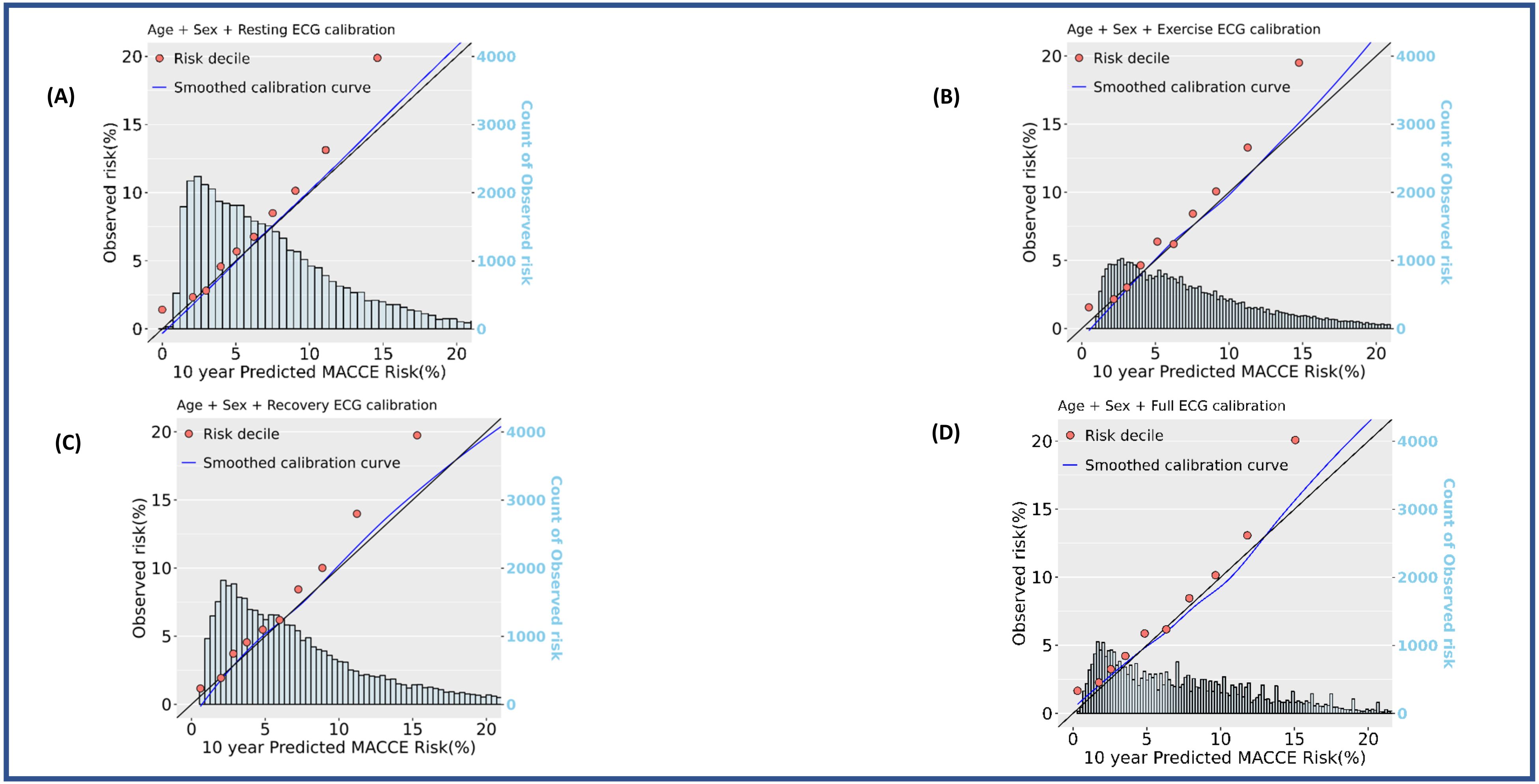

The red dots show the mean predicted risk vs observed risk at each risk tenth. The black line shows perfect calibration, where the predicted risk perfectly matches the observed risk. Observed risk was estimated using pseudo probabilities obtained via a jackknife approach. ECG scores were derived from a deep survival network trained on ECG recordings during 15-seconds of rest, the final 15-seconds of exercise (peak exercise intensity), the final 15-seconds of recovery and the full 7-minute 15-second submaximal exercise ECG recording.

**Supplementary figure 4 : Smoothed calibration curves for 10-year MACCE risk prediction using Cox clinical + ECG risk scores derived from ECG measurements during (A) rest, (B) peak exercise, (C) late-stage recovery, and (D) the complete 7-minute 15-second submaximal exercise ECG recording.**

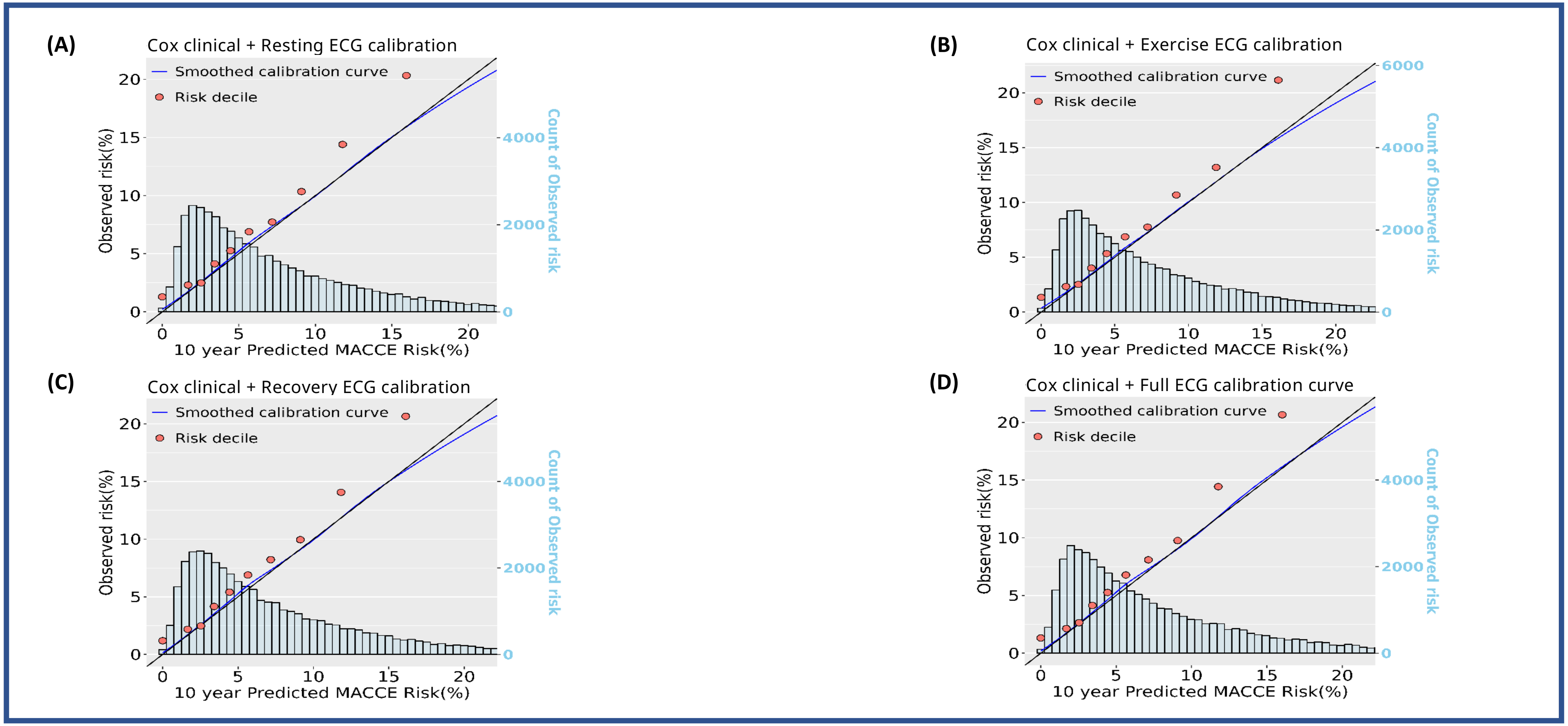

The red dots show the mean predicted risk vs observed risk at each risk tenth. The black line shows perfect calibration, where the predicted risk perfectly matches the observed risk. Observed risk was estimated using pseudo probabilities obtained via a jackknife approach. ECG scores were derived from a deep survival network trained on ECG recordings during 15-seconds of rest, the final 15-seconds of exercise (peak exercise intensity), the final 15-seconds of recovery and full 7-minute 15-second submaximal exercise ECG recording. Cox clinical: Age FP1 + Age FP2 + BMI FP1 + BMI FP2 + SBP + SBP5 + Diabetes type 1 + Diabetes type 2 + HDL ratio + Townsend deprivation index + Smoker former + Smoker current + Smoker never + Ethnic Background + Corticosteroid + Migraine + Hypertension + Severe mental illness + Age FP1 × Townsend deprivation index + Age FP2 × Townsend deprivation index + Age FP1 × BMI FP 1 + Age FP1 × BMI FP 2 + Age FP1 × BP + Age FP1 × Diabetes type 1 + Age FP1 × Diabetes type 2 + Age FP1 × hypertension + Age FP2 × BMI FP1 + Age FP2 × BMI FP2 + Age FP2 × SBP + Age FP2 × Diabetes type1 + Age FP2 × Diabetes type2 + Age FP2 × Hypertension. Fractional polynomials: *Age FP1 (males)* = $(age/10)^{-1}$, *Age FP2 (males)* = $(age/10)^3$, *BMI FP 1 (males)* = $(BMI/10)^{-2}$, *BMI FP 2 (males)* = $(BMI/10)^{-2}\ X\ log(BMI/10)$, *Age FP1 (Females)* = $age/10^{-2}$, *Age FP2 (Females)* = *age/10*, *BMI FP 1 (Females)* = $(BMI/10)^{-2}$, *BMI FP 2 (Females)* = $(BMI/10)^{-2}\ X\ log\ (BMI/10)$.

**Supplementary figure 5 : Smoothed calibration curves for 10-year MACCE risk prediction using clinical deep survival network (DSN) models developed from QRISK3 risk factors and ECG data measured during (A) rest, (B) peak exercise, (C) late-stage recovery, and (D) the complete 7-minute 15-second submaximal exercise ECG recording**

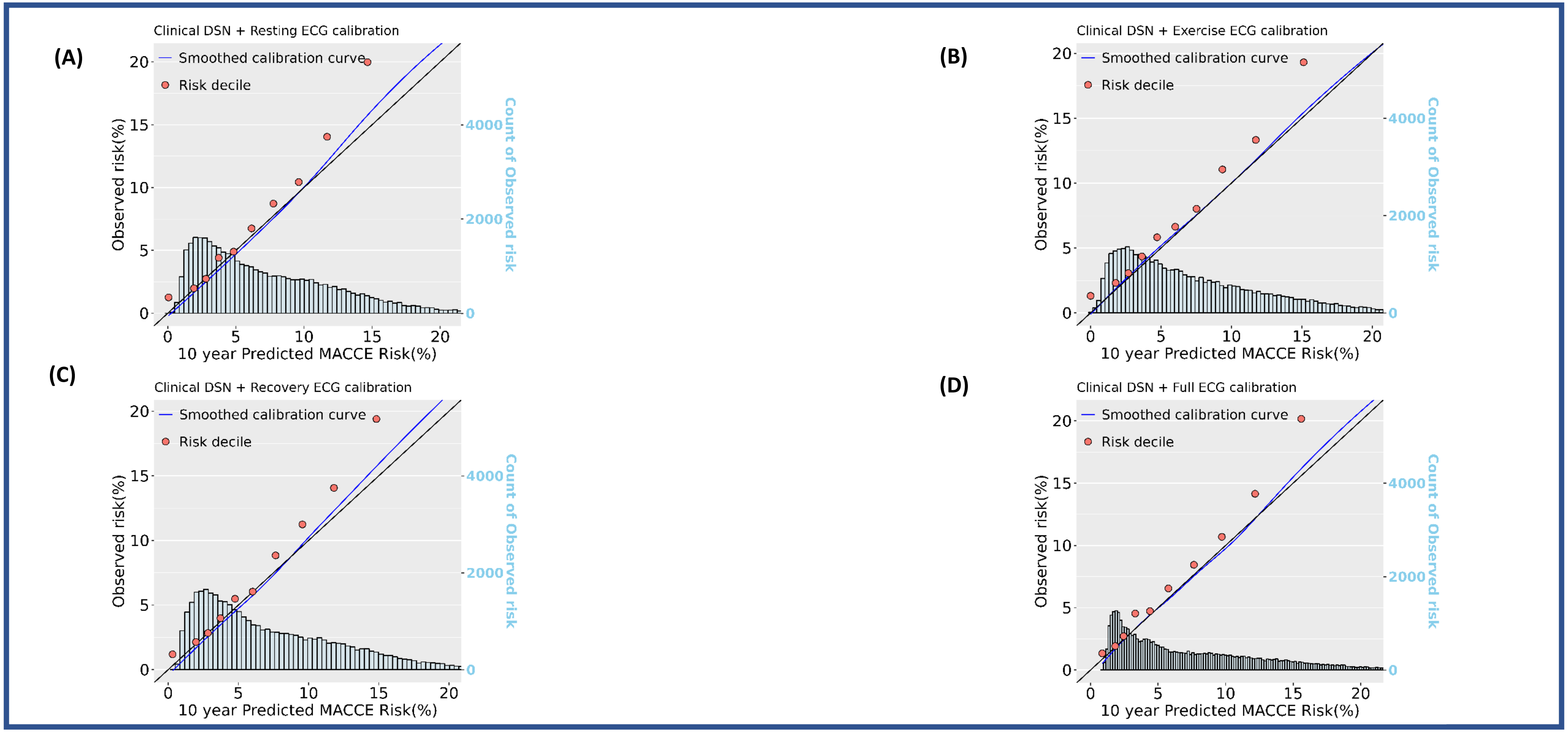

Abbreviations: DSN = Deep survival network

The red dots show the mean predicted risk vs observed risk at each risk tenth. The black line shows perfect calibration, where the predicted risk perfectly matches the observed risk. Observed risk was estimated using pseudo probabilities obtained via a jackknife approach. ECG scores were derived from a deep survival network trained on ECG recordings during 15-seconds of rest, the final 15-seconds of exercise (peak exercise intensity), the final 15-seconds of recovery and full 7-minute 15-second submaximal exercise ECG recording. Clinical DSN: Age + BMI + systolic blood-pressure + SBP5 +Diabetes type 1+ Diabetes type 2 + HDL ratio + Townsend deprivation index+ Smoker former + Smoker current + Smoker never+ Ethnic Background+ Corticosteroid+ Migraine + Hypertension + Severe mental illness.

**Supplementary figure 6: Hazard ratio per standard deviation increase in Deep survival ECG risk scores ($ECG_{AI}$) and conventional ECG parameters derived from 15 second resting ECG recordings.**

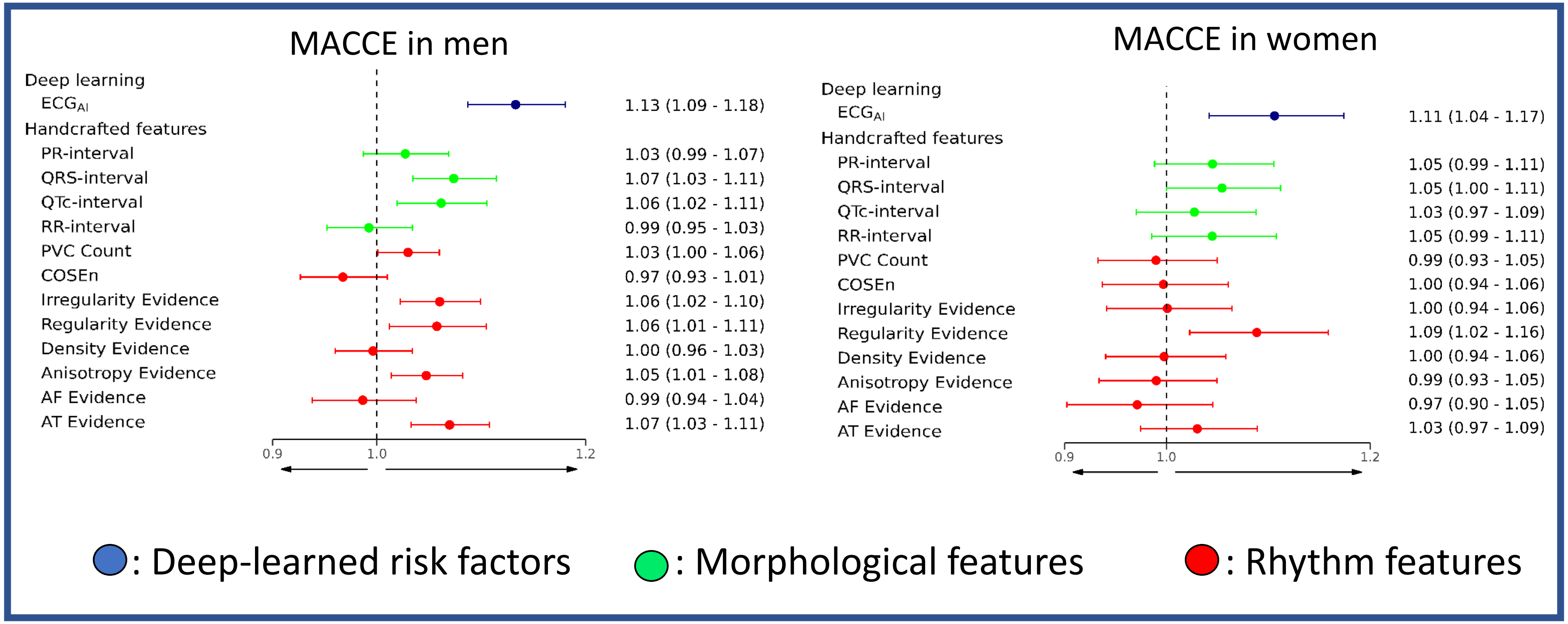

Hazard ratios are derived from separate Cox proportional hazard models adjusted for age, sex, smoking status, social deprivation measured by Townsend deprivation index, and ethnicity. Irregularity, Regularity, Anisotropy and AF evidence were adjusted for the number of beats in the signal to account for higher absolute values in participants with faster heart rates. Hazard ratios are reported per standard deviation increase. Cox proportional hazard models were developed on the aggregated test sets across five cross-validation folds, consisting of 35,597 participants and 3,463 (9·7%) MACCE over 12.5 years of follow-up.

**Supplementary figure 7: Hazard ratio per standard deviation increase in Deep survival ECG risk scores ($ECG_{AI}$) and conventional ECG parameters derived from the final 15 seconds (maximum intensity of exercise) of a 6-min period of submaximal cycle ergometry exercise test.**

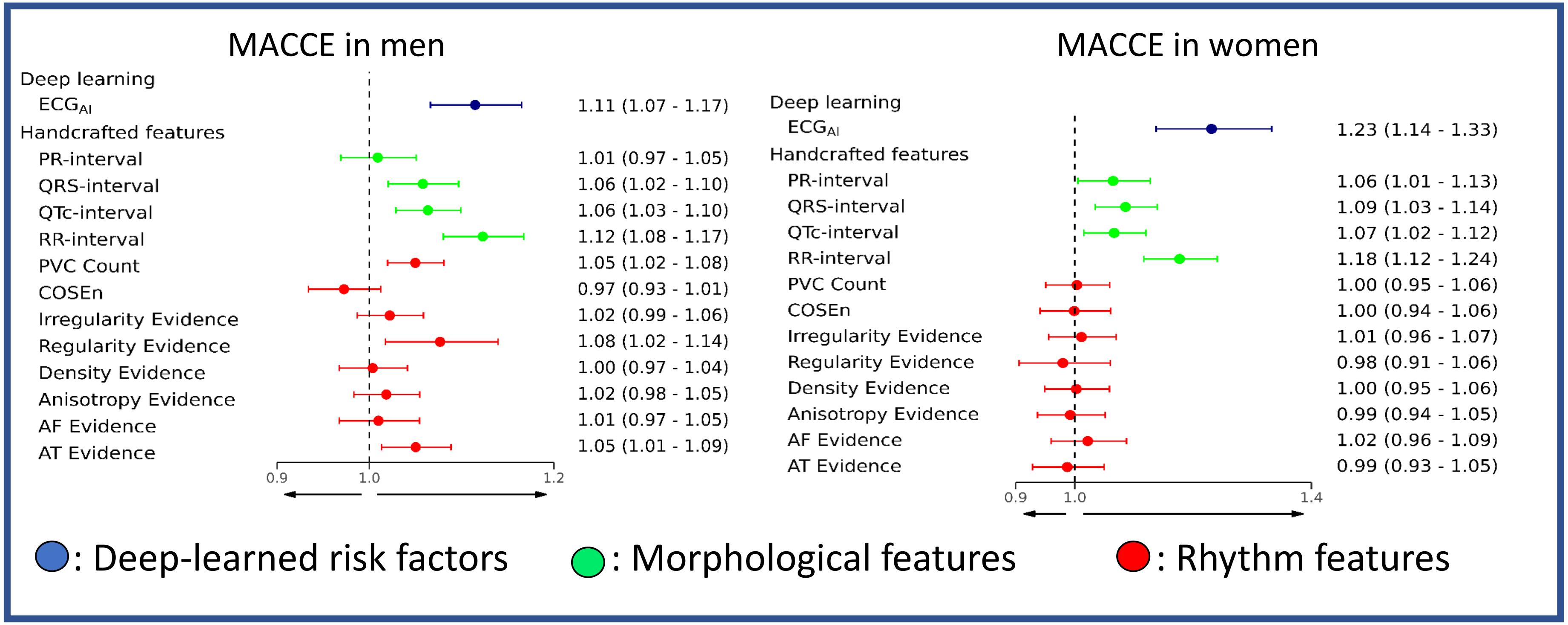

Hazard ratios are derived from separate Cox proportional hazard models adjusted for age, sex, smoking status, social deprivation measured by Townsend deprivation index, and ethnicity. Irregularity, Regularity, Anisotropy and AF evidence were adjusted for the number of beats in the signal to account for higher absolute values in participants with faster heart rates. Hazard ratios are reported per standard deviation increase. Cox proportional hazard models were developed on the aggregated test sets across five cross-validation folds, consisting of 35,597 participants and 3,463 (9·7%) MACCE over 12.5 years of follow-up.

**Supplementary figure 8**: **Hazard ratio per standard deviation increases in Deep survival ECG risk scores ($ECG_{AI}$) and conventional ECG parameters derived from the final 15 seconds of 1-min post exercise recovery period**

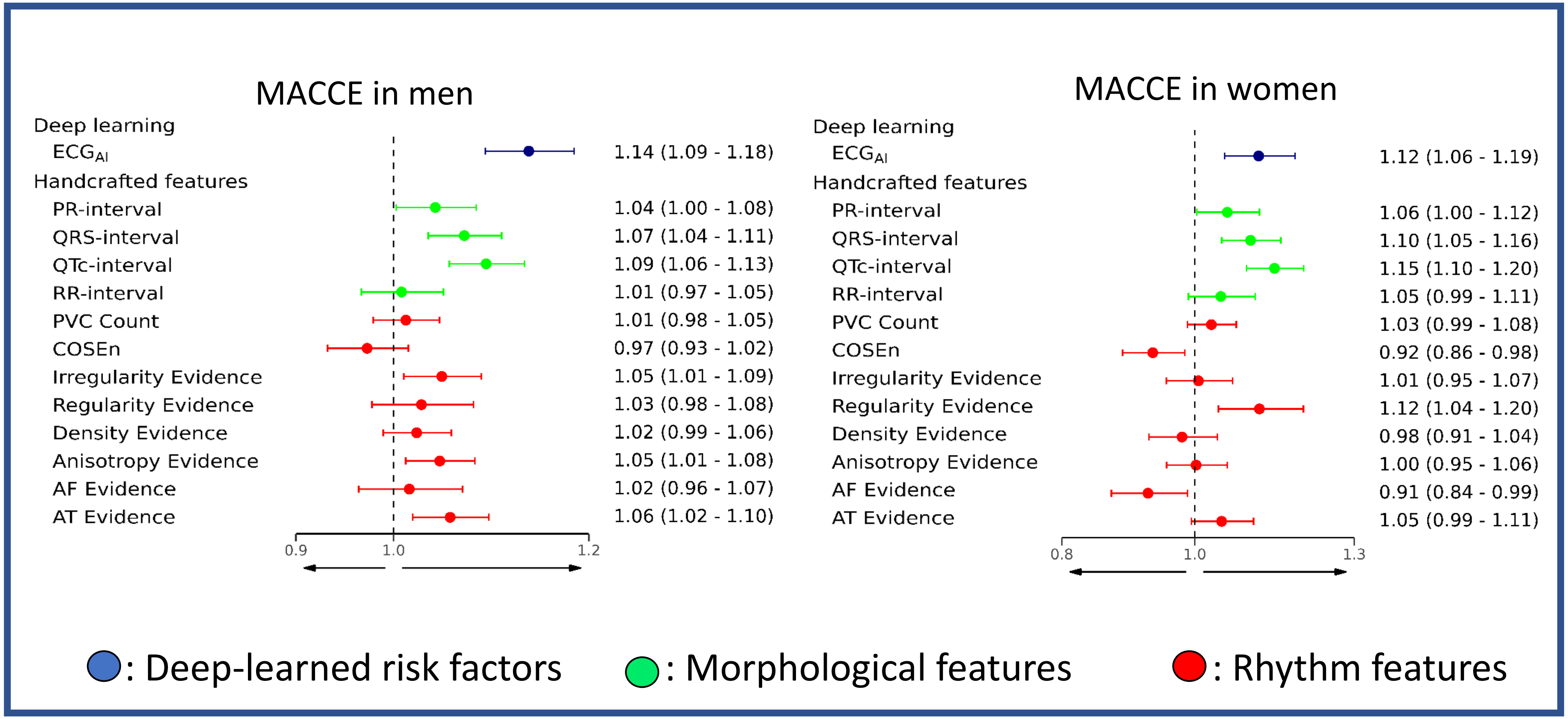

Hazard ratios are derived from separate Cox proportional hazard models adjusted for age, sex, smoking status, social deprivation measured by Townsend deprivation index, and ethnicity. Irregularity, Regularity, Anisotropy and AF evidence were adjusted for the number of beats in the signal to account for higher absolute values in participants with faster heart rates. Hazard ratios are reported per standard deviation increase. Cox proportional hazard models were developed on the aggregated test sets across five cross-validation folds, consisting of 35,597 participants and 3,463 (9·7%) MACCE over 12.5 years of follow-up.

**Supplementary Figure 9 : Forest plot of Harrell's C-index for 10-year MACCE risk prediction comparing Age + sex, Cox clinical, and Clinical Deep Survival Network (DSN) models combined with 15-second resting ECG data ($ECG_{AI}$) in 41,076 UK Biobank participants (2009–2013)**

| | N | MACCE | C-index (95% CI) | Change in C-index (95% CI) |
|---|---|---|---|---|
| **Male** | | | | |
| Age + Sex | 20419 | 2092 | 0.663 (0.653, 0.673) | |
| Age + Sex + $ECG_{AI}$ | | | 0.675 (0.666, 0.685) | +0.011 (+0.006, +0.016) |
| Cox clinical | | | 0.675 (0.666, 0.685) | +0.012 (+0.006, +0.019) |
| Cox clinical + $ECG_{AI}$ | | | 0.677 (0.668, 0.686) | +0.013 (+0.006, +0.018) |
| Clinical DSN | | | 0.678 (0.669, 0.688) | +0.014 (+0.007, +0.021) |
| Clinical DSN + $ECG_{AI}$ | | | 0.681 (0.671, 0.691) | +0.017 (+0.010, +0.023) |
| **Female** | | | | |
| Age + Sex | 20657 | 961 | 0.670 (0.656, 0.682) | |
| Age + Sex + $ECG_{AI}$ | | | 0.673 (0.660, 0.686) | +0.004 (-0.002, +0.009) |
| Cox clinical | | | 0.674 (0.660, 0.688) | +0.005 (-0.004, +0.015) |
| Cox clinical + $ECG_{AI}$ | | | 0.675 (0.663, 0.690) | +0.006 (-0.003, +0.015) |
| Clinical DSN | | | 0.678 (0.664, 0.691) | +0.009 (0.000, +0.018) |
| Clinical DSN+ $ECG_{AI}$ | | | 0.689 (0.678, 0.704) | +0.019 (+0.010, +0.029) |

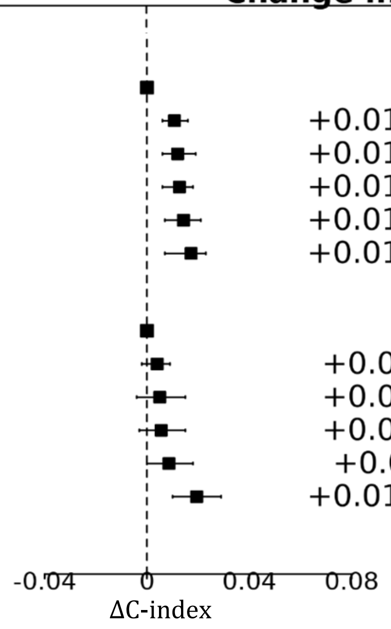

Abbreviations: CI = Confidence interval, MACCE = Major Adverse Cardiovascular & Cerebrovascular Events; DSN: Deep survival network; 95% confidence intervals were derived via 1000 bootstrap iterations.

Squares represent change in C-index compared to age + sex models and horizontal lines are 95% confidence intervals generated using 1,000 bootstrap samples. 95 % CIs were derived via bootstrapping for 1000 iterations: significance threshold = 0·05.

$ECG_{AI}$ scores were derived from a deep survival network trained on ECG recordings obtained during the final 15-seconds of exercise, corresponding to the maximum intensity exercise obtained

Clinical DSN: Age, BMI, systolic blood-pressure, SBP5, Diabetes type 1+ Diabetes type 2, HDL ratio, Townsend deprivation index+ Smoker former, Smoker current, Smoker never+ Ethnic Background, Corticosteroid+ Migraine, Hypertension, Severe mental illness, Rheumatoid Arthritis.

Cox clinical: Age FP1 + Age FP2 + BMI FP1 + BMI FP2 + SBP + SBP5 + Diabetes type 1 + Diabetes type 2 + HDL ratio + Townsend deprivation index + Smoker former + Smoker current + Smoker never + Ethnic Background + Corticosteroid + Migraine + Hypertension + Severe mental illness + Age FP1 × Townsend deprivation index + Age FP2 × Townsend deprivation index + Age FP1 × BMI FP 1 + Age FP1 × BMI FP 2 + Age FP1 × BP + Age FP1 × Diabetes type 1 + Age FP1 × Diabetes type 2 + Age FP1 × hypertension + Age FP2 × BMI FP1 + Age FP2 × BMI FP2 + Age FP2 × SBP + Age FP2 × Diabetes type1 + Age FP2 × Diabetes type2 + Age FP2 × Hypertension.

Fractional polynomials (men): Age FP 1 = $(age/10)^{-1}$; Age FP 2 = $(age/10)^3$; BMI FP 1 = $(BMI/10)^{-2}$; BMI FP 2 = $(BMI/10)^{-2}\ X \log(BMI/10)$.
Fractional polynomials (Female): Age FP 1 = $age/10^{-2}$; Age FP 2 = age/10; BMI FP 1 = $(BMI/10)^{-2}$; BMI FP 2 = $(BMI/10)^{-2}\ X \log\ (BMI/10)$.

**Supplementary Figure 10 : Forest plot of Harrell's C-index for 10-year MACCE risk prediction comparing Age + sex, Cox clinical, and Clinical Deep Survival Network (DSN) models combined with $ECG_{AI}$ derived from the final 15 seconds of exercise ECG data in 41,076 UK Biobank participants (2009–2013)**

| | N | MACCE | C-index (95% CI) | Change in C-index (95% CI) |
|---|---|---|---|---|
| **Male** | | | | |
| Age + Sex | 20419 | 2092 | 0.663 (0.653, 0.673) | |
| Age + Sex + $ECG_{AI}$ | | | 0.670 (0.661, 0.679) | +0.006 (+0.003, +0.010) |
| Cox clinical | | | 0.675 (0.666, 0.685) | +0.012 (+0.006, +0.019) |
| Cox clinical + $ECG_{AI}$ | | | 0.678 (0.670, 0.689) | +0.014 (+0.007, +0.020) |
| Clinical DSN | | | 0.678 (0.669, 0.688) | +0.014 (+0.007, +0.021) |
| Clinical DSN + $ECG_{AI}$ | | | 0.667 (0.657, 0.677) | +0.003 (-0.006, +0.011) |
| **Female** | | | | |
| Age + Sex | 20657 | 961 | 0.670 (0.656, 0.682) | |
| Age + Sex + $ECG_{AI}$ | | | 0.674 (0.662, 0.688) | +0.005 (-0.002, +0.011) |
| Cox clinical | | | 0.674 (0.660, 0.688) | +0.005 (-0.004, +0.015) |
| Cox clinical + $ECG_{AI}$ | | | 0.677 (0.664, 0.693) | +0.007 (-0.001, +0.015) |
| Clinical DSN | | | 0.678 (0.664, 0.691) | +0.009 (0.000, +0.018) |
| Clinical DSN + $ECG_{AI}$ | | | 0.679 (0.666, 0.693) | +0.009 (-0.002, +0.021) |

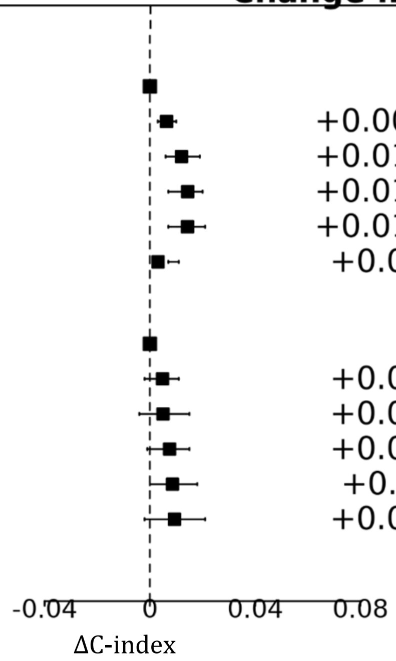

Abbreviations: CI = Confidence interval, MACCE = Major Adverse Cardiovascular & Cerebrovascular Events; DSN: Deep survival network95% confidence intervals were derived via 1000 bootstrap iterations.

Squares represent change in C-index compared to age + sex models and horizontal lines are 95% confidence intervals generated using 1,000 bootstrap samples. 95 % CIs were derived via bootstrapping for 1000 iterations: significance threshold = 0·05.

$ECG_{AI}$ scores were derived from a deep survival network trained on ECG recordings obtained during the final 15-seconds of exercise, corresponding to the maximum intensity exercise obtained.

Clinical DSN: Age, BMI, systolic blood-pressure, SBP5, Diabetes type 1+ Diabetes type 2, HDL ratio, Townsend deprivation index+ Smoker former, Smoker current, Smoker never+ Ethnic Background, Corticosteroid+ Migraine, Hypertension, Severe mental illness, Rheumatoid Arthritis.

Cox clinical: Age FP1 + Age FP2 + BMI FP1 + BMI FP2 + SBP + SBP5 + Diabetes type 1 + Diabetes type 2 + HDL ratio + Townsend deprivation index + Smoker former + Smoker current + Smoker never + Ethnic Background + Corticosteroid + Migraine + Hypertension + Severe mental illness + Age FP1 × Townsend deprivation index + Age FP2 × Townsend deprivation index + Age FP1 × BMI FP 1 + Age FP1 × BMI FP 2 + Age FP1 × BP + Age FP1 × Diabetes type 1 + Age FP1 × Diabetes type 2 + Age FP1 × hypertension + Age FP2 × BMI FP1 + Age FP2 × BMI FP2 + Age FP2 × SBP + Age FP2 × Diabetes type1 + Age FP2 × Diabetes type2 + Age FP2 × Hypertension.

Fractional polynomials (men): Age FP 1 = $(age/10)^{-1}$; Age FP 2 = $(age/10)^3$; BMI FP 1 = $(BMI/10)^{-2}$; BMI FP 2 = $(BMI/10)^{-2}\ X \log(BMI/10)$.
Fractional polynomials (Female): Age FP 1 = $age/10^{-2}$; Age FP 2 = age/10; BMI FP 1 = $(BMI/10)^{-2}$; BMI FP 2 = $(BMI/10)^{-2}\ X \log\ (BMI/10)$.

**Supplementary Figure 11 : rest plot of Harrell's C-index for 10-year MACCE risk prediction comparing Age + sex, Cox clinical, and Clinical Deep Survival Network (DSN) models combined with ECGAI derived from the final 15 seconds of recovery ECG data in 41,076 UK Biobank participants (2009–2013)**

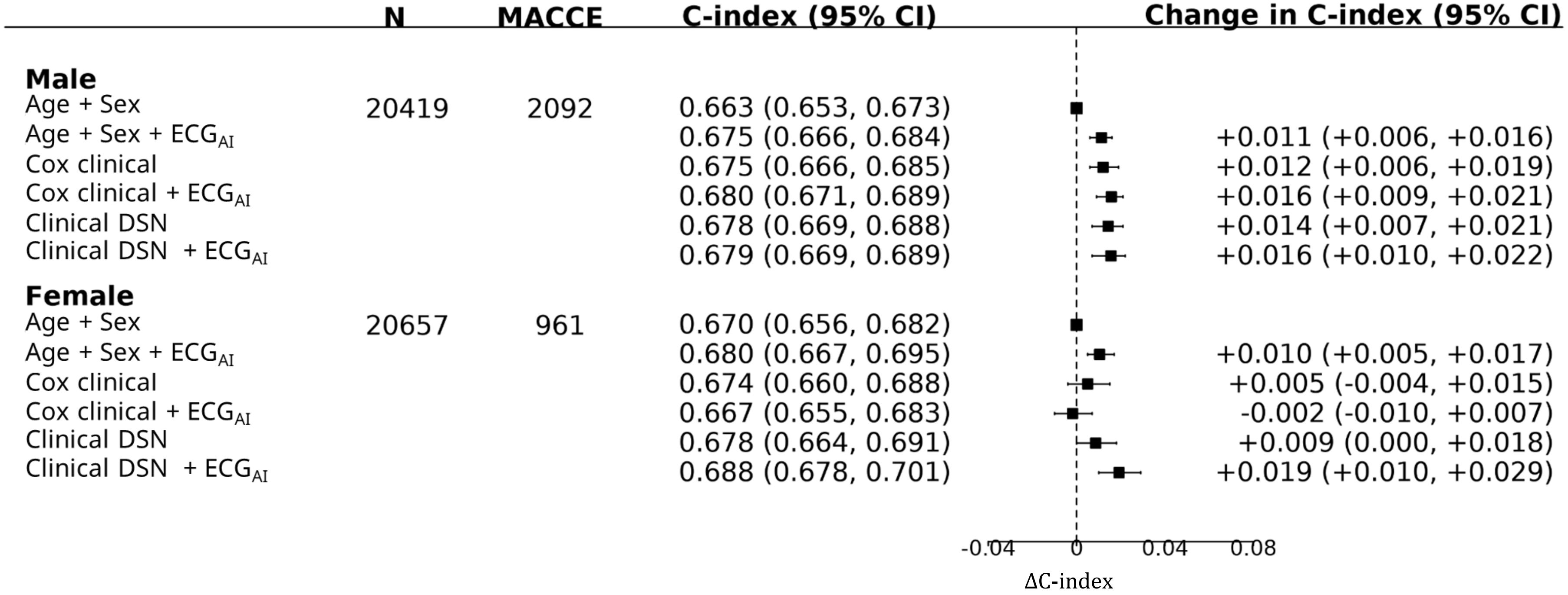

Abbreviations: CI = Confidence interval, MACCE = Major Adverse Cardiovascular & Cerebrovascular Events; DSN: Deep survival network95% confidence intervals were derived via 1000 bootstrap iterations.

Squares represent change in C-index compared to age + sex models and horizontal lines are 95% confidence intervals generated using 1,000 bootstrap samples. 95 % CIs were derived via bootstrapping for 1000 iterations: significance threshold = 0·05.

$ECG_{AI}$ scores were derived from a deep survival network trained on ECG recordings obtained during the final 15-seconds of exercise, corresponding to the maximum intensity exercise obtained

Clinical DSN: Age, BMI, systolic blood-pressure, SBP5, Diabetes type 1+ Diabetes type 2, HDL ratio, Townsend deprivation index+ Smoker former, Smoker current, Smoker never+ Ethnic Background, Corticosteroid+ Migraine, Hypertension, Severe mental illness, Rheumatoid Arthritis.

Cox clinical: Age FP1 + Age FP2 + BMI FP1 + BMI FP2 + SBP + SBP5 + Diabetes type 1 + Diabetes type 2 + HDL ratio + Townsend deprivation index + Smoker former + Smoker current + Smoker never + Ethnic Background + Corticosteroid + Migraine + Hypertension + Severe mental illness + Age FP1 × Townsend deprivation index + Age FP2 × Townsend deprivation index + Age FP1 × BMI FP 1 + Age FP1 × BMI FP 2 + Age FP1 × BP + Age FP1 × Diabetes type 1 + Age FP1 × Diabetes type 2 + Age FP1 × hypertension + Age FP2 × BMI FP1 + Age FP2 × BMI FP2 + Age FP2 × SBP + Age FP2 × Diabetes type1 + Age FP2 × Diabetes type2 + Age FP2 × Hypertension.

Fractional polynomials (men): Age FP 1 = $(age/10)^{-1}$; Age FP 2 = $(age/10)^3$; BMI FP 1 = $(BMI/10)^{-2}$; BMI FP 2 = $(BMI/10)^{-2}\ X \log(BMI/10)$.
Fractional polynomials (Female): Age FP 1 = $age/10^{-2}$; Age FP 2 = age/10; BMI FP 1 = $(BMI/10)^{-2}$; BMI FP 2 = $(BMI/10)^{-2}\ X \log(BMI/10)$.

### Supplementary Tables

#### Supplementary Table 1 ICD-(9/10) codes used to define MACCE

| ICD-10 Code | Condition |
|---|---|
| I20 | Angina pectoris |
| I21 | Acute myocardial infarction |
| I210 | Acute transmural myocardial infarction of anterior wall |
| I211 | Acute transmural myocardial infarction of inferior wall |
| I212 | Acute transmural myocardial infarction of other sites |
| I213 | Acute transmural myocardial infarction of unspecified site |
| I214 | Acute subendocardial myocardial infarction |
| I219 | Acute myocardial infarction, unspecified |
| I22 | Subsequent myocardial infarction |
| I220 | Subsequent myocardial infarction of anterior wall |
| I221 | Subsequent myocardial infarction of inferior wall |
| I228 | Subsequent myocardial infarction of other sites |
| I229 | Subsequent myocardial infarction of unspecified site |
| I23 | Certain current complications following acute myocardial infarction |
| I230 | Haemopericardium as current complication following acute myocardial infarction |
| I231 | Atrial septal defect as current complication following acute myocardial infarction |
| I232 | Ventricular septal defect as current complication following acute myocardial infarction |
| I233 | Rupture of cardiac wall without haemopericardium as current complication following acute myocardial infarction |

| | |
|---|---|
| I234 | Rupture of chordae tendineae as current complication following acute myocardial infarction |
| I235 | Rupture of papillary muscle as current complication following acute myocardial infarction |
| I236 | Thrombosis of atrium, auricular appendage and ventricle as current complications following acute myocardial infarction |
| I238 | Thrombosis of atrium, auricular appendage and ventricle as current complications following acute myocardial infarction |
| I50 | Heart failure |
| I50·0 | Congestive heart failure |
| I50·1 | Left ventricular failure |
| I50·9 | Heart failure, unspecified |
| I472 | Ventricular tachycardia |
| I490 | Ventricular fibrillation and flutter |
| I46·0 | Cardiac arrest with successful resuscitation |

| | |
|---|---|
| I461 | Sudden cardiac death, so described |
| I469 | Cardiac arrest, unspecified |
| I470 | Re-entry ventricular arrhythmia |
| Cerebral infarction | I63 |
| Stroke not specified as haemorrhage or infarction | I64 |
| Transient ischaemic attack and related syndromes | G45 |
| ICD-9 codes | Condition |
| 4109 | Acute myocardial infarction |
| 4280 | Congestive heart failure |
| 4281 | Left heart failure |
| 4289 | Heart failure, unspecified |

**Supplementary Table 2 Primary care codes for Major Adverse Cardiovascular and Cerebrovascular Events (MACCE)**

| | Read 2 code | Read 3 code | Term 3 code | Read 2 description | Read 3 description |
|---|---|---|---|---|---|
| 0 | G3... | XE2uV | Y201T | Ischaemic heart disease | Ischaemic heart disease |
| 1 | G30.. | XE0Uh | Y202N | Acute myocardial infarction | Acute myocardial infarction |
| 2 | G300. | G300. | Y202d | Acute anterolateral infarction | Acute anterolateral myocardial infarction |
| 3 | G301. | G301. | Y202Q | Other specified anterior myocardial infarction | Other specified anterior myocardial infarction |
| 4 | G3010 | G3010 | Y202V | Acute anteroapical infarction | Acute anteroapical infarction |
| 5 | G3011 | G3011 | Y202a | Acute anteroseptal infarction | Acute anteroseptal myocardial infarction |
| 6 | G301z | G301z | Y202R | Anterior myocardial infarction NOS | Anterior myocardial infarction NOS |
| 7 | G302. | G302. | Y202l | Acute inferolateral infarction | Acute inferolateral myocardial infarction |
| 8 | G303. | G303. | Y202h | Acute inferoposterior infarction | Acute inferoposterior infarction |
| 9 | G304. | G304. | Y202x | Posterior myocardial infarction NOS | Posterior myocardial infarction NOS |
| 10 | G305. | G305. | Y202p | Lateral myocardial infarction NOS | Lateral myocardial infarction NOS |
| 11 | G306. | G306. | Y202w | True posterior myocardial infarction | True posterior myocardial infarction |
| 12 | G307. | G307. | Y202W | Acute subendocardial infarction | Acute subendocardial infarction |
| 13 | G3070 | XaAzi | YaaA4 | Acute non-Q wave infarction | Acute non-Q wave infarction |
| 14 | G3071 | XaIwY | Yali5 | Acute non-ST segment elevation myocardial infarction | Acute non-ST segment elevation myocardial infarction |
| 15 | G308. | G308. | Y202i | Inferior myocardial infarction NOS | Inferior myocardial infarction NOS |
| 16 | G309. | XaAC3 | Yah3m | Acute Q-wave infarct | Acute Q-wave infarct |
| 17 | G30A. | X202q | YabUP | Mural thrombosis | Mural thrombosis |
| 18 | G30B. | XaJX0 | YambR | Acute posterolateral myocardial infarction | Acute posterolateral myocardial infarction |
| 19 | G30X. | Gyu34 | YMAH8 | Acute transmural myocardial infarction of unspecified site | Acute transmural myocardial infarction of unspecified site |
| 20 | G30X0 | XaIwM | Yalhn | Acute ST segment elevation myocardial infarction | Acute ST segment elevation myocardial infarction |
| 21 | G30y. | G30y. | Y202S | Other acute myocardial infarction | Other acute myocardial infarction |
| 22 | G30y0 | G30y0 | Y202X | Acute atrial infarction | Acute atrial infarction |
| 23 | G30y1 | G30y1 | Y202Y | Acute papillary muscle infarction | Acute papillary muscle infarction |

| | | | | | |
|---|---|---|---|---|---|
| 24 | G30y2 | G30y2 | Y202Z | Acute septal infarction | Acute septal infarction |
| 25 | G30yz | G30yz | Y202T | Other acute myocardial infarction NOS | Other acute myocardial infarction NOS |
| 26 | G30z. | G30z. | Y202U | Acute myocardial infarction NOS | Acute myocardial infarction NOS |
| 27 | G30..-1 | X200E | Y202H | Acute myocardial infarction | Myocardial infarction |
| 28 | G30..-2 | X200E | Ya0vv | Acute myocardial infarction | Coronary thrombosis |
| 29 | G30..-3 | X200e | Y203J | Acute myocardial infarction | Cardiac rupture after acute myocardial infarction |
| 30 | G30..-4 | X200E | Ya0rZ | Acute myocardial infarction | Heart attack |
| 31 | G30..-5 | XE0Uh | Y202N | Acute myocardial infarction | Acute myocardial infarction |
| 32 | G30..-6 | X203v | YM1kK | Acute myocardial infarction | Thrombosis - coronary |
| 33 | G30..-7 | X200a | Y2034 | Acute myocardial infarction | Silent myocardial infarction |
| 34 | G31.. | G31.. | Y201b | Other acute and subacute ischaemic heart disease | Other acute and subacute ischaemic heart disease |
| 35 | G310. | G310. | Y20CX | Postmyocardial infarction syndrome | Post-myocardial infarction syndrome |
| 36 | G311. | XE0Ui | Y2021 | Preinfarction syndrome | Worsening angina |
| 37 | G311. | XE0Ui | Y2021 | Preinfarction syndrome | Worsening angina |
| 38 | G311.-1 | XE0Ui | Y2023 | Preinfarction syndrome | Crescendo angina |
| 39 | G311.-2 | XE0Ui | Y2024 | Preinfarction syndrome | Impending infarction |
| 40 | G311.-3 | X2009 | Y201z | Preinfarction syndrome | Unstable angina |
| 41 | G311.-4 | X2007 | Y201u | Preinfarction syndrome | Angina at rest |
| 42 | G3110 | G3110 | Y2035 | Myocardial infarction aborted | Aborted myocardial infarction |
| 43 | G3110-1 | G3110 | Y2035 | Myocardial infarction aborted | Aborted myocardial infarction |
| 44 | G3111 | X2009 | Y201z | Unstable angina | Unstable angina |
| 45 | G3112 | X2007 | Y201u | Angina at rest | Angina at rest |
| 46 | G3113 | XaFsG | Yah38 | Refractory angina | Refractory angina |

| | | | | | |
|---|---|---|---|---|---|
| **47** | **G3114** | **XE0Ui** | **Y2021** | **Worsening angina** | **Worsening angina** |
| **48** | **G3115** | **XaINF** | **YakmH** | **Acute coronary syndrome** | **Acute coronary syndrome** |
| **49** | **G311z** | **G311z** | **Y2026** | **Preinfarction syndrome NOS** | **Preinfarction syndrome NOS** |
| **50** | **G312.** | **G312.** | **YMAkD** | **Coronary thrombosis not resulting in myocardial infarction** | **Coronary thrombosis not resulting in myocardial infarction** |
| **51** | **G31y.** | **G31..** | **Y201b** | **Other acute and subacute ischaemic heart disease** | **Other acute and subacute ischaemic heart disease** |
| **52** | **G31y0** | **G31y0** | **Y2022** | **Acute coronary insufficiency** | **Acute coronary insufficiency** |
| **53** | **G31y1** | **G31y1** | **Y201d** | **Microinfarction of heart** | **Microinfarction of heart** |
| **54** | **G31y2** | **G31y2** | **Y202F** | **Subendocardial ischaemia** | **Subendocardial ischaemia** |
| **55** | **G31y3** | **XaFsH** | **Yah39** | **Transient myocardial ischaemia** | **Transient myocardial ischaemia** |
| **56** | **G31yz** | **G31yz** | **Y201e** | **Other acute and subacute ischaemic heart disease NOS** | **Other acute and subacute ischaemic heart disease NOS** |
| **57** | **G32..** | **XE2aA** | **Y202y** | **Old myocardial infarction** | **Old myocardial infarction** |
| **58** | **G32..-1** | **XE2aA** | **Y202z** | **Old myocardial infarction** | **Healed myocardial infarction** |
| **59** | **G32..-2** | **XaQk7** | **YatIS** | **Old myocardial infarction** | **History of myocardial infarction** |
| **60** | **G33..** | **G33..** | **Y201n** | **Angina pectoris** | **Angina pectoris** |
| **61** | **G330.** | **G330.** | **Y201w** | **Angina decubitus** | **Angina decubitus** |
| **62** | **G3300** | **G3300** | **Y201v** | **Nocturnal angina** | **Nocturnal angina** |
| **63** | **G330z** | **G330z** | **Y201x** | **Angina decubitus NOS** | **Angina decubitus NOS** |
| **64** | **G331.** | **G331.** | **Y202A** | **Prinzmetal's angina** | **Prinzmetal's angina** |
| **65** | **G331.-1** | **G331.** | **Y202A** | **Prinzmetal's angina** | **Prinzmetal's angina** |
| **66** | **G332.** | **X200B** | **Y2028** | **Coronary artery spasm** | **Coronary artery spasm** |
| **67** | **G33z.** | **G33z.** | **Y201p** | **Angina pectoris NOS** | **Angina pectoris NOS** |
| **68** | **G33z0** | **G33z0** | **Y201q** | **Status anginosus** | **Status anginosus** |
| **69** | **G33z1** | **G33z1** | **Y201r** | **Stenocardia** | **Stenocardia** |

| | | | | | |
|---|---|---|---|---|---|
| 70 | G33z2 | G33z2 | Y201s | Syncope anginosa | Syncope anginosa |
| 71 | G33z3 | Xa7nH | YaYAM | Angina on effort | Angina on effort |
| 72 | G33z4 | Ua1eH | YMFz7 | Ischaemic chest pain | Ischaemic chest pain |
| 73 | G33z5 | XaEXt | Yaf3g | Post infarct angina | Post infarct angina |
| 74 | G33z6 | X200A | Y2020 | New onset angina | New onset angina |
| 75 | G33z7 | X2008 | Y201y | Stable angina | Stable angina |
| 76 | G33zz | G33z. | Y201p | Angina pectoris NOS | Angina pectoris NOS |
| 77 | G34.. | G34.. | Y201f | Other chronic ischaemic heart disease | Other chronic ischaemic heart disease |
| 78 | G340. | XM0rN | Y201U | Coronary atherosclerosis | Coronary atherosclerosis |
| 79 | G3400 | G3400 | YaYAN | Single coronary vessel disease | Single coronary vessel disease |
| 80 | G3401 | G3401 | YaYAO | Double coronary vessel disease | Double coronary vessel disease |
| 81 | G340.-1 | X2006 | Y201g | Coronary atherosclerosis | Triple vessel disease of the heart |
| 82 | G340.-2 | XE2uV | Y201Z | Coronary atherosclerosis | Coronary artery disease |
| 83 | G342. | G70.. | YMAjY | Atherosclerotic cardiovascular disease | Atherosclerotic cardiovascular disease |
| 84 | G343. | G343. | YMAkP | Ischaemic cardiomyopathy | Ischaemic cardiomyopathy |
| 85 | G344. | X200D | Y202G | Silent myocardial ischaemia | Silent myocardial ischaemia |
| 86 | G34y. | G34y. | Y201h | Other specified chronic ischaemic heart disease | Other specified chronic ischaemic heart disease |
| 87 | G34y0 | G34y1 | Y202E | Chronic coronary insufficiency | Chronic coronary insufficiency |
| 88 | G34y1 | G34y1 | Y202D | Chronic myocardial ischaemia | Chronic myocardial ischaemia |
| 89 | G34yz | G34yz | Y201i | Other specified chronic ischaemic heart disease NOS | Other specified chronic ischaemic heart disease NOS |
| 90 | G34z. | G34z. | Y201j | Other chronic ischaemic heart disease NOS | Other chronic ischaemic heart disease NOS |
| 91 | G34z0 | XaG1Q | YahGt | Asymptomatic coronary heart disease | Asymptomatic coronary heart disease |
| 92 | G35.. | G35.. | YMAk7 | Subsequent myocardial infarction | Subsequent myocardial infarction |

| | | | | | |
|---|---|---|---|---|---|
| 93 | G350. | G350. | YMAk8 | Subsequent myocardial infarction of anterior wall | Subsequent myocardial infarction of anterior wall |
| 94 | G351. | G351. | YMAk9 | Subsequent myocardial infarction of inferior wall | Subsequent myocardial infarction of inferior wall |
| 95 | G353. | G353. | Ya3p0 | Subsequent myocardial infarction of other sites | Subsequent myocardial infarction of other sites |
| 96 | G35X. | Gyu36 | YMAHA | Subsequent myocardial infarction of unspecified site | Subsequent myocardial infarction of unspecified site |
| 97 | G36.. | G36.. | YMAkA | Certain current complications following acute myocardial infarction | Certain current complications following acute myocardial infarction |
| 98 | G360. | G360. | YMAkB | Haemopericardium as current complication following acute myocardial infarction | Haemopericardium as current complication following acute myocardial infarction |
| 99 | G361. | G361. | YMAkC | Atrial septal defect as current complication following acute myocardial infarction | Atrial septal defect as current complication following acute myocardial infarction |
| 100 | G362. | X200d | YMAkE | Ventricular septal defect as current complication following acute myocardial infarction | Ventricular septal defect as current complication following acute myocardial |
| 101 | G363. | G363. | YMAkF | Rupture of cardiac wall without haemopericardium as current complication following acute myocardial infarction | Rupture of cardiac wall without haemopericardium as current complication following acute myocardial infarction |
| 102 | G364. | G364. | YMAzL | Rupture of chordae tendinae as current complication following acute myocardial infarction | Rupture of chordae tendinae as current complication following acute myocardial infarction |
| 103 | G365. | G365. | YMAzM | Rupture of papillary muscle as current complication following acute myocardial infarction | Rupture of papillary muscle as current complication following acute myocardial infarction |
| 104 | G366. | G366. | YMAkT | Thrombosis of atrium, auricular appendage, and ventricle as current complications following acute myocardial infarction | Thrombosis of atrium, auricular appendage, and ventricle as current complications following acute myocardial infarction |
| 105 | G38.. | XaD2b | Yad42 | Postoperative myocardial infarction | Postoperative myocardial infarction |
| 106 | G380. | XaD2d | Yad46 | Postoperative transmural myocardial infarction of anterior wall | Postoperative transmural myocardial infarction of anterior wall |
| 107 | G381. | XaD2e | Yad47 | Postoperative transmural myocardial infarction of inferior wall | Postoperative transmural myocardial infarction of inferior wall |

| | | | | | |
|---|---|---|---|---|---|
| 108 | G382. | XaD2f | Yad48 | Postoperative transmural myocardial infarction of other sites | Postoperative transmural myocardial infarction of other sites |
| 109 | G383. | XaD2g | Yad49 | Postoperative transmural myocardial infarction of unspecified site | Postoperative transmural myocardial infarction of unspecified site |
| 110 | G384. | XaD2h | Yad4A | Postoperative subendocardial myocardial infarction | Postoperative subendocardial myocardial infarction |
| 111 | G38z. | XaD2i | Yad4B | Postoperative myocardial infarction, unspecified | Postoperative myocardial infarction, unspecified |
| 112 | G3y.. | G3y.. | Y201k | Other specified ischaemic heart disease | Other specified ischaemic heart disease |
| 113 | G3z.. | G3z.. | Y201l | Ischaemic heart disease NOS | Ischaemic heart disease NOS |
| 114 | G3…-1 | XE2uV | Y201T | Ischaemic heart disease | Ischaemic heart disease |
| 115 | G3…-2 | XE2uV | Y201Y | Ischaemic heart disease | Atherosclerotic heart disease |
| 116 | G3…-3 | XE2uV | Y201W | Ischaemic heart disease | IHD - Ischaemic heart disease |
| 117 | G501. | X201u | YadYu | Post infarction pericarditis | Post-infarction pericarditis |
| 118 | G64.. | XE0VI | Y20FP | Cerebral arterial occlusion | Cerebral arterial occlusion |
| 119 | G640. | G640. | Y00cj | Cerebral thrombosis | Cerebral thrombosis |
| 120 | G6400 | G6400 | YMAiF | Cerebral infarction due to thrombosis of cerebral arteries | Cerebral infarction due to thrombosis of cerebral arteries |
| 121 | G641. | G641. | Y20G2 | Cerebral embolism | Cerebral embolism |
| 122 | G6410 | G6410 | YMAiG | Cerebral infarction due to embolism of cerebral arteries | Cerebral infarction due to embolism of cerebral arteries |
| 123 | G641.-1 | G641. | Y20G1 | Cerebral embolism | Cerebral embolus |
| 124 | G64z. | XE0VJ | Y00cn | Cerebral infarction NOS | Cerebral infarction NOS |
| 125 | G64z0 | Xa00K | Ya0re | Brainstem infarction | Brainstem infarction |
| 126 | G64z1 | Xa00M | Ya0rg | Wallenberg syndrome | Wallenberg syndrome |
| 127 | G64z1-1 | Xa00M | Ya3VD | Wallenberg syndrome | Lateral medullary syndrome |
| 128 | G64z2 | XaBEC | YaaWG | Left sided cerebral infarction | Left sided cerebral infarction |
| 129 | G64z3 | XaBED | YaaWH | Right sided cerebral infarction | Right sided cerebral infarction |

| | | | | | |
|---|---|---|---|---|---|
| **130** | **G64z4** | **XaJgQ** | **YamlC** | **Infarction of basal ganglia** | **Infarction of basal ganglia** |
| **131** | **G64z.-1** | **X00D9** | **Y00d0** | **Cerebral infarction NOS** | **Brainstem infarction NOS** |
| **132** | **G64z.-2** | **Xa00J** | **Ya0rd** | **Cerebral infarction NOS** | **Cerebellar infarction** |
| **133** | **G64..-1** | **X00D3** | **Y00co** | **Cerebral arterial occlusion** | **CVA - cerebrovascular accident due to cerebral artery occlusion** |
| **134** | **G64..-2** | **Xa0kZ** | **Y00ci** | **Cerebral arterial occlusion** | **Cerebral infarction** |
| **135** | **G64..-3** | **G640.** | **Y00cj** | **Cerebral arterial occlusion** | **Cerebral thrombosis** |
| **136** | **G65..** | **XE0VK** | **Y00ds** | **Transient cerebral ischaemia** | **Transient cerebral ischaemia** |
| **137** | **G650.** | **G650.** | **Y00e3** | **Basilar artery syndrome** | **Basilar artery syndrome** |
| **138** | **G650.-1** | **G650.** | **Y00e4** | **Basilar artery syndrome** | **Insufficiency - basilar artery** |
| **139** | **G652.** | **G652.** | **Y00e6** | **Subclavian steal syndrome** | **Subclavian steal syndrome** |
| **140** | **G653.** | **G653.** | **YMAiN** | **Carotid artery syndrome hemispheric** | **Carotid artery syndrome hemispheric** |
| **141** | **G654.** | **G654.** | **YMAiQ** | **Multiple and bilateral precerebral artery syndromes** | **Multiple and bilateral precerebral artery syndromes** |
| **142** | **G656.** | **X00DW** | **Y00e5** | **Vertebrobasilar insufficiency** | **Vertebrobasilar insufficiency** |
| **143** | **G65y.** | **G65y.** | **Y00dv** | **Other transient cerebral ischaemia** | **Other transient cerebral ischaemia** |
| **144** | **G65z.** | **G65z.** | **Y00du** | **Transient cerebral ischaemia NOS** | **Transient cerebral ischaemia NOS** |
| **145** | **G65z1** | **G65z1** | **Y00cZ** | **Intermittent cerebral ischaemia** | **Intermittent cerebral ischaemia** |
| **146** | **G65zz** | **G65z.** | **Y00du** | **Transient cerebral ischaemia NOS** | **Transient cerebral ischaemia NOS** |
| **147** | **G65..-1** | **Xa00T** | **Ya0ry** | **Transient cerebral ischaemia** | **Drop attack** |
| **148** | **G65..-2** | **XE0VK** | **Y00dq** | **Transient cerebral ischaemia** | **Transient ischaemic attack** |
| **149** | **G65..-3** | **X00DW** | **Y00e5** | **Transient cerebral ischaemia** | **Vertebrobasilar insufficiency** |
| **150** | **G66..** | **XE2aB** | **Y00ch** | **Stroke and cerebrovascular accident unspecified** | **Stroke and cerebrovascular accident unspecified** |
| **151** | **G667.** | **G667.** | **YaYAW** | **Left sided CVA** | **Left sided CVA** |
| **152** | **G668.** | **G668.** | **YaYAX** | **Right sided CVA** | **Right sided CVA** |

| | | | | | |
|---|---|---|---|---|---|
| 153 | G66..-1 | XE2aB | Y00ch | Stroke and cerebrovascular accident unspecified | Stroke and cerebrovascular accident unspecified |
| 154 | G66..-2 | G66.. | YaeBs | Stroke and cerebrovascular accident unspecified | Stroke unspecified |
| 155 | G66..-3 | G66.. | YabW4 | Stroke and cerebrovascular accident unspecified | CVA - Cerebrovascular accident unspecified |
| 156 | Gyu34 | Gyu34 | YMAH8 | Acute transmural myocardial infarction of unspecified site | Acute transmural myocardial infarction of unspecified site |
| 157 | F4236 | F4236 | Y00dy | Amaurosis fugax | Amaurosis fugax |
| 158 | Fyu55 | Fyu55 | YMADV | Other transient cerebral ischaemic attacks and related syndromes | Other transient cerebral ischaemic attacks and related syndromes |
| 159 | G63y0 | G63y0 | YMAiD | Cerebral infarct due to thrombosis of precerebral arteries | Cerebral infarct due to thrombosis of precerebral arteries |
| 160 | G63y1 | G63y1 | YMAiE | Cerebral infarction due to embolism of precerebral arteries | Cerebral infarction due to embolism of precerebral arteries |
| 161 | G6760 | G6760 | YadUZ | Cerebral infarction due to cerebral venous thrombosis, nonpyogenic | Cerebral infarction due to cerebral venous thrombosis, nonpyogenic |
| 162 | G6W.. | Gyu6G | YMAIB | Cerebral infarction due to unspecified occlusion or stenosis of precerebral arteries | Cerebral infarction due to unspecified occlusion or stenosis of precerebral arteries |
| 163 | G6X.. | Gyu63 | YMAHy | Cerebral infarction due to unspecified occlusion or stenosis of cerebral arteries | Cerebral infarction due to unspecified occlusion or stenosis of cerebral arteries |
| 164 | Gyu63 | Gyu63 | YMAHy | Cerebral infarction due to unspecified occlusion or stenosis of cerebral arteries | Cerebral infarction due to unspecified occlusion or stenosis of cerebral arteries |
| 165 | Gyu64 | Gyu64 | YMAHz | Other cerebral infarction | Other cerebral infarction |
| 166 | Gyu65 | Gyu65 | YMAI0 | Occlusion and stenosis of other precerebral arteries | Occlusion and stenosis of other precerebral arteries |
| 167 | Gyu66 | Gyu66 | YMAI1 | Occlusion and stenosis of other cerebral arteries | Occlusion and stenosis of other cerebral arteries |
| 168 | ZV12D | XaX16 | YatuJ | [V]Personal history of transient ischaemic attack | [V]Personal history of transient ischaemic attack |
| 169 | 14A6. | 14A6. | Ya04n | H/0: Heart failure | H/0: Heart failure |
| 170 | G580.. | XEV8 | Y20Bq | Heart failure | Heart failure |

**Supplementary Table 3 Definition of QRISK3risk factors included in Cox clinical risk scores, within the UK Biobank**

| Risk factor | UKBB data field | Hospital episode statistics reported ICD-10 codes with date of event/visit | Hospital episode statistics reported ICD-9 codes with date of event/visit |
|---|---|---|---|
| Age | 34,52 | - | - |
| Sex | 31 | - | - |
| BMI | 21001 | - | - |
| HDL ratio | 30760, 30690,30780 | - | - |
| Townsend deprivation index | 189 | - | - |
| Ethnicity | 21000 | - | - |
| Smoker status | 20116 | - | |
| Systolic blood pressure | 93,4080 | - | - |
| Atrial Fibrillation | 131350 | I48 | - |
| Diabetes type 1 and 2 | 2443 | E10, O230, E11, O231 | 25001, 25011, 25021, 25031, 25041, 25051, 25061, 25071, 25081, 25091, 25003, 25013, 25023, 25033, 25043, 25053, 25063, 25073, 25083, 25093<br>25000, 25010, 25020, 25030, 25040, 25050, 25060, 25070, 25080, 25090, 25002, 25012, 25022, 25032, 25042, 25052, 25062, 25072, 25082, 25092 |
| Primary hyper-tension | 131287 | I10 | - |
| HDL ratio | 30764 | - | - |
| Previous Corticosteroid use | - | Z7951, Z7952 | - |
| Migraine | - | G43, G440, N943 | 346 |
| Prevalent Rheumatoid arthritis | - | M05, M06 | 714 |
| Systolic blood pressure standard deviation | 93,4080 | - | - |
| Severe mental illness | | F03, F068, F09, F20, F22, F23, F259, F28, F29, F31, F39, F53, F333 | 295, 298, 296 |

**Supplementary Table 4 Cardiovascular medication, with accompanying Anatomical Therapeutic Chemical (ATC) codes**

| ATC code | Description |
|---|---|
| C08D | Calcium channel Blocker |
| C01BD01 | Anti-arrhythmic class: I/III |
| C01AA05 | Cardiac Glycosides |
| C07AA02 | Beta blocking agents (non-selective) |
| C07AA03 | Beta blocking agents (non-selective) |
| C07AA05Â | Beta blocking agents(non-selective) |
| C07AA06Â | Beta blocking agents (non-selective) |
| C07AA07Â | Beta blocking agents (non-selective) |
| C07AA12Â | Beta blocking agents (non-selective) |
| C07AA15Â | Beta blocking agents (non-selective) |
| C07AA23Â | Beta blocking agents (non-selective) |
| C07AB02Â | Beta blocking agents (Selective) |
| C07AB03Â | Beta blocking agents (Selective) |
| C07AB04Â | Beta blocking agents (Selective) |
| C07AB05Â | Beta blocking agents (Selective) |
| C07AB07Â | Beta blocking agents (Selective) |
| C07AB08Â | Beta blocking agents (Selective) |
| C07AB12Â | Beta blocking agents (Selective) |
| C07AG01Â | Alpha/Beta blocking agents |
| C07AG02Â | Alpha/Beta blocking agents |

**Supplementary table 5** : **Minimum sample size calculation for Cox clinical + ECG and risk scores**

| Model | # Model parameters | Event rate | Estimate Cox-snell R-squared | Minimum EPP | # Minimum MACCE | Minimum Sample size |
|---|---|---|---|---|---|---|
| **Cox clinical** | | | | | | |
| Male | 32 | 0.011 | 0.056 | 19.95 | 639 | 4,980 |
| Female | 32 | 0.005 | 0.04 | 14.31 | 430 | 6,619 |
| **Cox clinical + ECG** | | | | | | |
| Male | 71 | 0.011 | 0.056 | 19.94 | 1,417 | 11,049 |
| Female | 71 | 0.005 | 0.04 | 14.31 | 1,017 | 15,665 |

Abbreviations: EPP = Event per parameter, MACCE = Major Adverse Cardiovascular & Cerebrovascular Event

**Supplementary Table 6 Summary of ECG parameters measured during the final 15-seconds of exercise during a sub-maximal cycle ergometry test, corresponding to peak exercise intensity**

| ECG parameter | Male (N=17,376) | Female (N = 18,221) | Overall (N=35,597) |
|---|---|---|---|
| PR interval(ms) | 152.4±22.3 | 144.5±19.4 | 148·5·1±21·3 |
| QRS interval(ms) | 96.1±9.5 | 93.4±7.7 | 94·8±8·7 |
| QTc interval(ms) | 398.1±25.4 | 387.2±23.8 | 397·7±24·6 |
| RR interval: (ms) | 561.2±69.1 | 538.7±61.3 | 550·2±66·4 |
| PVC count | 0.1±0.6 | 0.1±0.5 | 0·1±0·6 |
| CosEn | -2.9±0.4 | -2.9±0.4 | -2·9±0·4 |
| Irregularity evidence | 0.7±2.2 | 0.8±2.2 | 0·8±2·2 |
| Regularity evidence | 7.0±2.1 | 7.4±2.1 | 7·2±2·1 |
| Density evidence | 0.1±0.9 | 0.1±0.8 | 0·1±0·8 |
| Anisotropy evidence | 0.4±1.6 | 0.5±1.6 | 0·5±1·6 |
| PAC evidence | 0.0±0.5 | 0.0±0.4 | >0·1±0·5 |
| Atrial fibrillation evidence | -18.5±9.2 | -19.3±9.3 | -18·9±9·2 |
| Atrial tachycardia evidence | 8.1±3.9 | 8.6±3.8 | 8·4±3·9 |

Data are shown as means±SD

Data are reported on 35,597 participants with measurable ECG parameters, 3,463 (9·7%) MACCE were reported over 12.5 years of follow-up. 95% confidence intervals were obtained via 1000 bootstrap iterations.

*ECG measurements were recorded from the final 15 seconds of rest, exercise, recovery*

All ECG intervals (PR, QRS, QTc, RR) are reported in milliseconds. Premature ventricular contraction (PVC) represents counts of ectopic beats observed within the 15-second resting ECG segment. CosEn is a dimensionless score presenting ECG signal entropy. Evidence-based indices (irregularity, regularity, density, anisotropy) count the number of ΔRR-intervals across pre-defined bins from a Poincare representation over the 15-second ECG recording. Atrial fibrillation, and atrial tachycardia evidence are dimensionless scores derived from the RR-interval series and Poincaré-plot representation, with higher values indicating greater rhythm irregularity or stronger evidence for the respective arrhythmia patterns.

**Supplementary Table 7 Summary of ECG parameters measured during 15-seconds of recovery following a sub-maximal cycle ergometry test**

| ECG parameter | Male (N=17,376) | Female (N = 18,221) | Overall (N=35,597) |
|---|---|---|---|
| PR interval(ms) | 152.4±19.8 | 146.2±17.4 | 156·5±20·6 |
| QRS interval(ms) | 95.5±9.9 | 92.9±8.0 | 93·0±10·9 |
| QTc interval(ms) | 389.4±17.8 | 393.0±16.9 | 420·2±21·1 |
| RR interval: (ms) | 754.6±137.8 | 731.8±116.0 | 874·8±136·7 |
| PVC count | 0.1±0.8 | 0.1±0.5 | 0·1±0·4 |
| CosEn | -2.4±0.5 | -2.3±0.5 | -2·5±0·5 |
| Irregularity evidence | 1.1±2.4 | 1.0±2.2 | 0·9±1·9 |
| Regularity evidence | 2.7±2.0 | 2.9±1.9 | 2·0±1·5 |
| Density evidence | 0.1±0.5 | 0.1±0.5 | 0·1±0·4 |
| Anisotropy evidence | 0.5±1.4 | 0.4±1.3 | 0·3±1·1 |
| PAC evidence | 0.0±0.4 | 0.0±0.4 | >0·1±0·3 |
| Atrial fibrillation evidence | -8.0±8.4 | -7.9±8.2 | -5·6±6·4 |
| Atrial tachycardia evidence | 4.2±3.7 | 4.2±3.5 | 3·2±3·0 |

Data are shown as means±SD

Data are reported on 35,597 participants with measurable ECG parameters, 3,463 (9·7%) MACCE were reported over 12.5 years of follow-up.95% confidence intervals were obtained via 1000 bootstrap iterations.

*ECG measurements were recorded from the final 15 seconds of rest, exercise, recovery*

All ECG intervals (PR, QRS, QTc, RR) are reported in milliseconds. Premature ventricular contraction (PVC) represents counts of ectopic beats observed within the 15-second resting ECG segment. CosEn is a dimensionless score presenting ECG signal entropy. Evidence-based indices (irregularity, regularity, density, anisotropy) count the number of ΔRR-intervals across pre-defined bins from a Poincare representation over the 15-second ECG recording. Atrial fibrillation, and atrial tachycardia evidence are dimensionless scores derived from the RR-interval series and Poincaré-plot representation, with higher values indicating greater rhythm irregularity or stronger evidence for the respective arrhythmia patterns.

**Supplementary Table 8 Univariate performance of QRISK3 risk factors ECG parameters measured during rest exercise and recovery**

| ECG parameters | C-index (95% CI) | Hazard ratio (95% CI) |
|---|---|---|
| **Resting ECG** | | |
| PR interval(ms) | 0.53 (0.52 - 0.54) | 1.08 (1.04 - 1.12) |
| QRS interval(ms) | 0.53 (0.52 - 0.54) | 1.07 (1.04 - 1.11) |
| QTc interval(ms) | 0.51 (0.5 - 0.52) | 0.98 (0.94 - 1.02) |
| RR interval: (ms) | 0.5 (0.49 - 0.51) | 1.02 (0.98 - 1.06) |
| PVC count | 0.51 (0.5 - 0.52) | 1.02 (1 - 1.04) |
| CosEn | 0.56 (0.55 - 0.57) | 0.94 (0.9 - 0.98) |
| Irregularity evidence | 0.5 (0.49 - 0.51) | 1.06 (1.02 - 1.1) |
| Regularity evidence | 0.54 (0.53 - 0.55) | 1.07 (1.03 - 1.11) |
| Atrial fibrillation evidence | 0.54 (0.53 - 0.55) | 0.96 (0.92 – 1.00) |
| Atrial tachycardia evidence | 0.54 (0.52 - 0.55) | 1.07 (1.03 - 1.11) |
| Antisotropy evidence | 0.51 (0.49 - 0.52) | 1.04 (1.01 - 1.07) |
| Density evidence | 0.5 (0.49 - 0.51) | 0.98 (0.94 - 1.02) |
| **Exercise ECG** | | |
| PR interval(ms) | 0.54 (0.53 - 0.55) | 1.08 (1.04 - 1.12) |
| QRS interval(ms) | 0.53 (0.53 - 0.55) | 1.1 (1.06 - 1.13) |
| QTc interval(ms) | 0.54 (0.53 - 0.55) | 1.07 (1.04 - 1.1) |
| RR interval: (ms) | 0.58 (0.57 - 0.6) | 1.2 (1.16 - 1.24) |
| PVC count | 0.5 (0.49 - 0.51) | 1.05 (1.02 - 1.08) |
| CosEn | 0.53 (0.52 - 0.55) | 0.96 (0.92 - 1) |
| Irregularity evidence | 0.52 (0.5 - 0.52) | 1.04 (1 - 1.07) |
| Regularity evidence | 0.53 (0.52 - 0.55 | 0.92 (0.89 - 0.96) |
| Atrial fibrillation evidence | 0.55 (0.54 - 0.56) | 1.08 (1.04 - 1.12) |
| Atrial tachycardia evidence | 0.55 (0.54 - 0.56) | 1 (0.96 - 1.04) |
| Anisotropy evidence | 0.51 (0.5 - 0.52) | 1.03 (1 - 1.06) |
| Density evidence | 0.5 (0.49 - 0.51) | 1.02 (0.99 - 1.05) |
| **Recovery ECG** | | |
| PR interval(ms) | 0.54 (0.53 - 0.56) | 1.1 (1.06 - 1.14) |
| QRS interval(ms) | 0.53 (0.52 - 0.55) | 1.1 (1.07 - 1.14) |
| QTc interval(ms) | 0.55 (0.54 - 0.56) | 1.09 (1.05 - 1.13) |
| RR interval: (ms) | 0.51 (0.5 - 0.52) | 1.04 (0.99 - 1.08) |
| PVC count | 0.51 (0.5 - 0.52) | 1.02 (0.99 - 1.04) |
| CosEn | 0.57 (0.56 - 0.58) | 0.93 (0.89 - 0.98) |
| Irregularity evidence | 0.5 (0.49 - 0.51) | 1.05 (1.01 - 1.09) |
| Regularity evidence | 0.53 (0.53 - 0.55) | 1.04 (1 - 1.08) |
| Atrial fibrillation evidence | 0.53 (0.52 - 0.54) | 0.98 (0.94 - 1.02) |
| Atrial tachycardia evidence | 0.54 (0.53 - 0.55) | 1.07 (1.04 - 1.11) |
| Anisotropy evidence | 0.51 (0.5 - 0.53) | 1.05 (1.02 - 1.08) |
| Density evidence | 0.5 (0.49 - 0.51) | 1.01 (0.97 - 1.04) |

| **QRISK3 risk factor** | | |
|---|---|---|
| Age | 0.66 (0.65 - 0.67) | 1.95 (1.86 - 2.05) |
| Body mass index | 0.57 (0.56 - 0.58) | 1.27 (1.23 - 1.31) |
| Ethnicity | 0.51 (0.5 - 0.51) | 1.16 (0.98 - 1.36) |
| Townsend deprivation index | 0.5 (0.49 - 0.51 | 1.03 (0.99 - 1.07) |
| Current smoker | 0.51 (0.51 - 0.52) | 1.7 (1.5 - 1.94) |
| Former smoker | 0.52 (0.51 - 0.53) | 1.17 (1.08 - 1.27) |
| Systolic blood pressure | 0.6 (0.59 - 0.61) | 1.19 (1.14 - 1.23) |
| Type 1 diabetes | 0.5 (0.5 - 0.5) | 2.15 (0.97 - 3.5) |
| Type 2 diabetes | 0.51 (0.51 - 0.51) | 2.41 (1.89 - 3.03) |
| HDL: ratio | 0.56 (0.56 - 0.57) | 1.27 (1.22 - 1.31) |
| Hypertension | 0.53 (0.53 - 0.54) | 1.92 (1.69 - 2.17) |
| Rheumatoid arthritis | 0.5 (0.5 - 0.5) | 1.34 (0.54 - 2.26) |
| Severe mental illness | 0.5 (0.5 - 0.5) | 1.2 (0.3 - 2.41) |

Data are reported on 35,597 participants with measurable ECG parameters, 2,591 (7·27%) MACCE were reported within 10-years. 95% confidence intervals were derived via bootstrapping for 1000 iterations

Each row corresponds to a separate univariate Cox model in which each listed ECG risk factors was fitted individually.

Hazard ratios are presented per Standard deviation increase for continuous risk factors.

*ECG measurements were recorded from the final 15 seconds of rest, exercise, recovery*

All ECG intervals (PR, QRS, QTc, RR) are reported in milliseconds. Premature ventricular contraction (PVC) represents counts of ectopic beats observed within the 15-second resting ECG segment. CosEn is a dimensionless score presenting ECG signal entropy. Evidence-based indices (irregularity, regularity, density, anisotropy) count the number of ΔRR-intervals across pre-defined bins from a Poincare representation over the 15-second ECG recording. Atrial fibrillation, and atrial tachycardia evidence are dimensionless scores derived from the RR-interval series and Poincaré-plot representation, with higher values indicating greater rhythm irregularity or stronger evidence for the respective arrhythmia patterns.

**Supplementary Table 9 Reclassification performance for 10-year MACCE risk prediction after adding deep learning–derived ECG risk scores to Cox clinical risk scores in men and women at a 10% risk threshold. ECG measurements were recorded during the first 15 seconds of rest, the final 15 seconds of exercise, the final 15 seconds of recovery, and the complete 7-minute 15-second submaximal exercise ECG recording**

| Model | Comparator | Net-benefit (95% CI) | Categorical NRI (95% CI) | Continuous NRI (95% CI) |
|---|---|---|---|---|
| **Male** | | | | |
| Cox clinical | Cox clinical+ Resting ECG | -0.005(-0.012 - 0.002) | * -0.012 ( -0.020 - -0.003) | * 0.065 (0.019 - 0.110) |
| Cox clinical | Cox clinical+ Exercise ECG | *-0.01(-0.018 - -0.002) | *-0.010 ( -0.018 - -0.001) | *0.053 (0.008 - 0.098) |
| Cox clinical | Cox clinical+ Recovery ECG | 0(-0.008 - 0.008) | -0.001 ( -0.008 - 0.007) | **0.090 (0.045 - 0.135) |
| Cox clinical | Cox clinical+ Complete ECG | -0.001(-0.01 - 0.008) | -0.001 ( -0.010 - 0.009) | **0.108 (0.063 - 0.152) |
| **Female** | | | | |
| Cox clinical | Cox clinical+ Resting ECG | -0.001(-0.006 - 0.004) | -0.001 ( -0.006 - 0.005) | 0.034 ( -0.031 - 0.098) |
| Cox clinical | Cox clinical+ Exercise ECG | 0.005(-0.007 - 0.017) | 0.004 ( -0.007 - 0.016_ | 0.069 (0.005 - 0.1342) |
| Cox clinical | Cox clinical+ Recovery ECG | -0.002(-0.007 - 0.005) | -0.002 ( -0.009 - 0.004) | -0.052 ( -0.117 - 0.0124) |
| Cox clinical | Cox clinical+ Complete ECG | 0.005(-0.002 - 0.014) | 0.007 ( -0.001 - 0.015) | * 0.093 (0.029 - 0.157) |

Abbreviations: NRI = Net re classification index, CI = Confidence interval
Data are reported on 41,076 participants with 3,053 (7·4%) Major Adverse Cardiovascular & Cerebrovascular events (MACCE) over 10 years. 95% confidence intervals and p– values were obtained via 1000 bootstrap iterations with a significance threshold of 0·05. **p value <0·05; ** p value < 0·001.*

Each row corresponds to a separate risk score, fitted independently with the selected risk factor and independent estimates of the baseline survival using Breslow's method, with mean-centred continuous variables, with all binary risk factors set to 0.

Clinical risk scores were derived using Cox proportional hazards regression. Extended Cox models additionally included the $ECG_{AI}$ scores as a covariate.

$ECG_{AI}$ risk scores were derived from separate deep survival networks trained on raw ECG recordings from: 15s rest, final 15s of exercise (peak intensity), final 15s recovery, and the full 7 min 15s submaximal ECG, respectively.

Cox clinical: Age FP1 + Age FP2 + BMI FP1 + BMI FP2 + SBP + SBP5 + Diabetes type 1 + Diabetes type 2 + HDL ratio + Townsend deprivation index + Smoker former + Smoker current + Smoker never + Ethnic Background + Corticosteroid + Migraine + Hypertension + Severe mental illness + Age FP1 × Townsend deprivation index + Age FP2 × Townsend deprivation index + Age FP1 × BMI FP 1 + Age FP1 × BMI FP 2 + Age FP1 × BP + Age FP1 × Diabetes type 1 + Age FP1 × Diabetes type 2 + Age FP1 × hypertension + Age FP2 × BMI FP1 + Age FP2 × BMI FP2 + Age FP2 × SBP + Age FP2 × Diabetes type1 + Age FP2 × Diabetes type2 + Age FP2 × Hypertension.

Fractional polynomials: *Age FP1 (males)* = $(age/10)^{-1}$, *Age FP2 (males)* = $(age/10)^3$, *BMI FP 1 (males)* = $(BMI/10)^{-2}$, *BMI FP 2 (males)* = $(BMI/10)^{-2}\ X\ log(BMI/10)$, *Age FP1 (Females)* = $age/10^{-2}$, *Age FP2 (Females)* = *age/10*, *BMI FP 1 (Females)* = $(BMI/10)^{-2}$, *BMI FP 2 (Females)* = $(BMI/10)^{-2}\ X\ log\ (BMI/10)$.

**Supplementary Table 10: Reclassification performance for 10-year MACCE risk prediction comparing Cox clinical and Clinical Deep Survival Network (DSN) risk scores derived from QRISK3 risk factors, evaluated at a 10% risk threshold using net benefit, categorical NRI, and continuous NRI in the overall population and by sex**

| Model | Comparator | ΔNet-benefit (95% CI) | Categorical NRI (95% CI) | Continuous NRI (95% CI) |
|---|---|---|---|---|
| **Overall** | | | | |
| Cox clinical | Clinical DSN | -0.009(-0.022 - 0.003) | -0.009 ( -0.022 - 0.004) | **0.089 (0.052 - 0.126) |
| **Male** | | | | |
| Cox clinical | Clinical DSN | *-0.017(-0.032 - -0.001) | -0.017 (-0.031- -0.002) | **0.163 (0.118 - 0.208) |
| **Female** | | | | |
| Cox clinical | Clinical DSN | 0.006(-0.02 - 0.032) | ** 0.056 (0.033 - 0.080) | **0.144 (0.081 - 0.207) |

Abbreviations: NRI = Net re classification index, DSN = Deep survival network, CI = Confidence interval
Data are reported on 41,076 participants with 3,053 (7·4%) Major Adverse Cardiovascular & Cerebrovascular events (MACCE) over 10 years. 95% confidence intervals and p– values were obtained via 1000 bootstrap iterations with a significance threshold of 0·05. **p value <0·05; ** p value < 0·001.*

Each row corresponds to a separate risk score, fitted independently with the selected risk factor and independent estimates of the baseline survival using Breslow's method, with mean-centred continuous variables, with all binary risk factors set to 0.

Clinical risk scores were derived using Cox proportional hazards regression.

Cox clinical: Age FP1 + Age FP2 + BMI FP1 + BMI FP2 + SBP + SBP5 + Diabetes type 1 + Diabetes type 2 + HDL ratio + Townsend deprivation index + Smoker former + Smoker current + Smoker never + Ethnic Background + Corticosteroid + Migraine + Hypertension + Severe mental illness + Age FP1 × Townsend deprivation index + Age FP2 × Townsend deprivation index + Age FP1 × BMI FP 1 + Age FP1 × BMI FP 2 + Age FP1 × BP + Age FP1 × Diabetes type 1 + Age FP1 × Diabetes type 2 + Age FP1 × hypertension + Age FP2 × BMI FP1 + Age FP2 × BMI FP2 + Age FP2 × SBP + Age FP2 × Diabetes type1 + Age FP2 × Diabetes type2 + Age FP2 × Hypertension.

Fractional polynomials: *Age FP1 (males) =* $(age/10)^{-1}$, *Age FP2 (males) = (age/10)³, BMI FP 1 (males) =* $(BMI/10)^{-2}$, *BMI FP 2 (males) =* $(BMI/10)^{-2}\ X\ log(BMI/10)$, *Age FP1 (Females) =* $age/10^{-2}$, *Age FP2 (Females) = age/10, BMI FP 1 (Females) =* $(BMI/10)^{-2}$, *BMI FP 2 (Females) =* $(BMI/10)^{-2}\ X\ log\ (BMI/10)$.

Clinical DSN: Age + BMI + systolic blood-pressure + SBP5 +Diabetes type 1+ Diabetes type 2 + HDL ratio + Townsend deprivation index+ Smoker former + Smoker current + Smoker never+ Ethnic Background+ Corticosteroid+ Migraine + Hypertension + Severe mental illness, Migraine, Hypertension, Severe mental illness.

**Supplementary Table 11 Reclassification performance for 10-year MACCE risk prediction comparing Cox clinical and Clinical Deep Survival Network (DSN) + ECG models at a 10% risk threshold using ECG measurements from rest, exercise, recovery, and the complete 7-minute 15-second recording, assessed using net benefit, categorical NRI, and continuous NRI**

| Model | Comparator | Net-benefit (95% CI) | Categorical NRI (95% CI) | Continuous NRI (95% CI) |
|---|---|---|---|---|
| **Male** | | | | |
| Cox clinical | Clinical DSN + Resting ECG | -0.020(-0.033 - -0.007) | *-0.020 ( -0.033 - -0.007) | ** 0.213 (0.168 - 0.258) |
| Cox clinical | Clinical DSN + Exercise ECG | *-0.049(-0.064 - -0.032) | **-0.048 ( -0.064 - -0.031) | ** 0.181 (0.136 - 0.226) |
| Cox clinical | Clinical DSN + Recovery ECG | *-0.018(-0.03 - -0.006) | *-0.018 ( -0.031 - -0.004) | **0.196 (0.151 - 0.241) |
| Cox clinical | Clinical DSN + Complete ECG | -0.013(-0.025 - 0) | -0.013 ( -0.025 - 0) | **0.268 (0.224 - 0.313) |
| **Female** | | | | |
| Cox clinical | Clinical DSN + Resting ECG | 0.011(-0.014 - 0.04) | ** 0.079 (0.055 - 0.103) | ** 0.228 (0.169 - 0.287) |
| Cox clinical | Clinical DSN + Exercise ECG | 0.014(-0.012 - 0.038) | **0.067 (0.041 - 0.092) | ** 0.172 (0.110 - 0.235) |
| Cox clinical | Clinical DSN + Recovery ECG | 0.015(-0.01 - 0.04) | **0.080 (0.055 - 0.104) | **0.218 (0.158 - 0.277) |
| Cox clinical | Clinical DSN + Complete ECG | 0.015(-0.007 - 0.037) | **0.085 (0.061 - 0.108) | ** 0.252 (0.189 - 0.314) |

Abbreviations: NRI = Net re classification index, DSN = Deep survival network, CI = Confidence interval
Data are reported on 41,076 participants with 3,053 (7·4%) Major Adverse Cardiovascular & Cerebrovascular events (MACCE) over 10 years. 95% confidence intervals and p– values were obtained via 1000 bootstrap iterations with a significance threshold of 0·05. **p value <0·05; ** p value < 0·001.*

Each row corresponds to a separate risk score, fitted independently with the selected risk factor and independent estimates of the baseline survival using Breslow's method, with mean-centred continuous variables, with all binary risk factors set to 0.

Clinical risk scores were derived using Cox proportional hazards regression. Extended Cox models additionally included the $ECG_{AI}$ scores as a covariate.

Cox clinical: Age FP1 + Age FP2 + BMI FP1 + BMI FP2 + SBP + SBP5 + Diabetes type 1 + Diabetes type 2 + HDL ratio + Townsend deprivation index + Smoker former + Smoker current + Smoker never + Ethnic Background + Corticosteroid + Migraine + Hypertension + Severe mental illness + Age FP1 × Townsend deprivation index + Age FP2 × Townsend deprivation index + Age FP1 × BMI FP 1 + Age FP1 × BMI FP 2 + Age FP1 × BP + Age FP1 × Diabetes type 1 + Age FP1 × Diabetes type 2 + Age FP1 × hypertension + Age FP2 × BMI FP1 + Age FP2 × BMI FP2 + Age FP2 × SBP + Age FP2 × Diabetes type1 + Age FP2 × Diabetes type2 + Age FP2 × Hypertension.

Fractional polynomials: *Age FP1 (males) =* $(age/10)^{-1}$, *Age FP2 (males) =* $(age/10)^3$, *BMI FP 1 (males) =* $(BMI/10)^{-2}$, *BMI FP 2 (males) =* $(BMI/10)^{-2}\ X\ log(BMI/10)$, *Age FP1 (Females) =* $age/10^{-2}$, *Age FP2 (Females) = age/10, BMI FP 1 (Females) =* $(BMI/10)^{-2}$, *BMI FP 2 (Females) =* $(BMI/10)^{-2}\ X\ log\ (BMI/10)$.

Clinical DSN: Age + BMI + systolic blood-pressure + SBP5 +Diabetes type 1+ Diabetes type 2 + HDL ratio + Townsend deprivation index+ Smoker former + Smoker current + Smoker never+ Ethnic Background+ Corticosteroid+ Migraine + Hypertension + Severe mental illness,

Clinical DSN + ECG risk scores were derived from separate deep survival networks trained on raw ECG recordings from: 15s rest, final 15s of exercise (peak intensity), final 15s recovery, and the full 7 min 15s submaximal ECG, respectively.

**Supplementary Table 12 Hazard ratio of $ECG_{AI}$ scores and conventional ECG parameters measured during rest, exercise, and recovery after excluding events occurring within the first 2 years of follow-up**

| ECG Parameter | Rest | Exercise | Recovery |
|---|---|---|---|
| $ECG_{AI}$ | **1·129 (1·072, 1·168) | **1·124 (1·068, 1·183) | **1·135 (1·089, 1·183) |
| PR interval(ms) | 1·005 (0·965, 1·046) | 0·985 (0·945, 1·026) | 1·013(0·973 ,1·054) |
| QRS interval(ms) | 1·040 (1·001, 1·080) | 1·038 (1·000, 1·076) | *1·051(1·014 ,1·089) |
| QTc interval(ms) | *1·052 (1·009, 1·097) | *1·048 (1·011, 1·086) | **1·104(1·066 ,1·144) |
| RR interval: (ms) | 0·996 (0·956,1·038) | **1·125 (1·082, 1·169) | 1·003(0·963 ,1·046) |
| PVC count | 1·020 (0·991, 1·051) | *1·045 (1·016, 1·074) | 1·012(0·980 ,1·045) |
| CosEn | 0·979 (0·937, 1·023) | 0·988 (0·949, 1·030) | 0·959(0·919 ,1·002) |
| Irregularity evidence | *1·068 (1·029, 1·108) | 1·031 (0·995, 1·069) | 1·036(0·996 ,1·077) |
| Regularity evidence | *1·074 (1·027,1·122) | 1·043(0·985, 1·104) | *1·060(1·008 ,1·115) |
| Density evidence | 0·982 (0·941, 1·024) | 1·017(0·983, 1·052) | 1·012(0·975 ,1·051) |
| Anisotropy evidence | *1·040 (1·005,1·076) | 1·025 (0·990, 1·062) | *1·043(1·008 ,1·080) |
| PAC evidence | 1·026 (0·993, 1·059) | 0·999 (0·962, 1·038) | 1·003(0·963 ,1·044) |
| Atrial fibrillation evidence | 0·985 (0·936, 1·037) | 1·029 (0·985, 1·075) | 0·974(0·923 ,1·028) |
| Atrial tachycardia evidence | **1·068 (1·031,1·107) | *1·047 (1·008, 1·086) | *1·060(1·021 ,1·100) |

Abbreviations: PVC =Premature ventricular contraction Index, CI = Confidence interval

Data are reported on 35,597 participants with measurable ECG parameters, 3,463 (9·7%) MACCE were reported over 12.5 years of follow-up

*ECG measurements were recorded from the final 15 seconds of rest, exercise, recovery*

All ECG intervals (PR, QRS, QTc, RR) are reported in milliseconds. Premature ventricular contraction (PVC) represents counts of ectopic beats observed within the 15-second resting ECG segment. CosEn is a dimensionless score presenting ECG signal entropy. Evidence-based indices (irregularity, regularity, density, anisotropy) count the number of ΔRR-intervals across pre-defined bins from a Poincare representation over the 15-second ECG recording. Atrial fibrillation, and atrial tachycardia evidence are dimensionless scores derived from the RR-interval series and Poincaré-plot representation, with higher values indicating greater rhythm irregularity or stronger evidence for the respective arrhythmia patterns.

Each row corresponds to a separate univariate Cox model in which each listed ECG risk factors was fitted individually.

Hazard ratios are presented per Standard deviation increase in selected risk factors.

95% confidence intervals and p-values were derived via bootstrapping for 1000 iterations, with a significance threshold of 0·05, following adjustment for multiple testing using the Benjamini/Hochberg procedure[260]; **p value < 0·05; ** p value < 0·001.*

**Supplementary Table 13 Hazard ratio of $ECG_{AI}$ scores and conventional ECG parameters measured during rest, exercise, and recovery after excluding events occurring within the first 5 years of follow-up**

| ECG Parameter | Rest (95% CI) | Exercise (95% CI) | Recovery (95% CI) |
|---|---|---|---|
| $ECG_{AI}$ | **1·123(1·066, 1·183) | **1·161(1·090, 1·236) | **1·108(1·053, 1·165) |
| PR interval(ms) | 1·034(0·985, 1·085) | 1·006(0·957, 1·058) | 1·034(0·985, 1·085 |
| QRS interval(ms) | 1·032(0·985, 1·081) | 1·043(0·997, 1·090) | *1·060(1·014, 1·107 |
| QTc interval(ms) | 1·042(0·989, 1·097) | *1·072(1·030, 1·115) | **1·108(1·061, 1·156) |
| RR interval: (ms) | 1·031(0·982, 1·084) | **1·151(1·098, 1·206) | 1·030(0·979, 1·083) |
| PVC count | 1·030(0·997, 1·065) | *1·057(1·024, 1·091) | 1·016(0·978, 1·055) |
| CosEn | 1·005(0·953, 1·060) | 1·006(0·958, 1·057) | 0·990(0·940, 1·044) |
| Irregularity evidence | *1·065(1·018, 1·115) | 1·037(0·993, 1·083) | 1·039(0·991, 1·090) |
| Regularity evidence | *1·063(1·007, 1·122) | 1·027(0·959, 1·101) | 1·004(0·943, 1·068) |
| Density evidence | 0·985(0·936, 1·037) | 1·008(0·965, 1·053) | 1·015(0·970, 1·062) |
| Anisotropy evidence | *1·046(1·005, 1·090) | 1·022(0·978, 1·067) | *1·055(1·013, 1·098) |
| Atrial fibrillation evidence | 0·990(0·930, 1·054) | 1·048(0·995, 1·105) | 1·002(0·939, 1·070) |
| Atrial tachycardia evidence | *1·062(1·016, 1·110) | *1·055(1·009, 1·103) | 1·048(1·001, 1·098) |

Abbreviations: PVC =Premature ventricular contraction Index, CI = Confidence interval

Data are reported on 35,597 participants with measurable ECG parameters, 3,463 (9·7%) MACCE were reported over 12.5 years of follow-up

*ECG measurements were recorded from the final 15 seconds of rest, exercise, recovery*

All ECG intervals (PR, QRS, QTc, RR) are reported in milliseconds. Premature ventricular contraction (PVC) represents counts of ectopic beats observed within the 15-second resting ECG segment. CosEn is a dimensionless score presenting ECG signal entropy. Evidence-based indices (irregularity, regularity, density, anisotropy) count the number of ΔRR-intervals across pre-defined bins from a Poincare representation over the 15-second ECG recording. Atrial fibrillation, and atrial tachycardia evidence are dimensionless scores derived from the RR-interval series and Poincaré-plot representation, with higher values indicating greater rhythm irregularity or stronger evidence for the respective arrhythmia patterns.

Each row corresponds to a separate univariate Cox model in which each listed ECG risk factors was fitted individually.

Hazard ratios are presented per Standard deviation increase in selected risk factors.

95% confidence intervals and p-values were derived via bootstrapping for 1000 iterations, with a significance threshold of 0·05, following adjustment for multiple testing using the Benjamini/Hochberg procedure[260]; **p value <0·05; ** p value < 0·001.*

#### Supplementary Table 14: Reclassification performance for 10-year MACCE risk prediction at a 10% risk threshold among participants aged ≤45 years comparing Cox clinical and Clinical Deep Survival Network (DSN) + ECG models derived from rest, exercise, recovery, and the complete submaximal exercise ECG recording

| Model | Comparator | Net-benefit (95% CI) | Categorical NRI (95% CI) | Continuous NRI (95% CI) |
|---|---|---|---|---|
| **Overall** | | | | |
| Cox clinical | Clinical DSN + Resting ECG | **0.035(0.019 - 0.053) | **0.006 (0.003 - 0.009) | 0.102 ( -0.113 - 0.317) |
| Cox clinical | Clinical DSN + Exercise ECG | *0.021(0.004 - 0.04) | *0.004 (0.001- 0.007) | 0.006 ( -0.205 - 0.217) |
| Cox clinical | Clinical DSN + Recovery ECG | **0.033(0.016 - 0.053) | **0.006 (0.003 - 0.009) | -0.020 ( -0.233 - 0.193) |
| Cox clinical | Clinical DSN + Complete ECG | **0.034(0.017 - 0.053) | **0.006 (0.003 - 0.009) | -0.008 ( -0.219 - 0.203) |
| **Male** | | | | |
| Cox clinical | Clinical DSN + Resting ECG | **0.05(0.026 - 0.079) | **0.011 (0.006 - 0.017) | 0.039 ( -0.214 - 0.293) |
| Cox clinical | Clinical DSN + Exercise ECG | *0.029(0.005 - 0.058) | * 0.007 (0.001 - 0.013) | 0.006 ( -0.247 - 0.260) |
| Cox clinical | Clinical DSN + Recovery ECG | **0.048(0.021 - 0.079) | ** 0.011 (0.005 - 0.017) | -0.049 ( -0.303 - 0.205) |
| Cox clinical | Clinical DSN + Complete ECG | **0.048(0.023 - 0.077) | ** 0.011 (0.005 - 0.017) | -0.017 ( -0.253 - 0.219) |
| **Female** | | | | |
| Cox clinical | Clinical DSN + Resting ECG | 0.005(0 - 0.018) | 0.001 ( -0.001 - 0.002) | 0.252 ( -0.144 - 0.649) |
| Cox clinical | Clinical DSN + Exercise ECG | 0.005(0 - 0.018) | 0.001 ( -0.001 - 0.002) | 0.033 ( -0.347 - 0.413) |
| Cox clinical | Clinical DSN + Recovery ECG | 0.005(0 - 0.018) | 0.001 ( -0.001 - 0.002) | 0.066 ( -0.324 - 0.456) |
| Cox clinical | Clinical DSN + Complete ECG | 0.005(0 - 0.018) | 0.001 ( -0.001 - 0.002) | 0.155 ( -0.246 - 0.557) |

Abbreviations: NRI = Net re classification index, DSN = Deep survival network, CI = Confidence interval
Data are reported on 41,076 participants with 3,053 (7·4%) Major Adverse Cardiovascular & Cerebrovascular events (MACCE) over 10 years. 95% confidence intervals and p– values were obtained via 1000 bootstrap iterations with a significance threshold of 0·05. **p value <0·05; ** p value < 0·001.*

Each row corresponds to a separate risk score, fitted independently with the selected risk factor and independent estimates of the baseline survival using Breslow's method, with mean-centred continuous variables, with all binary risk factors set to 0.

Clinical risk scores were derived using Cox proportional hazards regression. Extended Cox models additionally included the $ECG_{AI}$ scores as a covariate.

Cox clinical: Age FP1 + Age FP2 + BMI FP1 + BMI FP2 + SBP + SBP5 + Diabetes type 1 + Diabetes type 2 + HDL ratio + Townsend deprivation index + Smoker former + Smoker current + Smoker never + Ethnic Background + Corticosteroid + Migraine + Hypertension + Severe mental illness + Age FP1 × Townsend deprivation index + Age FP2 × Townsend deprivation index + Age FP1 × BMI FP 1 + Age FP1 × BMI FP 2 + Age FP1 × BP + Age FP1 × Diabetes type 1 + Age FP1 × Diabetes type 2 + Age FP1 × hypertension + Age FP2 × BMI FP1 + Age FP2 × BMI FP2 + Age FP2 × SBP + Age FP2 × Diabetes type1 + Age FP2 × Diabetes type2 + Age FP2 × Hypertension.

Fractional polynomials: *Age FP1 (males) =* $(age/10)^{-1}$, *Age FP2 (males) =* $(age/10)^{3}$, *BMI FP 1 (males) =* $(BMI/10)^{-2}$, *BMI FP 2 (males) =* $(BMI/10)^{-2}\ X\ log(BMI/10)$, *Age FP1 (Females) =* $age/10^{-2}$, *Age FP2 (Females) = age/10, BMI FP 1 (Females) =* $(BMI/10)^{-2}$, *BMI FP 2 (Females) =* $(BMI/10)^{-2}\ X\ log\ (BMI/10)$.

Clinical DSN: Age + BMI + systolic blood-pressure + SBP5 +Diabetes type 1+ Diabetes type 2 + HDL ratio + Townsend deprivation index+ Smoker former + Smoker current + Smoker never+ Ethnic Background+ Corticosteroid+ Migraine + Hypertension + Severe mental illness,

Clinical DSN + ECG risk scores were derived from separate deep survival networks trained on raw ECG recordings from: 15s rest, final 15s of exercise (peak intensity), final 15s recovery, and the full 7 min 15s submaximal ECG, respectively.

**Supplementary Table 15: Reclassification performance for 10-year MACCE risk prediction at a 10% risk threshold among participants aged 45–55 years comparing Cox clinical and Clinical Deep Survival Network (DSN) + ECG models derived from rest, exercise, recovery, and the complete submaximal exercise ECG recording**

| Model | Comparator | Net-benefit (95% CI) | Categorical NRI (95% CI) | Continuous NRI (95% CI) |
|---|---|---|---|---|
| **Overall** | | | | |
| Cox clinical | Clinical DSN + Resting ECG | -0.006(-0.031 - 0.019) | -0.008 ( -0.034 - 0.018) | 0.051 ( -0.0411 - 0.143) |
| Cox clinical | Clinical DSN + Exercise ECG | -0.026(-0.058 - 0.008) | -0.012 ( -0.042 - 0.018) | 0.061 ( -0.033 - 0.155) |
| Cox clinical | Clinical DSN + Recovery ECG | -0.001(-0.029 - 0.028) | -0.003 ( -0.031- 0.025) | 0.037 ( -0.056 - 0.131) |
| Cox clinical | Clinical DSN + Complete ECG | -0.008(-0.036 - 0.021) | -0.009 ( -0.035- 0.018) | 0.049 ( -0.040 - 0.139) |
| **Male** | | | | |
| Cox clinical | Clinical DSN + Resting ECG | -0.005(-0.041 - 0.029) | -0.010 ( -0.046 - 0.025) | *0.172 (0.065 - 0.279) |
| Cox clinical | Clinical DSN + Exercise ECG | -0.031(-0.076 - 0.016) | -0.019 ( -0.0618 - 0.023) | *0.131 (0.020 - 0.242) |
| Cox clinical | Clinical DSN + Recovery ECG | 0.007(-0.033 - 0.048) | 0.0018 ( -0.037 - 0.041) | *0.179 (0.069 - 0.288) |
| Cox clinical | Clinical DSN + Complete ECG | -0.005(-0.042 - 0.033 | -0.009 ( -0.046 - 0.028) | ** 0.201 (0.096 - 0.306) |
| **Female** | | | | |
| Cox clinical | Clinical DSN + Resting ECG | 0.009(-0.034 - 0.02) | 0.004 ( -0.022 - 0.029) | 0.105 ( -0.065 - 0.275) |
| Cox clinical | Clinical DSN + Exercise ECG | -0.015(-0.036 - 0.006) | -0.003 ( -0.023 - 0.018) | 0.137 ( -0.03 - 0.307) |
| Cox clinical | Clinical DSN + Recovery ECG | -0.021(-0.048 - 0.005) | -0.010 ( -0.035 - 0.015) | 0.074 ( -0.096 - 0.243) |
| Cox clinical | Clinical DSN + Complete ECG | -0.014(-0.036 - 0.008) | -0.003 ( -0.023 - 0.018) | 0.007 ( -0.161 - 0.174) |

Abbreviations: NRI = Net re classification index, DSN = Deep survival network, CI = Confidence interval
Data are reported on 41,076 participants with 3,053 (7·4%) Major Adverse Cardiovascular & Cerebrovascular events (MACCE) over 10 years. 95% confidence intervals and p– values were obtained via 1000 bootstrap iterations with a significance threshold of 0·05. **p value <0·05; ** p value < 0·001.*

Each row corresponds to a separate risk score, fitted independently with the selected risk factor and independent estimates of the baseline survival using Breslow's method, with mean-centred continuous variables, with all binary risk factors set to 0.

Clinical risk scores were derived using Cox proportional hazards regression. Extended Cox models additionally included the $ECG_{AI}$ scores as a covariate.

Cox clinical: Age FP1 + Age FP2 + BMI FP1 + BMI FP2 + SBP + SBP5 + Diabetes type 1 + Diabetes type 2 + HDL ratio + Townsend deprivation index + Smoker former + Smoker current + Smoker never + Ethnic Background + Corticosteroid + Migraine + Hypertension + Severe mental illness + Age FP1 × Townsend deprivation index + Age FP2 × Townsend deprivation index + Age FP1 × BMI FP 1 + Age FP1 × BMI FP 2 + Age FP1 × BP + Age FP1 × Diabetes type 1 + Age FP1 × Diabetes type 2 + Age FP1 × hypertension + Age FP2 × BMI FP1 + Age FP2 × BMI FP2 + Age FP2 × SBP + Age FP2 × Diabetes type1 + Age FP2 × Diabetes type2 + Age FP2 × Hypertension.

Fractional polynomials: *Age FP1 (males) =* $(age/10)^{-1}$, *Age FP2 (males) =* $(age/10)^3$, *BMI FP 1 (males) =* $(BMI/10)^{-2}$, *BMI FP 2 (males) =* $(BMI/10)^{-2}\ X\ log(BMI/10)$, *Age FP1 (Females) =* $age/10^{-2}$, *Age FP2 (Females) = age/10, BMI FP 1 (Females) =* $(BMI/10)^{-2}$, *BMI FP 2 (Females) =* $(BMI/10)^{-2}\ X\ log\ (BMI/10)$.

Clinical DSN: Age + BMI + systolic blood-pressure + SBP5 +Diabetes type 1+ Diabetes type 2 + HDL ratio + Townsend deprivation index+ Smoker former + Smoker current + Smoker never+ Ethnic Background+ Corticosteroid+ Migraine + Hypertension + Severe mental illness,

Clinical DSN + ECG risk scores were derived from separate deep survival networks trained on raw ECG recordings from: 15s rest, final 15s of exercise (peak intensity), final 15s recovery, and the full 7 min 15s submaximal ECG, respectively.

**Supplementary Table 16 Reclassification performance for 10-year MACCE risk prediction at a 10% risk threshold among participants aged 55–65 years comparing Cox clinical and Clinical Deep Survival Network (DSN) + ECG models derived from rest, exercise, recovery, and the complete submaximal exercise ECG recording.**

| Model | Comparator | Net-benefit (95% CI) | Categorical NRI (95% CI) | Continuous NRI (95% CI) |
|---|---|---|---|---|
| **Overall** | | | | |
| Cox clinical | Clinical DSN + Resting ECG | *-0.029(-0.049 - -0.009) | -0.024 ( -0.043 - -0.005) | 0.033 ( -0.0205 - 0.087) |
| Cox clinical | Clinical DSN + Exercise ECG | *-0.034(-0.057 - -0.012) | *-0.030 ( -0.052 - -0.009) | *0.085 (0.031 - 0.138) |
| Cox clinical | Clinical DSN + Recovery ECG | *-0.024(-0.043 - -0.004) | -0.019 ( -0.038 – 0.001) | * 0.061 (0.007 - 0.115) |
| Cox clinical | Clinical DSN + Complete ECG | *-0.023(-0.043 - -0.004) | -0.016 ( -0.035 - 0.002) | ** 0.176 (0.122 - 0.230) |
| **Male** | | | | |
| Cox clinical | Clinical DSN + Resting ECG | **-0.038(-0.064 - -0.013) | * -0.036( -0.061 - -0.012) | *0.096 (0.027 - 0.165) |
| Cox clinical | Clinical DSN + Exercise ECG | **-0.047(-0.075 - -0.016 | *-0.046( -0.075 - -0.018) | **0.114 (0.046 - 0.182) |
| Cox clinical | Clinical DSN + Recovery ECG | *-0.031(-0.055 - -0.008) | * -0.030 ( -0.054- -0.006) | * 0.115(0.046 - 0.184) |
| Cox clinical | Clinical DSN + Complete ECG | *-0.027(-0.051 - -0.003) | *-0.027 ( -0.051 - -0.002) | **0.170(0.102 - 0.237) |
| **Female** | | | | |
| Cox clinical | Clinical DSN + Resting ECG | -0.01(-0.045 - 0.023) | 0.047 (0.018 - 0.076) | ** 0.134 (0.054 - 0.214) |
| Cox clinical | Clinical DSN + Exercise ECG | -0.01(-0.045 - 0.026) | * 0.036 (0.005 - 0.067) | *0.114 (0.026 - 0.202) |
| Cox clinical | Clinical DSN + Recovery ECG | -0.011(-0.042 - 0.02) | * 0.042(0.014 - 0.071) | ** 0.161 (0.079 - 0.244) |
| Cox clinical | Clinical DSN + Complete ECG | -0.016(-0.046 - 0.015) | * 0.036 (0.009 - 0.064) | **0.203 (0.112 - 0.293) |

Abbreviations: NRI = Net re classification index, DSN = Deep survival network, CI = Confidence interval
Data are reported on 41,076 participants with 3,053 (7·4%) Major Adverse Cardiovascular & Cerebrovascular events (MACCE) over 10 years. 95% confidence intervals and p– values were obtained via 1000 bootstrap iterations with a significance threshold of 0·05. **p value <0·05; ** p value < 0·001.*

Each row corresponds to a separate risk score, fitted independently with the selected risk factor and independent estimates of the baseline survival using Breslow's method, with mean-centred continuous variables, with all binary risk factors set to 0.

Clinical risk scores were derived using Cox proportional hazards regression. Extended Cox models additionally included the $ECG_{AI}$ scores as a covariate.

Cox clinical: Age FP1 + Age FP2 + BMI FP1 + BMI FP2 + SBP + SBP5 + Diabetes type 1 + Diabetes type 2 + HDL ratio + Townsend deprivation index + Smoker former + Smoker current + Smoker never + Ethnic Background + Corticosteroid + Migraine + Hypertension + Severe mental illness + Age FP1 × Townsend deprivation index + Age FP2 × Townsend deprivation index + Age FP1 × BMI FP 1 + Age FP1 × BMI FP 2 + Age FP1 × BP + Age FP1 × Diabetes type 1 + Age FP1 × Diabetes type 2 + Age FP1 × hypertension + Age FP2 × BMI FP1 + Age FP2 × BMI FP2 + Age FP2 × SBP + Age FP2 × Diabetes type1 + Age FP2 × Diabetes type2 + Age FP2 × Hypertension.

Fractional polynomials: *Age FP1 (males)* = $(age/10)^{-1}$, *Age FP2 (males) = (age/10)*$^{3}$, *BMI FP 1 (males)* = $(BMI/10)^{-2}$, *BMI FP 2 (males)* = $(BMI/10)^{-2}\ X\ log(BMI/10)$, *Age FP1 (Females)* = $age/10^{-2}$, *Age FP2 (Females) = age/10, BMI FP 1 (Females)* = $(BMI/10)^{-2}$, *BMI FP 2 (Females)* = $(BMI/10)^{-2}\ X\ log\ (BMI/10)$.

Clinical DSN: Age + BMI + systolic blood-pressure + SBP5 +Diabetes type 1+ Diabetes type 2 + HDL ratio + Townsend deprivation index+ Smoker former + Smoker current + Smoker never+ Ethnic Background+ Corticosteroid+ Migraine + Hypertension + Severe mental illness,

Clinical DSN + ECG risk scores were derived from separate deep survival networks trained on raw ECG recordings from: 15s rest, final 15s of exercise (peak intensity), final 15s recovery, and the full 7 min 15s submaximal ECG, respectively.

#### Supplementary Table 17 Reclassification performance for 10-year MACCE risk prediction at a 10% risk threshold among participants aged >65 years comparing Cox clinical and Clinical Deep Survival Network (DSN) + ECG models derived from rest, exercise, recovery, and the complete submaximal exercise ECG recording.

| Model | Comparator | Net-benefit (95% CI) | Categorical NRI (95% CI) | Continuous NRI (95% CI) |
|---|---|---|---|---|
| **Overall** | | | | |
| Cox clinical | Clinical DSN + Resting ECG | 0.006(-0.014 - 0.026) | -0.005 ( -0.025 - 0.015) | -0.014 ( -0.072 - 0.045) |
| Cox clinical | Clinical DSN + Exercise ECG | *-0.025(-0.049 - 0) | -0.011 ( -0.032 - 0.011) | -0.028 ( -0.088 - 0.032) |
| Cox clinical | Clinical DSN + Recovery ECG | 0.006(-0.014 - 0.026) | -0.025 ( -0.051 – 0.001) | -0.012 ( -0.072 - 0.049) |
| Cox clinical | Clinical DSN + Complete ECG | 0.017(-0.002 - 0.036) | -0.011 ( -0.032 - 0.010) | -0.046 ( -0.105 - 0.014) |
| **Male** | | | | |
| Cox clinical | Clinical DSN + Resting ECG | -0.01(-0.025 - 0.002) | 0.0137 ( -0.001 - 0.028) | 0.023 ( -0.051 - 0.098) |
| Cox clinical | Clinical DSN + Exercise ECG | **-0.06(-0.083 - -0.036) | -0.004 ( -0.028 - 0.020) | -0.026 ( -0.100 - 0.049) |
| Cox clinical | Clinical DSN + Recovery ECG | *-0.017(-0.031 - -0.003) | 0.006 ( -0.009 - 0.021) | -0.019 ( -0.094 - 0.056) |
| Cox clinical | Clinical DSN + Complete ECG | -0.004(-0.015 - 0.006) | 0.010 ( -0.001 - 0.022) | -0.041 ( -0.113 - 0.032) |
| **Female** | | | | |
| Cox clinical | Clinical DSN + Resting ECG | 0.044(-0.016 - 0.104) | * 0.069 (0.015 - 0.123) | 0.000 ( -0.098 - 0.099) |
| Cox clinical | Clinical DSN + Exercise ECG | *0.058(-0.003 - 0.115) | * 0.076 (0.019 - 0.132) | 0.039 ( -0.0684 - 0.146) |
| Cox clinical | Clinical DSN + Recovery ECG | **0.46(0.396 - 0.527) | *0.085 (0.030 - 0.139) | 0.075 ( -0.025 - 0.174) |
| Cox clinical | Clinical DSN + Complete ECG | *0.068(0.01 - 0.125) | ** 0.095(0.042 - 0.148) | *0.099 (0.001- 0.198) |

Abbreviations: NRI = Net re classification index, DSN = Deep survival network, CI = Confidence interval
Data are reported on 41,076 participants with 3,053 (7·4%) Major Adverse Cardiovascular & Cerebrovascular events (MACCE) over 10 years. 95% confidence intervals and p– values were obtained via 1000 bootstrap iterations with a significance threshold of 0·05. **p value <0·05; ** p value < 0·001.*

Each row corresponds to a separate risk score, fitted independently with the selected risk factor and independent estimates of the baseline survival using Breslow's method, with mean-centred continuous variables, with all binary risk factors set to 0.

Clinical risk scores were derived using Cox proportional hazards regression. Extended Cox models additionally included the $ECG_{AI}$ scores as a covariate.

Cox clinical: Age FP1 + Age FP2 + BMI FP1 + BMI FP2 + SBP + SBP5 + Diabetes type 1 + Diabetes type 2 + HDL ratio + Townsend deprivation index + Smoker former + Smoker current + Smoker never + Ethnic Background + Corticosteroid + Migraine + Hypertension + Severe mental illness + Age FP1 × Townsend deprivation index + Age FP2 × Townsend deprivation index + Age FP1 × BMI FP 1 + Age FP1 × BMI FP 2 + Age FP1 × BP + Age FP1 × Diabetes type 1 + Age FP1 × Diabetes type 2 + Age FP1 × hypertension + Age FP2 × BMI FP1 + Age FP2 × BMI FP2 + Age FP2 × SBP + Age FP2 × Diabetes type1 + Age FP2 × Diabetes type2 + Age FP2 × Hypertension.

Fractional polynomials: *Age FP1 (males)* = $(age/10)^{-1}$, *Age FP2 (males) = (age/10)*$^3$, *BMI FP 1 (males)* = $(BMI/10)^{-2}$, *BMI FP 2 (males)* = $(BMI/10)^{-2}\ X\ log(BMI/10)$, *Age FP1 (Females)* = $age/10^{-2}$, *Age FP2 (Females) = age/10, BMI FP 1 (Females)* = $(BMI/10)^{-2}$, *BMI FP 2 (Females)* = $(BMI/10)^{-2}\ X\ log\ (BMI/10)$.

Clinical DSN: Age + BMI + systolic blood-pressure + SBP5 +Diabetes type 1+ Diabetes type 2 + HDL ratio + Townsend deprivation index+ Smoker former + Smoker current + Smoker never+ Ethnic Background+ Corticosteroid+ Migraine + Hypertension + Severe mental illness

Clinical DSN + ECG risk scores were derived from separate deep survival networks trained on raw ECG recordings from: 15s rest, final 15s of exercise (peak intensity), final 15s recovery, and the full 7 min 15s submaximal ECG, respectively.

**Supplementary Table 18 Overall performance for 10-year MACCE risk prediction comparing Age + sex and Cox clinical risk scores after the addition of $ECG_{AI}$ risk scores, assessed using Harrell's C-index, integrated calibration index (ICI), and net benefit at a 10% risk threshold**

| Model | C-Index | ICI | Net benefit |
|---|---|---|---|
| Age+ Sex | 0.693(0.686 - 0.7) | 0.0029(0.0016 - 0.0046) | 0.178(0.16 - 0.198) |
| Age + Sex + ECG | | | |
| Resting | 0.697(0.689 - 0.706) | 0.0032(0.0016 - 0.0051) | 0.185(0.165 - 0.206)) |
| Peak exercise | 0.697(0.69 - 0.704) | 0.0031(0.0016 - 0.0046) | 0.185(0.167 - 0.205) |
| Late-stage recovery | 0.698(0.691 - 0.705) | 0.0033(0.0019 - 0.0048) | 0.185(0.165 - 0.205) |
| Complete ECG | 0.699(0.692 - 0.706) | 0.0031(0.0017 - 0.0049) | 0.189(0.17 - 0.208) |
| Cox clinical | 0.672(0.665 - 0.678) | 0.0095 (0.0077 - 0.0116) | 0.218(0.196 - 0.238) |
| Cox clinical + ECG | | | |
| Resting | 0.672(0.665 - 0.68) | 0.0094(0.0076 - 0.0115) | 0.21(0.187 - 0.229) |
| Peak exercise | 0.674(0.667 - 0.682)) | 0.0095(0.0076 - 0.0118) | 0.213(0.191 - 0.236) |
| Late-stage recovery | 0.671(0.665 - 0.679) | 0.0104(0.0085 - 0.0123) | 0.217(0.193 - 0.236) |
| Complete ECG | 0.673(0.667 - 0.681) | 0.0096(0.0077 - 0.0118) | 0.219(0.195 - 0.239) |
| Cox clinical | 0.714(0.707 - 0.722) | 0.0035(0.002 - 0.0055) | 0.218(0.197 - 0.242) |
| Cox clinical+ ECG | | | |
| Resting | 0.711(0.704 - 0.718) | 0.0032(0.002 - 0.0049) | 0.212(0.191 - 0.235) |
| Peak exercise | 0.712(0.704 - 0.718) | 0.0035(0.0019 - 0.0053) | 0.215(0.193 - 0.235) |
| Late-stage recovery | 0.713(0.705 - 0.72) | 0.0031(0.0016 - 0.005) | 0.226(0.203 - 0.247) |
| Complete ECG | 0.716(0.705 - 0.72) | 0.0029(0.0015 - 0.0047) | 0.222(0.2 - 0.243) |

Abbreviations: ICI = Integrated calibration index; CI = confidence intervals.

Data are reported on 41,076 participants with 3,053 (7·4%) Major Adverse Cardiovascular & Cerebrovascular events (MACCE) over 10 years. 95% confidence intervals were obtained via 1000 bootstrap iterations

Each row corresponds to a separate risk score, fitted independently with the selected risk factors., and independent estimates of the baseline survival using Breslow's method, with mean-centred continuous variables, with all binary risk factors set to 0

Age + Sex and clinical risk factors were integrated with ECG recordings in separate deep survival networks trained on raw ECG recordings during 15-seconds of rest, the final 15-seconds of exercise (peak exercise intensity), final 15-seconds of recovery and full 7-minute 15-second submaximal exercise ECG recording, respectively.

Cox clinical: Age FP1 + Age FP2 + BMI FP1 + BMI FP2 + SBP + SBP5 + Diabetes type 1 + Diabetes type 2 + HDL ratio + Townsend deprivation index + Smoker former + Smoker current + Smoker never + Ethnic Background + Corticosteroid + Migraine + Hypertension + Severe mental illness + Age FP1 × Townsend deprivation index + Age FP2 × Townsend deprivation index + Age FP1 × BMI FP 1 + Age FP1 × BMI FP 2 + Age FP1 × BP + Age FP1 × Diabetes type 1 + Age FP1 × Diabetes type 2 + Age FP1 × hypertension + Age FP2 × BMI FP1 + Age FP2 × BMI FP2 + Age FP2 × SBP + Age FP2 × Diabetes type1 + Age FP2 × Diabetes type2 + Age FP2 × Hypertension.

Fractional polynomials: *Age FP1 (males) =* $(age/10)^{-1}$, *Age FP2 (males) = (age/10)*$^3$, *BMI FP 1 (males) =* $(BMI/10)^{-2}$, *BMI FP 2 (males) =* $(BMI/10)^{-2}\ X\ log(BMI/10)$, *Age FP1 (Females) =* $age/10^{-2}$, *Age FP2 (Females) = age/10, BMI FP 1 (Females) =* $(BMI/10)^{-2}$, *BMI FP 2 (Females) =* $(BMI/10)^{-2}\ X\ log\ (BMI/10)$.

**Supplementary Table 19: Change in discrimination (C-index) and calibration (ICI) for 10-year MACCE risk prediction after adding $ECG_{AI}$ risk scores derived from 15-second rest, exercise, recovery, and complete 7-minute 15-second ECG recordings from the submaximal exercise test to Age + sex and Cox clinical risk scores.**

| Model | Comparator | ΔC-index | ΔICI |
|---|---|---|---|
| **Study population** | | | |
| Age + Sex | Age + Sex+ Resting ECG | ** 0.004(0.002 - 0.005) | 0(-0.001 - 0.001) |
| Age + Sex | Age + Sex+ Exercise ECG | * 0.003(0.001 - 0.005) | 0(-0.001 - 0.001) |
| Age + Sex | Age + Sex+ Recovery ECG | ** 0.004(0.003 - 0.006) | 0.000(-0.001 - 0.002) |
| Age + Sex | Age + Sex+ Complete ECG | ** 0.005(0.004 - 0.007) | 0(-0.001 - 0.001) |
| Cox clinical | Cox clinical+ Resting ECG | 0(-0.001 - 0.001) | |
| Cox clinical | Cox clinical+ Exercise ECG | ** 0.002(0.001 - 0.004) | 0(-0.001 - 0.001) |
| Cox clinical | Cox clinical+ Recovery ECG | 0(-0.002 - 0.001) | *0.001(0 - 0.002) |
| Cox clinical | Cox clinical+ Complete ECG | ** 0.002(0 - 0.003) | 0(-0.001 - 0.001) |
| **Male** | | | |
| Age + Sex | Age + Sex+ Resting ECG | ** 0.005(0.002 - 0.008) | -0.000(-0.002 - 0.002) |
| Age + Sex | Age + Sex+ Exercise ECG | ** 0.003(0.001 - 0.006) | 0(-0.002 - 0.002) |
| Age + Sex | Age + Sex+ Recovery ECG | ** 0.007(0.005 - 0.01) | 0(-0.002 - 0.002) |
| Age + Sex | Age + Sex+ Complete ECG | ** 0.009(0.006 - 0.012) | -0.000(-0.002 - 0.002) |
| Cox clinical | Cox clinical+ Resting ECG | 0(-0.002 - 0.002) | *-0.002(-0.003 - 0) |
| Cox clinical | Cox clinical+ Exercise ECG | ** 0.002(0 - 0.004) | 0(-0.002 - 0.001) |
| Cox clinical | Cox clinical+ Recovery ECG | ** 0.003(0.002 - 0.005) | 0(-0.001 - 0.002) |
| Cox clinical | Cox clinical+ Complete ECG | ** 0.003(0.001 - 0.005) | 0(-0.002 - 0.001) |
| **Female** | | | |
| Age + Sex | Age + Sex+ Resting ECG | 0.004(-0.001 - 0.008) | 0.000(-0.001 - 0.002) |
| Age + Sex | Age + Sex+ Exercise ECG | ** 0.006(0.002 - 0.01) | 0.000(-0.001 - 0.001) |
| Age + Sex | Age + Sex+ Recovery ECG | ** 0.005(0.001 - 0.009) | 0(-0.001 - 0.001) |
| Age + Sex | Age + Sex+ Complete ECG | ** 0.005(0.001 - 0.008) | 0.000(-0.001 - 0.001) |
| Cox clinical | Cox clinical+ Resting ECG | 0.001(-0.001 - 0.003) | 4e-04(0 - 0.001) |
| Cox clinical | Cox clinical+ Exercise ECG | ** 0.003(0 - 0.005) | 3e-04(-0.001 - 0.001) |
| Cox clinical | Cox clinical+ Recovery ECG | -0.007(-0.01 - -0.004) | -0.001(-0.001 - 0.001) |
| Cox clinical | Cox clinical+ Complete ECG | 0(-0.002 - 0.002) | 0.000(-0.001 - 0.001) |

Data are reported on 41,076 participants, 3,053 (7·4%) MACCE were reported within 10-years. 95% confidence intervals and p– values were obtained via 1000 bootstrap iterations with a significance threshold of 0·05. **p value < 0·05; ** p value < 0·001*

Each row corresponds to a separate risk score, fitted independently with the selected risk factors independent estimates of the baseline survival using Breslow's method, with mean-centred continuous variables, with all binary risk factors set to 0

Age+Sex and Cox clinical risk scores were derived using Cox proportional hazards regression. Extended Cox models additionally included the $ECG_{AI}$ scores as a covariate (Age+Sex+ $ECG_{AI}$; Cox clinical+ $ECG_{AI}$).

$ECG_{AI}$ scores were derived from separate deep survival networks trained on raw ECG recordings from: 15 s rest, final 15 s of exercise (peak intensity), final 15 s recovery, and the full 7 min 15 s submaximal ECG, respectively.

Cox clinical: Age FP1 + Age FP2 + BMI FP1 + BMI FP2 + SBP + SBP5 + Diabetes type 1 + Diabetes type 2 + HDL ratio + Townsend deprivation index + Smoker former + Smoker current + Smoker never + Ethnic Background + Corticosteroid + Migraine + Hypertension + Severe mental illness + Age FP1 × Townsend deprivation index + Age FP2 × Townsend deprivation index + Age FP1 × BMI FP 1 + Age FP1 × BMI FP 2 + Age FP1 × BP + Age FP1 × Diabetes type 1 + Age FP1 × Diabetes type 2 + Age FP1 × hypertension + Age FP2 × BMI FP1 + Age FP2 × BMI FP2 + Age FP2 × SBP + Age FP2 × Diabetes type1 + Age FP2 × Diabetes type2 + Age FP2 × Hypertension.

Fractional polynomials: *Age FP1 (males) =* $(age/10)^{-1}$, *Age FP2 (males) = (age/10)*$^3$, *BMI FP 1 (males) =* $(BMI/10)^{-2}$, *BMI FP 2 (males) =* $(BMI/10)^{-2}\ X\ log(BMI/10)$, *Age FP1 (Females) =* $age/10^{-2}$, *Age FP2 (Females) = age/10, BMI FP 1 (Females) =* $(BMI/10)^{-2}$, *BMI FP 2 (Females) =* $(BMI/10)^{-2}\ X\ log\ (BMI/10)$.

**Supplementary Table 20: Overall performance for 10-year MACCE risk prediction of Clinical Deep Survival Network (DSN) models using Age + sex or QRISK3 risk factors with and without ECG data from rest, peak exercise, late-stage recovery, and the complete 7-minute 15-second submaximal exercise ECG recording, assessed using Harrell's C-index, integrated calibration index (ICI), and net benefit at a 10% risk threshold.**

| Model | C-Index (95% CI) | ICI (95% CI) | Net benefit (95% CI) |
|---|---|---|---|
| Age+ Sex | 0.693(0.686 - 0.7) | 0.0030(0.0015 - 0.0049) | 0.178(0.16 - 0.198) |
| Age + Sex + ECG (DSN) | | | |
| Resting | 0.700(0.693 - 0.707) | 0.0026(0.0013 - 0.0044) | 0.191(0.175 - 0.208) |
| Peak exercise | 0.692(0.685 - 0.699) | 0.0032(0.0017 - 0.0048) | 0.186(0.167 - 0.206) |
| Late-stage recovery | 0.702(0.695 - 0.708) | 0.0031(0.0019 - 0.0047) | 0.204(0.182 - 0.221) |
| Complete ECG | 0.692(0.685 - 0.699) | 0.0044(0.0026 - 0.0063) | 0.185(0.165 - 0.205) |
| Clinical DSN | 0.7(0.692 - 0.708) | 0.004(0.003 - 0.006) | 0.209(0.193 - 0.232) |
| Clinical DSN + ECG | | | |
| Resting | 0.709(0.702 - 0.717) | 0.0040 (0.0020 - 0.0061) | 0.207(0.188 - 0.229) |
| Peak exercise | 0.697(0.69 - 0.704) | 0.0035 (0.0016 - 0.0055) | 0.189(0.168 - 0.208 |
| Late-stage recovery | 0.708(0.701 - 0.715) | 0.0026 (0.0013 - 0.0041) | 0.211(0.189 - 0.23 |
| Complete ECG | 0.704(0.697 - 0.712) | 0.0026 (0.0011 - 0.0042) | 0.214 (0.194 0.238) |

Abbreviations: ICI = Integrated calibration index; DSN = Deep survival network; CI = confidence intervals.

Data are reported on 41,076 participants with 3,053 (7·4%) Major Adverse Cardiovascular & Cerebrovascular events (MACCE) over 10 years. 95% confidence intervals were obtained via 1000 bootstrap iterations

Each row corresponds to a separate risk score, fitted independently with the selected risk factors., and independent estimates of the baseline survival using Breslow's method, with mean-centred continuous variables, with all binary risk factors set to 0

Age + Sex and clinical risk factors were integrated with ECG recordings in separate deep survival networks trained on raw ECG recordings during 15-seconds of rest, the final 15-seconds of exercise (peak exercise intensity), final 15-seconds of recovery and full 7-minute 15-second submaximal exercise ECG recording, respectively.

Clinical DSN: Age + BMI + systolic blood-pressure + SBP5 +Diabetes type 1+ Diabetes type 2 + HDL ratio + Townsend deprivation index+ Smoker former + Smoker current + Smoker never+ Ethnic Background+ Corticosteroid+ Migraine + Hypertension + Severe mental illness

Clinical DSN + ECG risk scores were derived from separate deep survival networks trained on raw ECG recordings from: 15s rest, final 15s of exercise (peak intensity), final 15s recovery, and the full 7 min 15s submaximal ECG, respectively.

**Supplementary Table 21: Change in discrimination (C-index) and calibration (ICI) for 10-year MACCE risk prediction after adding ECG data derived from 15-second rest, exercise, recovery, and complete 7-minute 15-second ECG recordings from the submaximal exercise test to Deep Survival Network (DSN) models based on Age + sex and QRISK3 risk factors.**

| Model | Comparator | ΔC-index | ΔICI |
|---|---|---|---|
| **Study population** | | | |
| Age + Sex | Age + Sex+ Resting ECG | **0.006(0.003 - 0.01) | 0.000(-0.002 - 0.002) |
| Age + Sex | Age + Sex+ Exercise ECG | *0.004(0.002 - 0.007) | 0.000(-0.001 - 0.002) |
| Age + Sex | Age + Sex+ Recovery ECG | **0.008(0.005 - 0.012) | 0.000(-0.002 - 0.002) |
| Age + Sex | Age + Sex+ Complete ECG | -0.002(-0.005 - 0.002) | 0.001(-0.001 - 0.004) |
| Cox clinical | Clinical DSN + Resting ECG | **0.037(0.032 - 0.042) | **-0.006(-0.007 - -0.004) |
| Cox clinical | Clinical DSN + Exercise ECG | **0.025(0.02 - 0.031) | **-0.007(-0.009 - -0.004) |
| Cox clinical | Clinical DSN + Recovery ECG | **0.036(0.03 - 0.04) | **-0.006(-0.008 - -0.004) |
| Cox clinical | Clinical DSN + Complete ECG | **0.033(0.027 - 0.038) | **-0.007(-0.009 - -0.005) |
| | | **0.006(0.003 - 0.01) | |
| Clinical DSN | Clinical DSN + Resting ECG | *0.004(0.002 - 0.007) | -0.001(-0.002 - 0.001) |
| Clinical DSN | Clinical DSN + Exercise ECG | **0.008(0.005 - 0.012) | -0.002(-0.005 - 0.001) |
| Clinical DSN | Clinical DSN + Recovery ECG | **-0.002(-0.005 - 0.002) | -0.001(-0.002 - 0) |
| Clinical DSN | Clinical DSN + Complete ECG | 0.001(-0.002 - 0.003) | *-0.002(-0.004 - 0) |
| **Male** | | | |
| Age + Sex | Age + Sex+ Resting ECG | **0.011(0.006 - 0.016) | 0.001(-0.005 - 0.003) |
| Age + Sex | Age + Sex+ Exercise ECG | *0.007(0.003 - 0.011) | 0.000(-0.004 - 0.004) |
| Age + Sex | Age + Sex+ Recovery ECG | **0.012(0.007 - 0.017) | -0.001(-0.004 - 0.003) |
| Age + Sex | Age + Sex+ Complete ECG | -0.001(-0.006 - 0.003) | 0(-0.004 - 0.004) |
| Cox clinical | Clinical DSN + Resting ECG | 0.005(-0.001 - 0.01) | **-0.017(-0.02 - -0.014) |
| Cox clinical | Clinical DSN + Exercise ECG | **-0.009(-0.016 - -0.003 | **-0.0192(-0.024 - -0.014) |
| Cox clinical | Clinical DSN + Recovery ECG | 0.003(-0.002 - 0.008) | **-0.0173(-0.02 - -0.015) |
| Cox clinical | Clinical DSN + Complete ECG | -0.002(-0.007 - 0.003) | **-0.0198(-0.023 - -0.016) |
| Clinical DSN | Clinical DSN + Resting ECG | 0.003(-0.002 - 0.007) | -0.001(-0.003 - 0.001) |
| Clinical DSN | Clinical DSN + Exercise ECG | -0.011(-0.017 - -0.006) | -0.003(-0.007 - 0) |
| Clinical DSN | Clinical DSN + Recovery ECG | 0.001(-0.004 - 0.006) | 0.001(-0.003 - 0.001) |
| Clinical DSN | Clinical DSN + Complete ECG | -0.004(-0.007 - 0.001) | **-0.004(-0.006 - -0.002) |
| **Female** | | | |
| Age + Sex | Age + Sex+ Resting ECG | 0.004(-0.002 - 0.011) | 0.0001(-0.001 - 0.002) |
| Age + Sex | Age + Sex+ Exercise ECG | 0.005(0 - 0.01) | *0.0019(0 - 0.003) |
| Age + Sex | Age + Sex+ Recovery ECG | *0.01(0.003 - 0.017) | -0.000(-0.001 - 0.002) |
| Age + Sex | Age + Sex+ Complete ECG | -0.004(-0.012 - 0.003) | 0.0021(0 - 0.004) |
| Cox clinical | Clinical DSN + Resting ECG | **0.015(0.008 - 0.021) | -0.0026(-0.006 - 0.001) |
| Cox clinical | Clinical DSN + Exercise ECG | 0.005(-0.005 - 0.013) | *-0.004(-0.009 - -0.001) |
| Cox clinical | Clinical DSN + Recovery ECG | **0.014(0.009 - 0.021) | -0.0034(-0.007 - 0) |

| | | | |
|---|---|---|---|
| Cox clinical | Clinical DSN + Complete ECG | **0.011(0.005 - 0.018) | -0.0032(-0.008 - 0.001) |
| Clinical DSN | Clinical DSN + Resting ECG | **0.011(0.003 - 0.019) | 0.001(-0.001 - 0.002) |
| Clinical DSN | Clinical DSN + Exercise ECG | 0.001(-0.007 - 0.008) | -0.001(-0.003 - 0.001) |
| Clinical DSN | Clinical DSN + Recovery ECG | **0.011(0.003 - 0.018) | 0(-0.002 - 0.001) |
| Clinical DSN | Clinical DSN + Complete ECG | **0.007(0.002 - 0.011) | 0(-0.001 - 0.001 |

Abbreviations: ICI = Integrated calibration index; DSN = Deep survival network; CI = confidence intervals.

Data are reported on 41,076 participants with 3,053 (7·4%) Major Adverse Cardiovascular & Cerebrovascular events (MACCE) over 10 years. 95% confidence intervals and p– values were obtained via 1000 bootstrap iterations with a significance threshold of 0·05. ** p value < 0·05; ** p value < 0·001.*

Each row corresponds to a separate risk score, fitted independently with the selected risk factors independent estimates of the baseline survival using Breslow's method, with mean-centred continuous variables, with all binary risk factors set to 0.

Age+Sex and Cox clinical risk scores were derived using Cox proportional hazards regression. Extended Cox models additionally included the $ECG_{AI}$ score as a covariate (Age+Sex+ $ECG_{AI}$; Cox clinical+ $ECG_{AI}$).

$ECG_{AI}$ scores were derived from separate deep survival networks trained on raw ECG recordings from: 15 s rest, final 15 s of exercise (peak intensity), final 15 s recovery, and the full 7 min 15 s submaximal ECG, respectively.

Clinical DSN + ECG integrated QRISK3 risk factors and raw ECG data directly within a single deep survival model.

Cox clinical: Age FP1 + Age FP2 + BMI FP1 + BMI FP2 + SBP + SBP5 + Diabetes type 1 + Diabetes type 2 + HDL ratio + Townsend deprivation index + Smoker former + Smoker current + Smoker never + Ethnic Background + Corticosteroid + Migraine + Hypertension + Severe mental illness + Age FP1 × Townsend deprivation index + Age FP2 × Townsend deprivation index + Age FP1 × BMI FP 1 + Age FP1 × BMI FP 2 + Age FP1 × BP + Age FP1 × Diabetes type 1 + Age FP1 × Diabetes type 2 + Age FP1 × hypertension + Age FP2 × BMI FP1 + Age FP2 × BMI FP2 + Age FP2 × SBP + Age FP2 × Diabetes type1 + Age FP2 × Diabetes type2 + Age FP2 × Hypertension.

Fractional polynomials: *Age FP1 (males) =* $(age/10)^{-1}$, *Age FP2 (males) =* $(age/10)^{3}$, *BMI FP 1 (males) =* $(BMI/10)^{-2}$, *BMI FP 2 (males) =* $(BMI/10)^{-2}\ X\ log(BMI/10)$, *Age FP1 (Females) =* $age/10^{-2}$, *Age FP2 (Females) = age/10, BMI FP 1 (Females) =* $(BMI/10)^{-2}$, *BMI FP 2 (Females) =* $(BMI/10)^{-2}\ X\ log\ (BMI/10)$.

Clinical DSN: Age, BMI, systolic blood-pressure, SBP5, Diabetes type 1, Diabetes type 2, HDL ratio, Townsend deprivation index, Smoker former, Smoker current, Smoker never, Ethnic Background, Corticosteroid, Migraine, Hypertension, Severe mental illness.

**Supplementary Table 22 Reclassification performance for 10-year MACCE risk prediction at a 10% risk threshold after adding ECG risk data measured during rest, exercise, recovery, and the complete 7-minute 15-second submaximal exercise ECG recording to Age + sex, Cox clinical, and Clinical Deep Survival Network (DSN) models.**

| Model | Comparator | Net-benefit (95% CI) | Categorical NRI (95% CI) | Continuous NRI (95% CI) |
|---|---|---|---|---|
| Age + Sex | Age + Sex+ Resting ECG | 0.007(-0.002 - 0.016) | 0.007 ( -0.0024 - 0.016) | **0.144 (0.107 - 0.181) |
| Age + Sex | Age + Sex+ Exercise ECG | 0.006(-0.002 - 0.015) | 0.006 (-0.0028 - 0.015) | **0.119 (0.082 - 0.156) |
| Age + Sex | Age + Sex+ Recovery ECG | 0.006(-0.001 - 0.015) | 0.006 (-0.0028 - 0.015) | **0.139 (0.102 - 0.176) |
| Age + Sex | Age + Sex+ Complete ECG | **0.01(0.002 - 0.018) | 0.009 ( -0.003 - 0.018) | **0.175 (0.139 - 0.211) |
| Cox clinical | Cox clinical+ Resting ECG | *-0.009(-0.015 - -0.003) | * -0.0095 ( -0.016 - -0.003) | *0.056 (0.019 - 0.093) |
| Cox clinical | Cox clinical+ Exercise ECG | -0.005(-0.011 - 0.001) | *-0.0071 ( -0.0137 - -5e-04) | **0.072 (0.035 - 0.109) |
| Cox clinical | Cox clinical+ Recovery ECG | -0.001(-0.007 - 0.004) | -0.0022 ( -0.0078 - 0.003) | 0.031 (-0.006 - 0.068) |
| Cox clinical | Cox clinical+ Complete ECG | 0.001(-0.006 - 0.007) | 0.000( -0.007 - 0.007) | **0.097 (0.061 - 0.133) |
| Clinical DSN | Clinical DSN +Resting ECG | -0.001(-0.013 - 0.01) | 0.005 ( -0.008 - 0.017) | **0.074 (0.038 - 0.111) |
| Clinical DSN | Clinical DSN +Exercise ECG | **-0.02(-0.03 - -0.005) | *-0.016 ( -0.030 - -0.003) | **0.074 (0.037 - 0.110) |
| Clinical DSN | Clinical DSN +Recovery ECG | 0.002(-0.01 - 0.012) | 0.008 ( -0.005 - 0.020) | **0.050 (0.013 - 0.086) |
| Clinical DSN | Clinical DSN +Complete ECG | 0.005(-0.007 - 0.016) | * 0.014 (0.003 - 0.025) | **0.272 (0.235 - 0.308) |

Abbreviations: NRI = Net re classification index, DSN = Deep survival network, CI = Confidence interval

Data are reported on 41,076 participants with 3,053 (7·4%) Major Adverse Cardiovascular & Cerebrovascular events (MACCE) over 10 years. 95% confidence intervals and p– values were obtained via 1000 bootstrap iterations with a significance threshold of 0·05. **p value < 0·05; ** p value < 0·001*

Each row corresponds to a separate risk score, fitted independently with the selected risk factors independent estimates of the baseline survival using Breslow's method, with mean-centred continuous variables, with all binary risk factors set to 0

Age+Sex and Cox clinical risk scores were derived using Cox proportional hazards regression. Extended Cox models additionally included the $ECG_{AI}$ score as a covariate (Age+Sex+ $ECG_{AI}$; Cox clinical+ $ECG_{AI}$).

$ECG_{AI}$ scores were derived from separate deep survival networks trained on raw ECG recordings from: 15 s rest, final 15 s of exercise (peak intensity), final 15 s recovery, and the full 7 min 15 s submaximal ECG, respectively.

Clinical DSN + ECG integrated QRISK3 risk factors and raw ECG data directly within a single deep survival model,

Cox clinical: Age FP1 + Age FP2 + BMI FP1 + BMI FP2 + SBP + SBP5 + Diabetes type 1 + Diabetes type 2 + HDL ratio + Townsend deprivation index + Smoker former + Smoker current + Smoker never + Ethnic Background + Corticosteroid + Migraine + Hypertension + Severe mental illness + Age FP1 × Townsend deprivation index + Age FP2 × Townsend deprivation index + Age FP1 × BMI FP 1 + Age FP1 × BMI FP 2 + Age FP1 × BP + Age FP1 × Diabetes type 1 + Age FP1 × Diabetes type 2 + Age FP1 × hypertension + Age FP2 × BMI FP1 + Age FP2 × BMI FP2 + Age FP2 × SBP + Age FP2 × Diabetes type1 + Age FP2 × Diabetes type2 + Age FP2 × Hypertension.

Fractional polynomials: *Age FP1 (males)* = $(age/10)^{-1}$, *Age FP2 (males)* = $(age/10)^3$, *BMI FP 1 (males)* = $(BMI/10)^{-2}$, *BMI FP 2 (males)* = $(BMI/10)^{-2}\ X\ log(BMI/10)$, *Age FP1 (Females)* = $age/10^{-2}$, *Age FP2 (Females)* = *age/10*, *BMI FP 1 (Females)* = $(BMI/10)^{-2}$, *BMI FP 2 (Females)* = $(BMI/10)^{-2}\ X\ log\ (BMI/10)$.

Clinical DSN: Age, BMI, systolic blood-pressure, SBP5, Diabetes type 1, Diabetes type 2, HDL ratio, Townsend deprivation index, Smoker former, Smoker current, Smoker never, Ethnic Background, Corticosteroid, Migraine, Hypertension, Severe mental illness.

**Supplementary Table 23 Reclassification performance for 10-year MACCE risk prediction at a 10% risk threshold comparing Age + sex and Cox clinical models with the Clinical Deep Survival Network (DSN) after adding ECG data from rest, peak exercise, late-stage recovery, and the complete 7-minute 15-second submaximal exercise ECG recording**

| **Model** | **Comparator** | Net-benefit (95% CI) | Categorical NRI (95% CI) | Continuous NRI (95% CI) |
|---|---|---|---|---|
| Age + Sex | Age + Sex+ Resting ECG (DSN) | *0.013(0.00 - 0.026) | 0.007 ( -0.006 - 0.019) | *0.055 (0.018 - 0.092) |
| Age + Sex | Age + Sex+ Exercise ECG (DSN) | 0.008(-0.004 - 0.02) | 0.004 ( -0.008 - 0.017) | 0.035 ( -0.002 - 0.071) |
| Age + Sex | Age + Sex+ Recovery ECG (DSN) | **0.026(0.013 - 0.038) | *0.018 (0.005 - 0.030) | **0.179 (0.142 - 0.216) |
| Age + Sex | Age + Sex+ Complete ECG (DSN) | 0.006(-0.005 - 0.017) | *0.013 (0.002 - 0.024) | **0.127 (0.090 - 0.164) |
| Cox clinical | Clinical DSN + Resting ECG | -0.011(-0.023 - 0.001) | -0.004 ( -0.016 - 0.008) | **0.146 (0.110 - 0.182) |
| Cox clinical | Clinical DSN +Exercise ECG | **-0.029(-0.041 - -0.016) | **-0.025 ( -0.039 - -0.011) | ** 0.144 (0.108 - 0.180) |
| Cox clinical | Clinical DSN +Recovery ECG | -0.007(-0.018 - 0.004) | -0.0014 ( -0.014 - 0.011) | **0.136 (0.099 - 0.172) |
| Cox clinical | Clinical DSN +Complete ECG | -0.004(-0.016 - 0.008) | 0.0051 ( -0.007 - 0.017) | **0.230 (0.193 - 0.266) |

Abbreviations: NRI = Net re classification index, DSN = Deep survival network, CI = Confidence interval
Data are reported on 41,076 participants with 3,053 (7·4%) Major Adverse Cardiovascular & Cerebrovascular events (MACCE) over 10 years. 95% confidence intervals and p– values were obtained via 1000 bootstrap iterations with a significance threshold of 0·05. **p value < 0·05; ** p value < 0·001*

Each row corresponds to a separate risk score, fitted independently with the selected risk factors independent estimates of the baseline survival using Breslow's method, with mean-centred continuous variables, with all binary risk factors set to 0

Age+Sex and Cox clinical risk scores were derived using Cox proportional hazards regression. Extended Cox models additionally included the $ECG_{AI}$ score as a covariate (Age+Sex+ $ECG_{AI}$; Cox clinical+ $ECG_{AI}$).

$ECG_{AI}$ scores were derived from separate deep survival networks trained on raw ECG recordings from: 15 s rest, final 15 s of exercise (peak intensity), final 15 s recovery, and the full 7 min 15 s submaximal ECG, respectively.

Clinical DSN + ECG integrated QRISK3 risk factors and raw ECG data directly within a single deep survival model.

Cox clinical: Age FP1 + Age FP2 + BMI FP1 + BMI FP2 + SBP + SBP5 + Diabetes type 1 + Diabetes type 2 + HDL ratio + Townsend deprivation index + Smoker former + Smoker current + Smoker never + Ethnic Background + Corticosteroid + Migraine + Hypertension + Severe mental illness + Age FP1 × Townsend deprivation index + Age FP2 × Townsend deprivation index + Age FP1 × BMI FP 1 + Age FP1 × BMI FP 2 + Age FP1 × BP + Age FP1 × Diabetes type 1 + Age FP1 × Diabetes type 2 + Age FP1 × hypertension + Age FP2 × BMI FP1 + Age FP2 × BMI FP2 + Age FP2 × SBP + Age FP2 × Diabetes type1 + Age FP2 × Diabetes type2 + Age FP2 × Hypertension.

Fractional polynomials: *Age FP1 (males)* = $(age/10)^{-1}$, *Age FP2 (males) = (age/10)*$^3$, *BMI FP 1 (males)* = $(BMI/10)^{-2}$, *BMI FP 2 (males)* = $(BMI/10)^{-2}\ X\ log(BMI/10)$, *Age FP1 (Females)* = $age/10^{-2}$, *Age FP2 (Females) = age/10, BMI FP 1 (Females)* = $(BMI/10)^{-2}$, *BMI FP 2 (Females)* = $(BMI/10)^{-2}\ X\ log\ (BMI/10)$.

Clinical DSN: Age, BMI, systolic blood-pressure, SBP5, Diabetes type 1, Diabetes type 2, HDL ratio, Townsend deprivation index, Smoker former, Smoker current, Smoker never, Ethnic Background, Corticosteroid, Migraine, Hypertension, Severe mental illness.

**Supplementary Table 24 Performance of multivariable Cox proportional hazards ECG risk scores for 10-year MACCE prediction, assessed using Harrell's C-index and net benefit at a 10% risk threshold in 35 597 participants with valid ECG parameters**

| Model | | Hazard Ratio (95% CI) | C-index (95% CI) | Net-benefit (95% CI) |
|---|---|---|---|---|
| Conventional ECG parameters: Cox proportional hazard | Rest | 1.09 (1.08 – 1.12) | 0.56 (0.55 0.57) | 0.07 (0.05 0.09) |
| | Exercise | 1.06 (1.05 -1.08) | 0.58 (0.576 0.59) | 0.09 (0.07 0.11) |
| | Recovery | 1.05 (1.04 – 1.09) | 0.57 (0.56 0.58) | 0.08 (0.06 0.11) |
| | Combined | 1.05 (1.05 -1.07) | 0.62(0.62 0.63) | 0.14 (0.12- 0.15) |
| Conventional ECG parameters: Deep survival | Rest | 1.06 (1.05 -1.10) | 0.59 (0.58 0.60) | 0.03 (0.02 0.04) |
| | Exercise | 1.06 (1.05 – 1.11) | 0.61 (0.60 0.62) | 0.07 (0.06 0.08) |
| | Recovery | 1.07 (1.06 -1.13) | 0.60 (0.59 0.61) | 0.05 (0.03 0.06) |
| | Combined | 1.09 (1.08 -1.11) | 0.64 (0.63 0.65) | 0.09 (0.07 0.11) |

*Abbreviation: NRI = Net reclassification index CI = Confidence intervals*

*Abbreviation: CI = Confidence intervals*
Data are reported on 35,597 participants with valid ECG parameters, 2,591 (7·27%) MACCE were reported within 10-years.95% confidence intervals were obtained via 1000 bootstrap iterations.

*Each row corresponds to a separate Cox model in which the model was fitted independently with the selected risk factors. Each risk score was derived from separate Cox proportional hazard models, with independent estimates of the baseline survival using Breslow's method, based on mean-centred continuous variables, with all binary risk factors set to 0*

Hazard ratios are derived from separate Cox proportional hazard models unadjusted for other risk factors.

Unadjusted Hazard ratios are reported per unit increase, while net benefit is reported at a 10% risk threshold.

*ECG measurements were recoded from the final 15 seconds of rest, exercise, recovery*

*Four ECG risk scores were constructed using a linear combination of risk factors from: (1) resting ECG parameters, (2) peak-exercise ECG parameters, (3) a late-stage recovery parameters, and (4) a combined ECG score including ECG parameters from all three phases.*

**ECG parameters**:

Resting ECG parameters: QRS interval, QTc interval, Anisotropy evidence, Atrial Tachycardia evidence, PVC count, regularity Evidence

Exercise ECG parameters: PR interval, QRS interval, QTc interval, RR Interval, Atrial Tachycardia evidence, PVC count, regularity evidence

Recovery ECG parameters: Irregularity evidence, QRS interval, QTc interval, Atrial Tachycardia evidence, regularity evidence

**Supplementary Table 25 Reclassification performance of Cox clinical after adding conventional ECG parameters for 10-year MACCE risk prediction at a 10% risk threshold. ECG parameters were derived from separate 15-second recordings obtained during rest, exercise, and recovery, and from combined intervals across all three stages**

| Model | Comparator | Net-benefit (95% CI) | Categorical NRI (95% CI) | Continuous NRI (95% CI) |
|---|---|---|---|---|
| **Male** | | | | |
| Cox clinical | Cox clinical+ Resting ECG | -0.005(-0.014 - 0.003) | -0.006 (-0.014 - 0.003)) | 0.03 (-0.021 - 0.08) |
| Cox clinical | Cox clinical+ Exercise ECG | 0.003(-0.008 - 0.016) | 0.003 (-0.009 - 0.015) | **0.104(0.057 - 0.156) |
| Cox clinical | Cox clinical+ Recovery ECG | 0.001(-0.009 - 0.012) | 0.001 (-0.009 - 0.011) | **0.085 (0.034 - 0.136) |
| Cox clinical | Cox clinical+ Combined ECG | -0.005(-0.02 - 0.01) | -0.006 (-0.02 - 0.009) | **0.106 (0.055 - 0.158) |
| **Female** | | | | |
| Cox clinical | Cox clinical+ Resting ECG | *-0.012(-0.024 - -0.001) | -0.010 ( -0.022 - 0.001) | 0.016 ( -0.055 - 0.087) |
| Cox clinical | Cox clinical+ Exercise ECG | 0.008(-0.015 - 0.031) | 0.009 ( -0.012 - 0.031) | 0.035 ( -0.035 - 0.105) |
| Cox clinical | Cox clinical+ Recovery ECG | 0(-0.016 - 0.018) | 0.003 ( -0.0136 - 0.020) | * 0.081 (0.011 - 0.151) |
| Cox clinical | Cox clinical+ Combined ECG | 0(-0.026 - 0.032) | 0.009 ( -0.015 - 0.033) | *0.094 (0.02 - 0.167) |

*Abbreviations: NRI = Net re classification index, CI = Confidence interval*

Data are reported on 35,597 participants with measurable ECG parameters, 2,591 (7·27%) MACCE were reported within 10-years. 95% confidence intervals and p– values were obtained via 1000 bootstrap iterations with a significance threshold of 0·05. **p value < 0·05; ** p value < 0·001*

Each row corresponds to a separate risk score, fitted independently with the selected risk factors independent estimates of the baseline survival using Breslow's method, with mean-centred continuous variables, with all binary risk factors set to 0

Age+Sex and Cox clinical risk scores were derived using Cox proportional hazards regression. Extended Cox models additionally included the $ECG_{AI}$ score as a covariate (Age+Sex+ $ECG_{AI}$; Cox clinical+ $ECG_{AI}$).

$ECG_{AI}$ scores were derived from separate deep survival networks trained on raw ECG recordings from: 15 s rest, final 15 s of exercise (peak intensity), final 15 s recovery, and the full 7 min 15 s submaximal ECG, respectively.

Cox clinical: Age FP1 + Age FP2 + BMI FP1 + BMI FP2 + SBP + SBP5 + Diabetes type 1 + Diabetes type 2 + HDL ratio + Townsend deprivation index + Smoker former + Smoker current + Smoker never + Ethnic Background + Corticosteroid + Migraine + Hypertension + Severe mental illness + Age FP1 × Townsend deprivation index + Age FP2 × Townsend deprivation index + Age FP1 × BMI FP 1 + Age FP1 × BMI FP 2 + Age FP1 × BP + Age FP1 × Diabetes type 1 + Age FP1 × Diabetes type 2 + Age FP1 × hypertension + Age FP2 × BMI FP1 + Age FP2 × BMI FP2 + Age FP2 × SBP + Age FP2 × Diabetes type1 + Age FP2 × Diabetes type2 + Age FP2 × Hypertension.

Fractional polynomials: *Age FP1 (males) =* $(age/10)^{-1}$, *Age FP2 (males) = (age/10)*$^{3}$, *BMI FP 1 (males) =* $(BMI/10)^{-2}$, *BMI FP 2 (males) =* $(BMI/10)^{-2}\ X\ log(BMI/10)$, *Age FP1 (Females) =* $age/10^{-2}$, *Age FP2 (Females) = age/10, BMI FP 1 (Females) =* $(BMI/10)^{-2}$, *BMI FP 2 (Females) =* $(BMI/10)^{-2}\ X\ log\ (BMI/10)$.

**Supplementary Table 26 Reclassification performance for 10-year MACCE risk prediction at a 10% risk threshold comparing Cox clinical with Clinical Deep Survival Network (DSN) models combined with conventional ECG parameters derived from rest, exercise, recovery, and a combination of all three stages intervals**

| Model | Comparator | Net-benefit (95% CI) | Categorical NRI (95% CI) | Continuous NRI (95% CI) |
|---|---|---|---|---|
| **Male** | | | | |
| Cox clinical | Clinical DSN + Resting ECG | -0.012(-0.027 - 0.003) | -0.012 ( -0.027 - 0.002) | **-0.131 ( -0.177 - -0.084) |
| Cox clinical | Clinical DSN + Exercise ECG | -0.011(-0.028 - 0.006) | -0.012 ( -0.029 - 0.005) | -0.0716 ( -0.119 - -0.024) |
| Cox clinical | Clinical DSN + Recovery ECG | 0.009(-0.006 - 0.025) | 0.009 ( -0.006 - 0.024) | **-0.110 ( -0.158 - -0.062) |
| Cox clinical | Clinical DSN + Combined ECG | -0.011(-0.025 - 0.004) | -0.011 ( -0.026 - 0.004) | ** -0.097 ( -0.144 - -0.049) |
| **Female** | | | | |
| Cox clinical | Clinical DSN + Resting ECG | -0.007(-0.03 - 0.016) | 0.008 ( -0.013 - 0.029) | *-0.076 ( -0.146 - -0.006) |
| Cox clinical | Clinical DSN + Exercise ECG | -0.011(-0.038 - 0.014) | 0.020 ( -0.006 - 0.046) | -0.031 ( -0.101 - 0.039) |
| Cox clinical | Clinical DSN + Recovery ECG | -0.018(-0.041 - 0.003) | 0.006 ( -0.016 - 0.027) | -0.021 ( -0.091 - 0.048) |
| Cox clinical | Clinical DSN + Combined ECG | 0.003(-0.019 - 0.025) | 0.011 ( -0.011 - 0.032) | * -0.115 ( -0.185 - -0.045) |

Abbreviations: NRI = Net re classification index, DSN = Deep survival network, CI = Confidence interval
Data are reported on 35,597 participants with measurable ECG parameters, 3,053 (7·4%) MACCE were reported within 10-years. P-values were derived via bootstrapping for 1000 iterations, with a significance threshold of 0·05.

Each row corresponds to a separate risk score, fitted independently with the selected risk factors independent estimates of the baseline survival using Breslow's method, with mean-centred continuous variables, with all binary risk factors set to 0

Age+Sex and Cox clinical risk scores were derived using Cox proportional hazards regression. Extended Cox models additionally included the $ECG_{AI}$ score as a covariate (Age+Sex+ $ECG_{AI}$; Cox clinical+ $ECG_{AI}$).

$ECG_{AI}$ scores were derived from separate deep survival networks trained on raw ECG recordings from: 15 s rest, final 15 s of exercise (peak intensity), final 15 s recovery, and the full 7 min 15 s submaximal ECG, respectively.

Clinical DSN + ECG integrated QRISK3 risk factors and raw ECG data directly within a single deep survival model,

Cox clinical: Age FP1 + Age FP2 + BMI FP1 + BMI FP2 + SBP + SBP5 + Diabetes type 1 + Diabetes type 2 + HDL ratio + Townsend deprivation index + Smoker former + Smoker current + Smoker never + Ethnic Background + Corticosteroid + Migraine + Hypertension + Severe mental illness + Age FP1 × Townsend deprivation index + Age FP2 × Townsend deprivation index + Age FP1 × BMI FP 1 + Age FP1 × BMI FP 2 + Age FP1 × BP + Age FP1 × Diabetes type 1 + Age FP1 × Diabetes type 2 + Age FP1 × hypertension + Age FP2 × BMI FP1 + Age FP2 × BMI FP2 + Age FP2 × SBP + Age FP2 × Diabetes type1 + Age FP2 × Diabetes type2 + Age FP2 × Hypertension.

Fractional polynomials: *Age FP1 (males) =* $(age/10)^{-1}$, *Age FP2 (males) = (age/10)*$^{3}$, *BMI FP 1 (males) =* $(BMI/10)^{-2}$, *BMI FP 2 (males) =* $(BMI/10)^{-2}$ *X* $log(BMI/10)$, *Age FP1 (Females) =* $age/10^{-2}$, *Age FP2 (Females) = age/10, BMI FP 1 (Females) =* $(BMI/10)^{-2}$, *BMI FP 2 (Females) =* $(BMI/10)^{-2}$ *X* $log\ (BMI/10)$.

Clinical DSN: Age, BMI, systolic blood-pressure, SBP5, Diabetes type 1, Diabetes type 2, HDL ratio, Townsend deprivation index, Smoker former, Smoker current, Smoker never, Ethnic Background, Corticosteroid, Migraine, Hypertension, Severe mental illness.

### Section C: Model coefficients, formulas and usage

#### Table 1C Cox clinical

| Risk Factor | Beta (SD) | SE (SD) | Fisher's p-value |
|---|---|---|---|
| **Cox clinical Male 10-year Baseline survival = 0.9161849602568788 (0.0005)** | | | |
| Corticosteroid | -0.350024 (0.265888) | 0.825519 (0.143430) | 0.958 |
| Ethnic Background | 0.322027 (0.032464) | 0.082256 (0.000262) | 0.000 |
| Migraine | 0.184532 (0.221974) | 0.656670 (0.063183) | 0.957 |
| Smoker current | 0.430147 (0.020874) | 0.068663 (0.000522) | 0.000 |
| Smoker former | 0.044320 (0.027935) | 0.046955 (0.000217) | 0.280 |
| Townsend deprivation index | 0.002594 (0.004377) | 0.008761 (0.000064) | 0.936 |
| Age FP 1 | -8.281537 (1.401022) | 3.997923 (0.035174) | 0.000 |
| Age FP 2 | 0.005247 (0.000401) | 0.001232 (0.000006) | 0.000 |
| BMI FP1 | 6.623690 (0.889086) | 3.541770 (0.065505) | 0.002 |
| BMI FP2 | -21.853227 (1.710794) | 6.882238 (0.111236) | 0.000 |
| Bp | 0.007739 (0.001017) | 0.001745 (0.000027) | 0.000 |
| Diabetes type 1 | 0.163047 (0.298926) | 0.383420 (0.091198) | 0.448 |
| Diabetes type 2 | 0.734839 (0.086062) | 0.162680 (0.006020) | 0.000 |
| HDL ratio | 0.114304 (0.003476) | 0.019567 (0.000135) | 0.000 |
| Hypertension | 0.449234 (0.058902) | 0.110537 (0.004092) | 0.000 |
| Rheumatoid Arthritis | 0.410248 (0.198477) | 0.400636 (0.031606) | 0.186 |
| SBP5 | 0.000081 (0.000149) | 0.000249 (0.000006) | 0.791 |
| Severe mental illness | -0.375736 (0.320073) | 0.521429 (0.096206) | 0.705 |
| Age FP1 × Townsend deprivation index | 0.389646 (0.859947) | 1.237686 (0.019319) | 0.655 |
| Age FP2 × Townsend deprivation index | 0.000208 (0.000262) | 0.000385 (0.000005) | 0.579 |
| Age FP 1 × BMI FP 1 | 180.825332 (140.377654) | 507.026874 (13.27348) | 0.964 |
| Age FP 1 × BMI FP 2 | 6.137352 (276.787202) | 1005.158734 (20.5467) | 0.996 |
| Age FP 1 × SBP | 0.559086 (0.038647) | 0.244762 (0.003328) | 0.000 |
| Age FP 1 × Diabetes type 1 | 1.025598 (34.260935) | 52.580719 (16.247853) | 0.917 |
| Age FP 1 × Diabetes type 2 | 31.543949 (10.307556) | 22.061615 (1.103530) | 0.026 |
| Age FP 1 × hypertension | 3.441496 (9.051975) | 15.118433 (0.697914) | 0.898 |
| Age FP 2 × BMI FP 1 | 0.021124 (0.050468) | 0.161500 (0.003384) | 0.985 |
| Age FP 2 × BMI FP 2 | 0.076797 (0.084614) | 0.319911 (0.005387) | 0.986 |
| Age FP 2 × SBP | 0.000127 (0.000018) | 0.000072 (0.000001) | 0.004 |
| Age FP 2 × Diabetes type 1 | -0.001408 (0.010203) | 0.017267 (0.003672) | 0.947 |
| Age FP 2 × Diabetes type 2 | 0.005581 (0.003391) | 0.006605 (0.000189) | 0.402 |
| Age FP 2 × hypertension | 0.000346 (0.002649) | 0.004225 (0.000148) | 0.887 |
| **Cox clinical Female baseline survival = 0.9642753682636174 (0.001)** | | | |
| Corticosteroid | -2.131646 (5.104756) | 114.333002 (226.662800) | 0.975 |
| Ethnic Background | 0.042184 (0.102007) | 0.137585 (0.004452) | 0.604 |
| Migraine | -0.650955 (0.273151) | 0.827189 (0.142690) | 0.617 |
| Smoker current | 0.582761 (0.048526) | 0.108828 (0.000903) | 0.000 |
| Smoker former | 0.062849 (0.027752) | 0.066280 (0.000827) | 0.321 |
| Townsend deprivation index | -0.002225 (0.009944) | 0.012890 (0.000183) | 0.698 |
| Age FP 1 | 23.438344 (9.796006) | 21.238341 (0.514248) | 0.163 |
| Age FP 2 | 1.064908 (0.096568) | 0.238237 (0.003999) | 0.000 |

| | | | |
|---|---|---|---|
| BMI FP1 | -3.412544 (1.597298) | 3.771709 (0.098042) | 0.387 |
| BMI FP2 | -2.647527 (3.095258) | 7.461164 (0.191163) | 0.900 |
| bp | 0.005591 (0.001549) | 0.002388 (0.000074) | 0.000 |
| Diabetes type 1 | -0.529207 (3.636028) | 1.235246 (1.539416) | 0.000 |
| Diabetes type 2 | -0.923512 (0.744213) | 0.841830 (0.469805) | 0.312 |
| HDL ratio | 0.120117 (0.001921) | 0.027162 (0.000197) | 0.000 |
| Hypertension | 0.553000 (0.078572) | 0.194864 (0.013675) | 0.000 |
| Rheumatoid Arthritis | 0.414370 (0.251212) | 0.382957 (0.043465) | 0.084 |
| SBP5 | -0.000233 (0.000073) | 0.000324 (0.000005) | 0.657 |
| Severe mental illness | 0.849891 (0.181746) | 0.513339 (0.058582) | 0.004 |
| Age FP1 × Townsend deprivation index | 0.231371 (3.668057) | 6.230333 (0.149250) | 0.898 |
| Age FP2 × Townsend deprivation index | -0.010007 (0.042433) | 0.072340 (0.001443) | 0.919 |
| Age FP 1 × BMI FP 1 | -644.273512 (1396.518951) | 1870.052338 (107.76346) | 0.775 |
| Age FP 1 × BMI FP 2 | 907.220093 (2798.051262) | 3717.504987 (203.6192) | 0.759 |
| Age FP 1 × SBP | 0.583749 (0.487352) | 1.108127 (0.029894) | 0.803 |
| Age FP 1 × Diabetes type 1 | -1012.073225 (1708.487561) | 556.827521 (625.965467) | 0.180 |
| Age FP 1 × Diabetes type 2 | -682.034530 (304.368144) | 404.067616 (175.569874) | 0.007 |
| Age FP 1 × hypertension | -93.313401 (40.250325) | 93.878203 (5.374447) | 0.307 |
| Age FP 2 × BMI FP 1 | -7.057003 (16.845839) | 21.728503 (1.039829) | 0.730 |
| Age FP 2 × BMI FP 2 | 12.53618 (33.51847) | 43.638486 (1.967705) | 0.731 |
| Age FP 2 × SBP | 0.006074 (0.005385) | 0.012155 (0.000239) | 0.829 |
| Age FP 2 × Diabetes type 1 | -14.682401 (23.657940) | 7.404937 (8.656602) | 0.068 |
| Age FP 2 × Diabetes type 2 | -6.854104 (2.579645) | 3.826777 (1.377621) | 0.003 |
| Age FP 2 × hypertension | -1.025834 (0.357974) | 0.946764 (0.038957) | 0.219 |

Abbreviations: FP= Fractional polynomial, BMI = Boday mass index, SBP5 = Standard deviation of two blood pressure readings, HDL = High-density lipoprotein, SBP = Systolic blood pressure
Results are presented as mean value (Standard deviation)
Beta estimates and standard errors are derived from the mean value across 5-cross validation folds.
P values are derived across 5-cross validation folds using fisher's approach.
Age FP 1 (men) = $(age/10)^{-1}$
Age FP 2 (men) = $(age/10)^3$
BMI FP 1 (men) = $(BMI/10)^{-2}$
BMI FP 2 (men) = $(BMI/10)^{-2}\ X \log\left(\frac{BMI}{10}\right)$
Age FP 1 (Women) = $age/10^{-2}$
Age FP 2 (Women) = age/10
BMI FP 1 (Women) = $(BMI/10)^{-2}$
BMI FP 2 (Women) = $(BMI/10)^{-2}\ X \log\left(\frac{BMI}{10}\right)$

#### Table 2C Cox clinical + $ECG_{AI}$

| Risk Factor | Beta (SD) | SE (SD) | Fisher's p-value |
|---|---|---|---|
| **Cox clinical + $ECG_{AI}$ Male 10-year Baseline survival = 0.9166735485427895 (0.0005)** | | | |
| Corticosteroid | -0.370238 (0.266196) | 0.825496 (0.143416) | 0.946 |
| Ethnic Background | 0.290444 (0.032006) | 0.082346 (0.000279) | 0.000 |
| Migraine | 0.210158 (0.221488) | 0.656778 (0.063105) | 0.945 |
| Smoker current | 0.436015 (0.020976) | 0.068675 (0.000530) | 0.000 |
| Smoker former | 0.046825 (0.027336) | 0.046968 (0.000219) | 0.233 |
| Townsend deprivation index | 0.002671 (0.004375) | 0.008753 (0.000066) | 0.934 |
| Age FP 1 | -8.518373 (1.295476) | 4.011238 (0.033318) | 0.000 |
| Age FP 2 | 0.005080 (0.000381) | 0.001237 (0.000006) | 0.000 |
| BMI FP1 | 6.757441 (0.875046) | 3.533547 (0.066149) | 0.001 |
| BMI FP2 | -20.898572 (1.745010) | 6.874848 (0.113803) | 0.000 |
| SBP | 0.007043 (0.001052) | 0.001755 (0.000028) | 0.000 |
| Diabetes type 1 | 0.194762 (0.293970) | 0.382739 (0.089915) | 0.375 |
| Diabetes type 2 | 0.735350 (0.087638) | 0.162933 (0.006129) | 0.000 |
| HDL ratio | 0.113551 (0.003666) | 0.019612 (0.000138) | 0.000 |
| Hypertension | 0.432364 (0.058689) | 0.110793 (0.004159) | 0.000 |
| Rheumatoid Arthritis | 0.415374 (0.203391) | 0.400724 (0.031577) | 0.174 |
| SBP5 | 0.000057 (0.000147) | 0.000249 (0.000006) | 0.851 |
| Severe mental illness | -0.440769 (0.327000) | 0.521602 (0.096207) | 0.546 |
| $ECG_{AI}$ | 0.462110 (0.043070) | 0.096530 (0.003854) | 0.000 |
| Age FP1 × Townsend deprivation index | 0.485253 (0.833823) | 1.238102 (0.019558) | 0.653 |
| Age FP2 × Townsend deprivation index | 0.000239 (0.000253) | 0.000386 (0.000005) | 0.568 |
| Age FP 1 × BMI FP 1 | 171.723203 (138.168174) | 507.958201 (13.061271) | 0.972 |
| Age FP 1 × BMI FP 2 | -6.817696 (258.165962) | 1007.034792 (20.238167) | 0.997 |
| Age FP 1 × SBP | 0.554449 (0.034274) | 0.246882 (0.003440) | 0.000 |
| Age FP 1 × Diabetes type 1 | 1.746634 (33.619637) | 52.596934 (16.070136) | 0.918 |
| Age FP 1 × Diabetes type 2 | 30.617253 (10.630635) | 22.185117 (1.149150) | 0.035 |
| Age FP 1 × hypertension | 3.280180 (9.148926) | 15.158096 (0.699693) | 0.891 |
| Age FP2 × BMI FP1 | 0.018698 (0.049549) | 0.161648 (0.003429) | 0.987 |
| Age FP2 × BMI FP2 | 0.075541 (0.080288) | 0.320294 (0.005579) | 0.989 |
| Age FP2 × SBP | 0.000124 (0.000019) | 0.000073 (0.000001) | 0.006 |
| Age FP 2 × Diabetes type1 | -0.001344 (0.010102) | 0.017343 (0.003665) | 0.951 |
| Age FP 2 × Diabetes type2 | 0.005360 (0.003489) | 0.006643 (0.000198) | 0.442 |
| Age FP 2 × Hypertension | 0.000250 (0.002659) | 0.004238 (0.000147) | 0.878 |
| Age FP 1 ×$ECG_{AI}$ | -6.395473 (5.826748) | 13.747958 (0.504315) | 0.803 |
| Age FP 2 ×$ECG_{AI}$ | -0.000762 (0.001674) | 0.004066 (0.000144) | 0.946 |
| **Cox clinical + $ECG_{AI}$ Female 10-year Baseline survival = 0.9644312655209759 (0.001)** | | | |

| | | | |
|---|---|---|---|
| Corticosteroid | -2.139429 (5.103004) | 114.458049 (226.912626) | 0.977 |
| Ethnic Background | 0.022929 (0.104888) | 0.137888 (0.004484) | 0.632 |
| Migraine | -0.677080 (0.270921) | 0.827188 (0.142713) | 0.575 |
| Smoker current | 0.590358 (0.049218) | 0.108917 (0.000905) | 0.000 |
| Smoker former | 0.064222 (0.027305) | 0.066286 (0.000832) | 0.300 |
| Townsend deprivation index | -0.002337 (0.009850) | 0.012894 (0.000183) | 0.707 |
| Age FP 1 | 23.825739 (9.609024) | 21.283544 (0.515667) | 0.154 |
| Age FP 2 | 1.064066 (0.094973) | 0.238916 (0.004066) | 0.000 |
| BMI FP1 | -3.530345 (1.637779) | 3.773620 (0.097908) | 0.348 |
| BMI FP2 | -1.658469 (3.252635) | 7.478075 (0.191622) | 0.933 |
| SBP | 0.005278 (0.001563) | 0.002389 (0.000073) | 0.000 |
| Diabetes type 1 | -0.454064 (3.522284) | 1.230700 (1.530636) | 0.000 |
| Diabetes type 2 | -0.973155 (0.721668) | 0.828864 (0.455175) | 0.218 |
| HDL ratio | 0.117952 (0.002153) | 0.027192 (0.000207) | 0.000 |
| Hypertension | 0.547235 (0.078572) | 0.194440 (0.013951) | 0.000 |
| Rheumatoid Arthritis | 0.417022 (0.256338) | 0.383005 (0.043505) | 0.078 |
| SBP5 | -0.000227 (0.000072) | 0.000323 (0.000006) | 0.681 |
| Severe mental illness | 0.790257 (0.169078) | 0.514784 (0.058477) | 0.011 |
| $ECG_{AI}$ | 0.384236 (0.072835) | 0.148093 (0.005023) | 0.000 |
| Age FP1 × Townsend deprivation index | 0.123873 (3.783475) | 6.217625 (0.147836) | 0.889 |
| Age FP2 × Townsend deprivation index | -0.010818 (0.043583) | 0.072232 (0.001433) | 0.910 |
| Age FP 1 × BMI FP 1 | -732.526109 (1397.753777) | 1875.995740 (105.434279) | 0.758 |
| Age FP 1 × BMI FP 2 | 1058.890455 (2764.128820) | 3732.372230 (200.383356) | 0.773 |
| Age FP 1 × SBP | 0.566389 (0.469419) | 1.109153 (0.028284) | 0.821 |
| Age FP 1 × Diabetes type 1 | -985.923818 (1647.445015) | 552.423937 (620.415592) | 0.185 |
| Age FP 1 × Diabetes type 2 | -694.088226 (287.494510) | 398.602823 (169.015913) | 0.004 |
| Age FP 1 × hypertension | -90.168697 (41.267719) | 93.623455 (5.479881) | 0.340 |
| Age FP2 × BMI FP1 | -7.792538 (16.906717) | 21.780690 (1.022958) | 0.731 |
| Age FP2 × BMI FP2 | 13.437998 (33.337130) | 43.767508 (1.946698) | 0.740 |
| Age FP2 × SBP | 0.006018 (0.005299) | 0.012177 (0.000225) | 0.837 |
| Age FP 2 × Diabetes type1 | -14.340859 (22.826342) | 7.348574 (8.584039) | 0.068 |
| Age FP 2 × Diabetes type2 | -6.953663 (2.430628) | 3.784072 (1.320469) | 0.002 |
| Age FP 2 × Hypertension | -0.990301 (0.366134) | 0.944880 (0.039521) | 0.251 |

| | | | |
|---|---|---|---|
| Age FP 1 ×$ECG_{AI}$ | -4.638380 (36.636766) | 72.965922 (2.715443) | 0.929 |
| Age FP 2 ×$ECG_{AI}$ | -0.227338 (0.419115) | 0.824399 (0.025195) | 0.904 |

Abbreviations: FP= Fractional polynomial, BMI = Boday mass index, SBP5 = Standard deviation of two blood pressure readings, HDL = High-density lipoprotein, SBP = Systolic blood pressure
Results are presented as mean value (Standard deviation)
Beta estimates and standard errors are derived from the mean value across 5-cross validation folds.
P values are derived across 5-cross validation folds using fisher's approach.

Age FP 1 (men) = $(age/10)^{-1}$
Age FP 2 (men) = $(age/10)^3$
BMI FP 1 (men) = $(BMI/10)^{-2}$
BMI FP 2 (men) = $(BMI/10)^{-2}\ X \log\left(\frac{BMI}{10}\right)$

Age FP 1 (Women) = $age/10^{-2}$
Age FP 2 (Women) = age/10
BMI FP 1 (Women) = $(BMI/10)^{-2}$
BMI FP 2 (Women) = $(BMI/10)^{-2}\ X \log\left(\frac{BMI}{10}\right)$

#### Table 3C Cox clinical in sub cohort of 35,597 with available conventional ECG parameters

| Risk Factor | Beta Estimate (SD) | Standard Error (SD) | Fisher's p-value |
|---|---|---|---|
| **Cox clinical Male 10-year Baseline survival = 0.9194677672636795 (0.0008)** | | | |
| Corticosteroid | -3.058324 (4.821383) | 93.970671 (185.938480) | 0.868 |
| Ethnic Background | 0.231215 (0.026039) | 0.094921 (0.000765) | 0.000 |
| Migraine | -2.969078 (4.840460) | 88.402198 (174.802476) | 0.927 |
| Smoker current | 0.409467 (0.030721) | 0.074885 (0.000529) | 0.000 |
| Smoker former | 0.032706 (0.033036) | 0.050601 (0.000222) | 0.552 |
| Townsend deprivation index | 0.003743 (0.003855) | 0.009491 (0.000066) | 0.933 |
| Age FP 1 | -8.239010 (1.216513) | 4.335058 (0.026215) | 0.001 |
| Age FP 2 | 0.005252 (0.000353) | 0.001333 (0.000004) | 0.000 |
| BMI FP1 | 5.737902 (1.501906) | 4.179753 (0.194800) | 0.048 |
| BMI FP2 | -20.451175 (2.824710) | 7.976247 (0.271968) | 0.000 |
| bp | 0.008276 (0.001047) | 0.001891 (0.000021) | 0.000 |
| Diabetes type 1 | -0.155624 (0.290918) | 0.489431 (0.086359) | 0.934 |
| Diabetes type 2 | 0.829499 (0.111686) | 0.180215 (0.006348) | 0.000 |
| HDL ratio | 0.126299 (0.004742) | 0.021151 (0.000133) | 0.000 |
| Hypertension | 0.470482 (0.060258) | 0.117152 (0.005449) | 0.000 |
| Rheumatoid Arthritis | 0.487297 (0.194260) | 0.427857 (0.028182) | 0.120 |
| SBP5 | -0.000004 (0.000131) | 0.000271 (0.000006) | 0.957 |
| Severe mental illness | -0.335986 (0.501976) | 0.616577 (0.194329) | 0.913 |
| Age FP1 × Townsend deprivation index | 0.739400 (0.788227) | 1.333724 (0.023397) | 0.672 |
| Age FP2 × Townsend deprivation index | 0.000366 (0.000247) | 0.000415 (0.000007) | 0.334 |
| Age FP 1 × BMI FP 1 | 378.082310 (163.583901) | 588.273681 (33.062337) | 0.739 |
| Age FP 1 × BMI FP 2 | -445.675590 (341.704069) | 1146.118123 (47.757791) | 0.950 |
| Age FP 1 × SBP | 0.346595 (0.009164) | 0.264453 (0.002704) | 0.084 |

| | | | |
|---|---|---|---|
| Age FP 1 × Diabetes type 1 | 30.476196 (26.056294) | 59.072283 (14.485844) | 0.705 |
| Age FP 1 × Diabetes type 2 | 25.773126 (8.472790) | 23.913016 (1.128218) | 0.203 |
| Age FP 1 × hypertension | 14.765519 (9.519468) | 15.957665 (0.960540) | 0.248 |
| Age FP 2 × BMI FP 1 | 0.096373 (0.071765) | 0.188144 (0.009526) | 0.859 |
| Age FP 2 × BMI FP 2 | -0.088298 (0.129062) | 0.366196 (0.013420) | 0.978 |
| Age FP 2 × SBP | 0.000063 (0.000013) | 0.000078 (0.000001) | 0.548 |
| Age FP 2 × Diabetes type 1 | 0.011983 (0.008168) | 0.019220 (0.002877) | 0.676 |
| Age FP 2 × Diabetes type 2 | 0.003589 (0.002686) | 0.007168 (0.000226) | 0.867 |
| Age FP 2 × hypertension | 0.003347 (0.002564) | 0.004511 (0.000196) | 0.486 |
| **Cox clinical Female baseline survival = 0.9674000393032509 (0.003)** | | | |
| Corticosteroid | -1.859872 (5.116782) | 121.300363 (240.596065) | 0.824 |
| Ethnic Background | -0.089060 (0.115363) | 0.167134 (0.005411) | 0.625 |
| Migraine | -3.411411 (4.984203) | 107.695642 (213.386305) | 0.605 |
| Smoker current | 0.550494 (0.052170) | 0.122283 (0.001990) | 0.000 |
| Smoker former | 0.093190 (0.025772) | 0.072182 (0.001013) | 0.077 |
| Townsend deprivation index | 0.001645 (0.011488) | 0.014240 (0.000256) | 0.636 |
| Age FP 1 | 16.735322 (12.314826) | 25.469804 (0.914644) | 0.584 |
| Age FP 2 | 1.051904 (0.107542) | 0.276632 (0.006315) | 0.000 |
| BMI FP1 | -6.897666 (0.837690) | 4.921792 (0.277077) | 0.050 |
| BMI FP2 | 1.489601 (1.523853) | 9.248453 (0.371809) | 0.998 |
| Bp | 0.004204 (0.001637) | 0.002628 (0.000061) | 0.007 |
| Diabetes type 1 | -24.687989 (50.004439) | 1189.912221 (2377.291972) | 0.553 |
| Diabetes type 2 | -0.960672 (0.749314) | 1.223582 (0.535338) | 0.701 |
| HDL ratio | 0.126130 (0.006158) | 0.029783 (0.000297) | 0.000 |
| Hypertension | 0.580602 (0.112253) | 0.202014 (0.022301) | 0.000 |
| Rheumatoid Arthritis | -0.086852 (0.269346) | 0.574247 (0.075469) | 0.939 |

| | | | |
|---|---|---|---|
| SBP5 | -0.000379 (0.000139) | 0.000363 (0.000009) | 0.246 |
| Severe mental illness | 0.851659 (0.185567) | 0.578152 (0.075784) | 0.017 |
| Age FP1 × Townsend deprivation index | -1.638954 (3.466888) | 7.031087 (0.181435) | 0.928 |
| Age FP2 × Townsend deprivation index | -0.033317 (0.038805) | 0.080747 (0.001885) | 0.862 |
| Age FP 1 × BMI FP 1 | -3783.364927 (756.469300) | 2586.829714 (147.201885) | 0.033 |
| Age FP 1 × BMI FP 2 | 6329.416234 (1418.786650) | 4886.174778 (221.731868) | 0.085 |
| Age FP 1 × SBP | 0.775537 (0.551594) | 1.250151 (0.025152) | 0.720 |
| Age FP 1 × Diabetes type 1 | -11416.091500 (22425.393370) | 531331.050924 (1061545.206705) | 0.981 |
| Age FP 1 × Diabetes type 2 | -786.445935 (291.852989) | 588.448527 (203.441580) | 0.075 |
| Age FP 1 × hypertension | -46.702277 (55.296550) | 100.715091 (10.086351) | 0.819 |
| Age FP 2 × BMI FP 1 | -35.685588 (9.610732) | 28.285309 (1.360998) | 0.096 |
| Age FP 2 × BMI FP 2 | 64.022863 (19.900540) | 54.558957 (2.090358) | 0.145 |
| Age FP 2 × SBP | 0.009105 (0.006177) | 0.013623 (0.000206) | 0.658 |
| Age FP 2 × Diabetes type 1 | -150.563739 (296.919332) | 6968.439068 (13926.117170) | 0.969 |
| Age FP 2 × Diabetes type 2 | -7.883906 (2.401493) | 5.406887 (1.598062) | 0.036 |
| Age FP 2 × hypertension | -0.570664 (0.519654) | 1.018803 (0.075489) | 0.732 |

Abbreviations: FP= Fractional polynomial, BMI = Boday mass index, SBP5 = Standard deviation of two blood pressure readings, HDL = High-density lipoprotein, SBP = Systolic blood pressure
Results are presented as mean value (Standard deviation)
Beta estimates and standard errors are derived from the mean value across 5-cross validation folds.
P values are derived across 5-cross validation folds using fisher's approach.

Age FP 1 (men) = $(age/10)^{-1}$

Age FP 2 (men) = $(\text{age}/10)^3$

BMI FP 1 (men) = $(BMI/10)^{-2}$

BMI FP 2 (men) = $(BMI/10)^{-2}\ X \log\left(\frac{BMI}{10}\right)$

Age FP 1 (Women) = $age/10^{-2}$

Age FP 2 (Women) = age/10

BMI FP 1 (Women) = $(BMI/10)^{-2}$

BMI FP 2 (Women) = $(BMI/10)^{-2}\ X \log\left(\frac{BMI}{10}\right)$

**Table 4C Cox clinical + Conventional ECG parameters in a sub cohort of 35,597 with available conventional ECG**

| Risk Factor | Beta Estimate (SD) | Standard Error (SD) | Fisher's p-value |
|---|---|---|---|
| **Cox clinical + ECG Male 10-year Baseline survival = 0.921173895014286 (0.0006)** | | | |
| Corticosteroid | -3.053305 (4.928603) | 103.763729 (205.523500) | 0.907 |
| Ethnic Background | 0.250329 (0.032275) | 0.096403 (0.000771) | 0.000 |
| Migraine | -2.977797 (4.910584) | 94.050879 (186.098164) | 0.939 |
| Smoker current | 0.393048 (0.030481) | 0.075608 (0.000497) | 0.000 |
| Smoker former | 0.031042 (0.033947) | 0.050802 (0.000236) | 0.559 |
| Townsend deprivation index | 0.004286 (0.003832) | 0.009566 (0.000061) | 0.901 |
| Age FP 1 | -8.417533 (0.762423) | 4.793997 (0.034306) | 0.005 |
| Age FP 2 | 0.004361 (0.000229) | 0.001452 (0.000008) | 0.000 |
| BMI FP1 | 5.271504 (1.474828) | 4.231986 (0.193271) | 0.096 |
| BMI FP2 | -18.155179 (2.846709) | 8.084144 (0.276214) | 0.000 |
| SBP | 0.007690 (0.000911) | 0.001911 (0.000022) | 0.000 |
| Diabetes type 1 | -0.178577 (0.275120) | 0.489687 (0.085230) | 0.935 |
| Diabetes type 2 | 0.842291 (0.101461) | 0.183713 (0.005994) | 0.000 |
| HDL ratio | 0.134318 (0.004345) | 0.021270 (0.000140) | 0.000 |
| Hypertension | 0.445556 (0.057521) | 0.118724 (0.005726) | 0.000 |
| Rheumatoid Arthritis | 0.438986 (0.181216) | 0.429511 (0.027734) | 0.215 |
| SBP5 | -0.000083 (0.000158) | 0.000271 (0.000005) | 0.798 |
| Severe mental illness | -0.387358 (0.510011) | 0.618803 (0.193933) | 0.859 |
| Age FP1 × Townsend deprivation index | 0.845549 (0.748499) | 1.353870 (0.022023) | 0.668 |
| Age FP2 × Townsend deprivation index | 0.000416 (0.000239) | 0.000421 (0.000006) | 0.233 |
| Age FP 1 × BMI FP 1 | 369.867793 (180.171687) | 597.982099 (32.585041) | 0.768 |
| Age FP 1 × BMI FP 2 | -419.371622 (327.293821) | 1166.765160 (49.373498) | 0.966 |
| Age FP 1 × SBP | 0.348686 (0.033844) | 0.269527 (0.003464) | 0.090 |
| Age FP 1 × Diabetes type 1 | 24.486686 (26.253692) | 59.814473 (14.027401) | 0.811 |
| Age FP 1 × Diabetes type 2 | 27.424743 (7.995900) | 24.413108 (1.004441) | 0.175 |
| Age FP 1 × hypertension | 11.603035 (9.715555) | 16.296883 (1.000434) | 0.483 |
| Age FP2 × BMI FP1 | 0.095773 (0.081485) | 0.190143 (0.009574) | 0.859 |
| Age FP2 × BMI FP2 | -0.085509 (0.136296) | 0.371030 (0.014183) | 0.983 |
| Age FP2 × SBP | 0.000069 (0.000019) | 0.000079 (0.000001) | 0.454 |
| Age FP 2 × Diabetes type1 | 0.009794 (0.007708) | 0.019483 (0.002827) | 0.797 |
| Age FP 2 × Diabetes type2 | 0.004038 (0.002644) | 0.007263 (0.000207) | 0.821 |
| Age FP 2 × Hypertension | 0.002422 (0.002649) | 0.004583 (0.000195) | 0.735 |
| Resting QRS interval | -0.001491 (0.002469) | 0.004261 (0.000231) | 0.771 |

| | | | |
|---|---|---|---|
| Resting QTc interval | 0.000274 (0.000965) | 0.001771 (0.000044) | 0.892 |
| Resting Anisotropy evidence | -0.014432 (0.019842) | 0.052518 (0.002170) | 0.937 |
| Resting Atrial Tachycardia evidence | 0.008658 (0.005951) | 0.017767 (0.000811) | 0.894 |
| Resting PVC count | 0.017749 (0.013509) | 0.065853 (0.012905) | 0.973 |
| Resting regularity Evidence | 0.021694 (0.007705) | 0.025395 (0.000503) | 0.465 |
| Exercise PR interval | -0.001065 (0.000418) | 0.001418 (0.000012) | 0.604 |
| Exercise QRS interval | -0.004797 (0.002576) | 0.007670 (0.000163) | 0.752 |
| Exercise QTc interval | 0.000648 (0.000422) | 0.001196 (0.000097) | 0.796 |
| Exercise RR Interval | 0.002148 (0.000268) | 0.000633 (0.000012) | 0.000 |
| Exercise Atrial Tachycardia evidence | 0.009421 (0.001185) | 0.008089 (0.000417) | 0.158 |
| Exercise PVC count | 0.007653 (0.023160) | 0.060136 (0.004161) | 0.979 |
| Exercise regularity evidence | 0.013677 (0.007157) | 0.017741 (0.000129) | 0.550 |
| Recovery Irregularity evidence | 0.013567 (0.005374) | 0.024248 (0.000731) | 0.833 |
| Recovery QRS interval | 0.005704 (0.003537) | 0.007506 (0.000155) | 0.538 |
| Recovery QTc interval | 0.002429 (0.000491) | 0.002024 (0.000173) | 0.132 |
| Recovery Atrial Tachycardia evidence | 0.001788 (0.003885) | 0.015750 (0.000312) | 0.994 |
| Recovery regularity evidence | 0.018562 (0.004559) | 0.022748 (0.000214) | 0.536 |
| Age FP 1 × Resting QRS interval | -0.099509 (0.283266) | 0.645164 (0.021378) | 0.937 |
| Age FP 1 × Resting QTc interval | 0.186839 (0.140490) | 0.249661 (0.009373) | 0.492 |
| Age FP 1 × Resting Anisotropy evidence | -7.323903 (2.947289) | 6.967514 (0.350952) | 0.237 |
| Age FP 1 × Resting Atrial tachycardia evidence | 1.478490 (0.626584) | 2.413532 (0.137146) | 0.769 |
| Age FP 1 × Resting PVC count | 1.877899 (5.945336) | 10.002778 (2.321090) | 0.789 |
| Age FP 1 × Resting regularity evidence | 0.081740 (1.813632) | 3.541294 (0.099657) | 0.960 |
| Age FP 1 × Exercise PR interval | 0.175616 (0.042983) | 0.199580 (0.000926) | 0.450 |
| Age FP 1 × Exercise QRS interval | 0.563302 (0.349615) | 1.115918 (0.016077) | 0.870 |
| Age FP 1 × Exercise QTc interval | -0.164137 (0.067765) | 0.176132 (0.013207) | 0.392 |
| Age FP 1 × Exercise RR interval | 0.004875 (0.049124) | 0.084489 (0.001378) | 0.870 |
| Age FP 1 × Exercise Atrial tachycardia evidence | 1.186696 (0.473332) | 1.060608 (0.048358) | 0.166 |
| Age FP 1 × Exercise PVC count | 0.619285 (3.045315) | 7.871910 (0.260239) | 0.980 |
| Age FP 1 × Exercise regularity evidence | -2.131547 (0.641735) | 2.470027 (0.022713) | 0.464 |
| Age FP 1 × Recovery Irregularity evidence | -2.946675 (1.156957) | 3.452341 (0.054666) | 0.461 |
| Age FP 1 × Recovery QRS interval | -0.006782 (0.493191) | 1.089161 (0.013787) | 0.948 |
| Age FP 1 × Recovery QTc interval | -0.015081 (0.035581) | 0.269611 (0.034398) | 1.000 |
| Age FP 1 × Recovery Atrial tachycardia evidence | 1.276569 (0.764689) | 2.185623 (0.021819) | 0.787 |
| Age FP 1 × Recovery regularity evidence | -2.922880 (0.683230) | 3.215600 (0.042486) | 0.414 |
| Age FP 2 × Resting QRS interval | -0.000021 (0.000084) | 0.000185 (0.000008) | 0.944 |
| Age FP 2 × Resting QTc interval | 0.000061 (0.000040) | 0.000079 (0.000002) | 0.498 |

| | | | |
|---|---|---|---|
| Age FP 2 × Resting Anisotropy evidence | -0.002281 (0.000956) | 0.001948 (0.000093) | 0.125 |
| Age FP 2 × Resting Atrial tachycardia evidence | 0.000569 (0.000310) | 0.000711 (0.000044) | 0.485 |
| Age FP 2 × Resting PVC count | 0.000969 (0.001750) | 0.002840 (0.000462) | 0.731 |
| Age FP 2 × Resting regularity evidence | -0.000113 (0.000538) | 0.001056 (0.000027) | 0.931 |
| Age FP 2 × Exercise PR interval | 0.000051 (0.000011) | 0.000059 (0.000000) | 0.478 |
| Age FP 2 × Exercise QRS interval | 0.000110 (0.000102) | 0.000323 (0.000003) | 0.967 |
| Age FP 2 × Exercise QTc interval | -0.000060 (0.000020) | 0.000056 (0.000003) | 0.239 |
| Age FP 2 × Exercise RR interval | 0.000012 (0.000014) | 0.000024 (0.000000) | 0.721 |
| Age FP 2 × Exercise Atrial tachycardia evidence | 0.000382 (0.000179) | 0.000300 (0.000015) | 0.078 |
| Age FP 2 × Exercise PVC count | 0.000388 (0.000901) | 0.001987 (0.000107) | 0.941 |
| Age FP 2 × Exercise regularity evidence | -0.000237 (0.000157) | 0.000762 (0.000010) | 0.980 |
| Age FP 2 × Recovery Irregularity evidence | -0.000857 (0.000370) | 0.000985 (0.000019) | 0.431 |
| Age FP 2 × Recovery QRS interval | 0.000109 (0.000172) | 0.000315 (0.000003) | 0.788 |
| Age FP 2 × Recovery QTc interval | -0.000025 (0.000011) | 0.000090 (0.000006) | 0.990 |
| Age FP 2 × Recovery Atrial tachycardia evidence | 0.000389 (0.000200) | 0.000616 (0.000005) | 0.744 |
| Age FP 2× Recovery regularity evidence | -0.001205 (0.000263) | 0.000948 (0.000010) | 0.093 |
| **Cox clinical + ECG Female baseline survival = 0.9693602558896434 (0.003)** | | | |
| Corticosteroid | -1.976697 (5.132148) | 124.923166 (247.831108) | 0.907 |
| Ethnic Background | -0.081440 (0.124964) | 0.168970 (0.005530) | 0.613 |
| Migraine | -3.609918 (5.165232) | 132.426195 (262.841494) | 0.482 |
| Smoker current | 0.507017 (0.060319) | 0.123494 (0.001923) | 0.000 |
| Smoker former | 0.077114 (0.024866) | 0.072560 (0.000997) | 0.224 |
| Townsend deprivation index | -0.000473 (0.011481) | 0.014366 (0.000259) | 0.676 |
| Age FP 1 | 3.721850 (16.802543) | 27.610876 (1.292915) | 0.858 |
| Age FP 2 | 0.857354 (0.158731) | 0.299144 (0.009086) | 0.000 |
| BMI FP1 | -7.514021 (1.135243) | 4.942824 (0.250173) | 0.023 |
| BMI FP2 | 3.057241 (2.200450) | 9.325692 (0.332151) | 0.978 |
| SBP | 0.003717 (0.001581) | 0.002632 (0.000054) | 0.028 |
| Diabetes type 1 | -22.379224 (45.542274) | 869.822657 (1737.289983) | 0.515 |
| Diabetes type 2 | -0.986806 (0.728800) | 1.228215 (0.579733) | 0.662 |
| HDL ratio | 0.126508 (0.005539) | 0.029975 (0.000306) | 0.000 |
| Hypertension | 0.126508 (0.005539) | 0.029975 (0.000306) | 0.000 |
| Rheumatoid Arthritis | 0.126508 (0.005539) | 0.029975 (0.000306) | 0.000 |
| SBP5 | -0.000341 (0.000142) | 0.000363 (0.000011) | 0.355 |

| | | | |
|---|---|---|---|
| Severe mental illness | 0.740934 (0.222092) | 0.583057 (0.076613) | 0.054 |
| Age FP1 × Townsend deprivation index | -2.087138 (3.108249) | 7.119993 (0.194404) | 0.932 |
| Age FP2 × Townsend deprivation index | -0.034975 (0.035063) | 0.081635 (0.001986) | 0.892 |
| Age FP 1 × BMI FP 1 | -3371.361814 (555.995914) | 2553.405342 (118.897499) | 0.077 |
| Age FP 1 × BMI FP 2 | 5473.807363 (1037.883455) | 4838.456657 (176.877494) | 0.189 |
| Age FP 1 × SBP | 0.538023 (0.503669) | 1.266520 (0.020029) | 0.916 |
| Age FP 1 × Diabetes type 1 | -10352.721262 (20406.986824) | 388406.333520 (775759.628090) | 0.991 |
| Age FP 1 × Diabetes type 2 | -720.655938 (264.868341) | 581.468761 (218.054415) | 0.115 |
| Age FP 1 × hypertension | -57.926764 (47.699574) | 100.693454 (9.196135) | 0.793 |
| Age FP2 × BMI FP1 | -30.370274 (7.966515) | 28.066404 (1.143397) | 0.222 |
| Age FP2 × BMI FP2 | 53.301410 (16.675324) | 54.326897 (1.744596) | 0.323 |
| Age FP2 × SBP | 0.006292 (0.005611) | 0.013833 (0.000169) | 0.879 |
| Age FP 2 × Diabetes type1 | -136.878917 (270.553390) | 5093.740868 (10177.089348) | 0.983 |
| Age FP 2 × Diabetes type2 | -7.099495 (2.127908) | 5.317743 (1.683223) | 0.068 |
| Age FP 2 × Hypertension | -0.688814 (0.442698) | 1.021964 (0.069304) | 0.693 |
| Resting QRS interval | -0.001761 (0.003582) | 0.006059 (0.000251) | 0.801 |
| Resting QTc interval | -0.002664 (0.000842) | 0.002806 (0.000044) | 0.351 |
| Resting Anisotropy evidence | -0.008412 (0.044730) | 0.074704 (0.004472) | 0.812 |
| Resting Atrial Tachycardia evidence | -0.002372 (0.009682) | 0.026471 (0.000984) | 0.973 |
| Resting PVC count | -0.114437 (0.290023) | 0.185674 (0.139764) | 0.815 |
| Resting regularity Evidence | 0.040423 (0.022615) | 0.037707 (0.000746) | 0.169 |
| Exercise PR interval | -0.000776 (0.001236) | 0.002382 (0.000053) | 0.884 |
| Exercise QRS interval | -0.012047 (0.004132) | 0.010324 (0.000601) | 0.147 |
| Exercise QTc interval | -0.003578 (0.001225) | 0.002502 (0.000174) | 0.039 |
| Exercise RR Interval | 0.002660 (0.000473) | 0.001001 (0.000040) | 0.000 |
| Exercise Atrial Tachycardia evidence | -0.005038 (0.002353) | 0.013421 (0.000504) | 0.964 |
| Exercise PVC count | -0.069540 (0.055870) | 0.087795 (0.018104) | 0.554 |
| Exercise regularity evidence | -0.033686 (0.004360) | 0.025142 (0.000513) | 0.069 |
| Recovery Irregularity evidence | 0.005679 (0.021770) | 0.036617 (0.001334) | 0.850 |
| Recovery QRS interval | 0.019844 (0.004846) | 0.010762 (0.000358) | 0.002 |
| Recovery QTc interval | 0.006335 (0.001739) | 0.003505 (0.000052) | 0.002 |

| | | | |
|---|---|---|---|
| Recovery Atrial Tachycardia evidence | 0.008599 (0.008700) | 0.022401 (0.000637) | 0.946 |
| Recovery regularity evidence | 0.030303 (0.023593) | 0.033592 (0.000497) | 0.231 |
| Age FP 1 × Resting QRS interval | 4.685456 (0.594696) | 3.009125 (0.067449) | 0.018 |
| Age FP 1 × Resting QTc interval | -0.176955 (0.349030) | 1.407983 (0.035738) | 0.993 |
| Age FP 1 × Resting Anisotropy evidence | 8.352663 (18.032953) | 30.291483 (2.122153) | 0.813 |
| Age FP 1 × Resting Atrial tachycardia evidence | 8.835450 (2.459381) | 11.138656 (0.469317) | 0.558 |
| Age FP 1 × Resting PVC count | -94.513262 (124.159024) | 84.552187 (62.640771) | 0.358 |
| Age FP 1 × Resting regularity evidence | -20.554959 (4.125426) | 17.691073 (0.536867) | 0.163 |
| Age FP 1 × Exercise PR interval | 2.199981 (0.298083) | 1.150989 (0.032806) | 0.001 |
| Age FP 1 × Exercise QRS interval | -6.852584 (1.632544) | 5.038986 (0.231211) | 0.052 |
| Age FP 1 × Exercise QTc interval | 0.574648 (0.351687) | 1.179718 (0.116625) | 0.874 |
| Age FP 1 × Exercise RR interval | 0.416345 (0.162504) | 0.482972 (0.016394) | 0.453 |
| Age FP 1 × Exercise Atrial tachycardia evidence | 5.560724 (3.275170) | 5.985036 (0.417762) | 0.261 |
| Age FP 1 × Exercise PVC count | 3.089575 (31.518151) | 39.706749 (13.408190) | 0.750 |
| Age FP 1 × Exercise regularity evidence | 2.454565 (7.280798) | 12.243339 (0.449209) | 0.869 |
| Age FP 1 × Recovery Irregularity evidence | 22.590040 (7.618744) | 16.768233 (0.871248) | 0.055 |
| Age FP 1 × Recovery QRS interval | 2.098954 (1.396131) | 5.444468 (0.145017) | 0.954 |
| Age FP 1 × Recovery QTc interval | -0.539083 (0.308646) | 1.735982 (0.047608) | 0.983 |
| Age FP 1 × Recovery Atrial tachycardia evidence | -19.636307 (4.706738) | 10.856460 (0.370094) | 0.002 |
| Age FP 1 × Recovery regularity evidence | 25.542789 (4.782398) | 16.519767 (0.376644) | 0.018 |
| Age FP 2 × Resting QRS interval | 0.043897 (0.002105) | 0.031205 (0.000893) | 0.048 |
| Age FP 2 × Resting QTc interval | -0.003399 (0.003709) | 0.015837 (0.000385) | 0.990 |
| Age FP 2 × Resting Anisotropy evidence | 0.092205 (0.211727) | 0.356851 (0.016620) | 0.838 |
| Age FP 2 × Resting Atrial tachycardia evidence | 0.103347 (0.029834) | 0.131377 (0.003768) | 0.563 |
| Age FP 2 × Resting PVC count | -1.079614 (1.026884) | 0.875222 (0.493449) | 0.172 |
| Age FP 2 × Resting regularity evidence | -0.262693 (0.048790) | 0.199977 (0.005325) | 0.078 |
| Age FP 2 × Exercise PR interval | 0.023647 (0.003339) | 0.012813 (0.000302) | 0.002 |
| Age FP 2 × Exercise QRS interval | -0.062718 (0.016338) | 0.056917 (0.002275) | 0.192 |
| Age FP 2 × Exercise QTc interval | 0.008246 (0.004597) | 0.014017 (0.000899) | 0.787 |
| Age FP 2 × Exercise RR interval | 0.004504 (0.001331) | 0.005247 (0.000145) | 0.472 |
| Age FP 2 × Exercise Atrial tachycardia evidence | 0.076592 (0.033659) | 0.065696 (0.002800) | 0.110 |
| Age FP 2 × Exercise PVC count | 0.023321 (0.312937) | 0.445748 (0.106361) | 0.782 |
| Age FP 2 × Exercise regularity evidence | 0.036292 (0.080110) | 0.140957 (0.002857) | 0.880 |
| Age FP 2 × Recovery Irregularity evidence | 0.295465 (0.087365) | 0.191575 (0.009956) | 0.017 |
| Age FP 2 × Recovery QRS interval | 0.019746 (0.015226) | 0.060462 (0.001508) | 0.974 |

| | | | |
|---|---|---|---|
| Age FP 2 × Recovery QTc interval | -0.005112 (0.005899) | 0.018979 (0.000831) | 0.975 |
| Age FP 2 × Recovery Atrial tachycardia evidence | -0.255495 (0.055515) | 0.122944 (0.004240) | 0.000 |
| Age FP 2 × Recovery regularity evidence | 0.309347 (0.058496) | 0.185464 (0.004070) | 0.008 |

Abbreviations: FP= Fractional polynomial, BMI = Body mass index, SBP5 = Standard deviation of two blood pressure readings, HDL = High-density lipoprotein, SBP = Systolic blood pressure, PVC = Premature ventricular contraction
Results are presented as mean value (Standard deviation)
Beta estimates and standard errors are derived from the mean value across 5-cross validation folds.
P values are derived across 5-cross validation folds using fisher's approach.
Age FP 1 (men) = $(age/10)^{-1}$
Age FP 2 (men) = $(age/10)^3$
BMI FP 1 (men) = $(BMI/10)^{-2}$
BMI FP 2 (men) = $(BMI/10)^{-2}\ X \log\left(\frac{BMI}{10}\right)$
Age FP 1 (Women) = $age/10^{-2}$
Age FP 2 (Women) = age/10
BMI FP 1 (Women) = $(BMI/10)^{-2}$
BMI FP 2 (Women) = $(BMI/10)^{-2}\ X \log\left(\frac{BMI}{10}\right)$

### Section D: TRIPOD guidelines for predictive model development.

TRIPOD Checklist: Prediction Model Development

| Section/Topic | Item | Checklist Item | Page |
|---|---|---|---|
| **Title and abstract** | | | |
| Title | 1 | Identify the study as developing and/or validating a multivariable prediction model, the target population, and the outcome to be predicted. | **p.1** |
| Abstract | 2 | Provide a summary of objectives, study design, setting, participants, sample size, predictors, outcome, statistical analysis, results, and conclusions. | **p.2** |
| **Introduction** | | | |
| Background and objectives | 3a | Explain the medical context (including whether diagnostic or prognostic) and rationale for developing or validating the multivariable prediction model, including references to existing models. | **p.3** |
| | 3b | Specify the objectives, including whether the study describes the development or validation of the model or both. | **p.3** |
| **Methods** | | | |
| Source of data | 4a | Describe the study design or source of data (e.g., randomized trial, cohort, or registry data), separately for the development and validation data sets, if applicable. | **p.5** |
| | 4b | Specify the key study dates, including start of accrual; end of accrual; and, if applicable, end of follow-up. | **p.5, p.7** |
| Participants | 5a | Specify key elements of the study setting (e.g., primary care, secondary care, general population) including number and location of centres. | **p.5** |
| | 5b | Describe eligibility criteria for participants. | **p.6, p.8** |
| | 5c | Give details of treatments received, if relevant. | **.** |
| Outcome | 6a | Clearly define the outcome that is predicted by the prediction model, including how and when assessed. | **p.6-p.7** |
| | 6b | Report any actions to blind assessment of the outcome to be predicted. | |
| Predictors | 7a | Clearly define all predictors used in developing or validating the multivariable prediction model, including how and when they were measured. | **p.5, p.7** |
| | 7b | Report any actions to blind assessment of predictors for the outcome and other predictors. | **.** |
| Sample size | 8 | Explain how the study size was arrived at. | **p.8** |
| Missing data | 9 | Describe how missing data were handled (e.g., complete-case analysis, single imputation, multiple imputation) with details of any imputation method. | **p.8** |
| Statistical analysis methods | 10a | Describe how predictors were handled in the analyses. | **p.6-p.7** |
| | 10b | Specify type of model, all model-building procedures (including any predictor selection), and method for internal validation. | **p.7** |
| | 10d | Specify all measures used to assess model performance and, if relevant, to compare multiple models. | **p.8** |
| Risk groups | 11 | Provide details on how risk groups were created, if done. | **p.8** |
| **Results** | | | |
| Participants | 13a | Describe the flow of participants through the study, including the number of participants with and without the outcome and, if applicable, a summary of the follow-up time. A diagram may be helpful. | **p.24** |
| | 13b | Describe the characteristics of the participants (basic demographics, clinical features, available predictors), including the number of participants with missing data for predictors and outcome. | **p.9, P.19** |

| | | | |
|---|---|---|---|
| Model development | 14a | Specify the number of participants and outcome events in each analysis. | **p.10, P.24** |
| | 14b | If done, report the unadjusted association between each candidate predictor and outcome. | **p.10, Appendix P. 35s** |
| Model specification | 15a | Present the full prediction model to allow predictions for individuals (i.e., all regression coefficients, and model intercept or baseline survival at a given time point). | **Appendix p.56 – p.69.** |
| | 15b | Explain how to the use the prediction model. | **p.6** |
| Model performance | 16 | Report performance measures (with CIs) for the prediction model. | **p.10-p.12** |
| **Discussion** | | | |
| Limitations | 18 | Discuss any limitations of the study (such as nonrepresentative sample, few events per predictor, missing data). | **p.14** |
| Interpretation | 19b | Give an overall interpretation of the results, considering objectives, limitations, and results from similar studies, and other relevant evidence. | **p.13-p.14** |
| Implications | 20 | Discuss the potential clinical use of the model and implications for future research. | **p.14** |
| **Other information** | | | |
| Supplementary information | 21 | Provide information about the availability of supplementary resources, such as study protocol, Web calculator, and data sets. | **p.14** |
| Funding | 22 | Give the source of funding and the role of the funders for the present study. | **p.9** |
